# Human Plasma Proteomics Links Modifiable Lifestyle Exposome to Disease Risk

**DOI:** 10.1101/2025.05.07.25327178

**Authors:** Shakson Isaac, Randall J. Ellis, Yon Ho Jee, Venkatesh L. Murthy, Miriam S. Udler, Benjamin M. Neale, Shamil Sunyaev, Alexander Gusev, Alicia R. Martin, Chirag J. Patel

## Abstract

Lifestyle exposures influence disease risk, yet the molecular signatures linking modifiable behaviors to human biology remain incompletely characterized. Here we show that the plasma proteome carries a measurable record of modifiable lifestyle exposures. We developed the Human Exposomic Architecture of the Proteome (HEAP), a framework linking lifestyle exposures, plasma proteins, and disease risk at population scale. Applying HEAP to more than 50,000 UK Biobank participants, 2,686 plasma proteins, and 169 lifestyle exposures, we identified exposure-associated protein signatures that organized along shared health-behavior patterns and connected lifestyle variation to disease-relevant biology. Integrating disease associations, mediation analyses, Mendelian randomization, and colocalization showed that only a minority of exposure-responsive proteins were consistent with causal intermediates, whereas many appeared to function as biological reporters of exposure burden, disease liability, and early disease processes. Comparisons with exercise-training and glucagon-like peptide-1 receptor agonist studies linked observational lifestyle signatures to intervention-induced proteomic changes, suggesting that part of the exposomic signal reflects biology modifiable by intervention. Furthermore, proteome-based exposure scores captured exposure-associated biology, tracked behavioral change over time, and predicted disease risk. Overall, HEAP provides a resource for advancing disease prevention and precision medicine by linking modifiable lifestyle exposures to disease-relevant protein signatures.

## Introduction

Cumulative lifestyle exposures^1^ such as diet, exercise, and smoking are well-recognized potential contributors to disease risk, but their precise biological mechanisms remain poorly understood^2–4^. Circulating plasma proteomics offers a powerful measurement tool to uncover these mechanisms, as proteins reflect key biological processes, including lipid and hormone transport, enzymatic activity, signaling, and immune functions^5^. Fluctuations in protein expression are linked to disease mechanisms and serve as biomarkers for chronic conditions, covering metabolic disorders, cardiovascular disease, and neurodegenerative diseases ^6–8^. Uncovering the effects of modifiable exposures and their interactions with genetics is crucial for developing effective prevention strategies, assessing interventions, and improving health outcomes.

Large-scale biobank studies leveraging high-throughput plasma proteomics now enable systematic investigation of the mechanisms linking genetics, the exposome, and disease. For example, the largest genetic study of the plasma proteome, using the UK Biobank, cataloged over 14,000 protein quantitative trait loci (pQTLs) and 1,100 gene-by-environment (GxE) interactions across nearly 3,000 plasma proteins in approximately 50,000 individuals^9,10^. Exposome-wide association studies (ExWAS) have traditionally identified lifestyle, chemical, and social factors associated with disease risk ^11,12^, but often without consideration of biological mediators. Recent studies have begun integrating omic profiling to assess exposure effects^13,14^; however, these are often limited by smaller sample sizes, model mis-specifications, and lack of consideration for multiple exposures simultaneously, leading to a fragmented literature of associations^4,15,16^. With large-scale biobanks now storing genetic, exposomic, and molecular data at the individual level, we can begin to disentangle how genetics and exposures drive protein expression and mediate disease risk. We hypothesize that if the lifestyle exposome is relevant to disease risk, its effects must be mediated by biological responses, such as changes in protein expression.

Here we introduce the **H**uman **E**xposomic **A**rchitecture of the **P**roteome (HEAP), a computational framework for systematically characterizing and triangulating how lifestyle exposures shape the plasma proteome. HEAP jointly models lifestyle exposures and inherited genetics to identify exposure-responsive proteins, quantify their relative contributions to protein variation, and prioritize candidate mediators linking lifestyle to disease. To strengthen inference, the framework integrates complementary genetic and interventional evidence^17^. And, as a resource, derives proteome-based exposure scores that enable lifestyle exposures to be inferred directly from circulating protein profiles.

We applied HEAP to the UK Biobank, with 2,686 plasma proteins and 169 exposures across 50,000 participants. We identified a broad landscape of exposure-responsive proteins, defined proteomic signatures of major lifestyle factors, showed these signatures were linked to disease risk, and showed the proteome can reconstruct lifestyle exposures and capture longitudinal changes. Overall, these analyses establish HEAP as a framework and resource for systematically characterizing how lifestyle exposures are reflected in the plasma proteome and connected to human disease.

## Results

### A spectrum of genetic and exposure-driven plasma proteins

HEAP partitions plasma protein variance into covariate, genetic, exposomic, and gene-by-environment components (Fig. 1a; Supplementary Fig. 1). Across the proteome, demographics (e.g., age, sex) explained most protein variation, followed by genetics (Supplementary Fig. 2; Supplementary Table 1; Supplementary Table 2), although 608 proteins carried reproducible aggregate exposure-responsive signal (R^2^_PXS_ ≥ 0.01) (Fig. 1b; Supplementary Table 1). These proteins were enriched for pathways related to adiposity, inflammation, lipid transport, and metabolic regulation (Supplementary Fig. 3; Supplementary Table 3).

**Figure 1:**
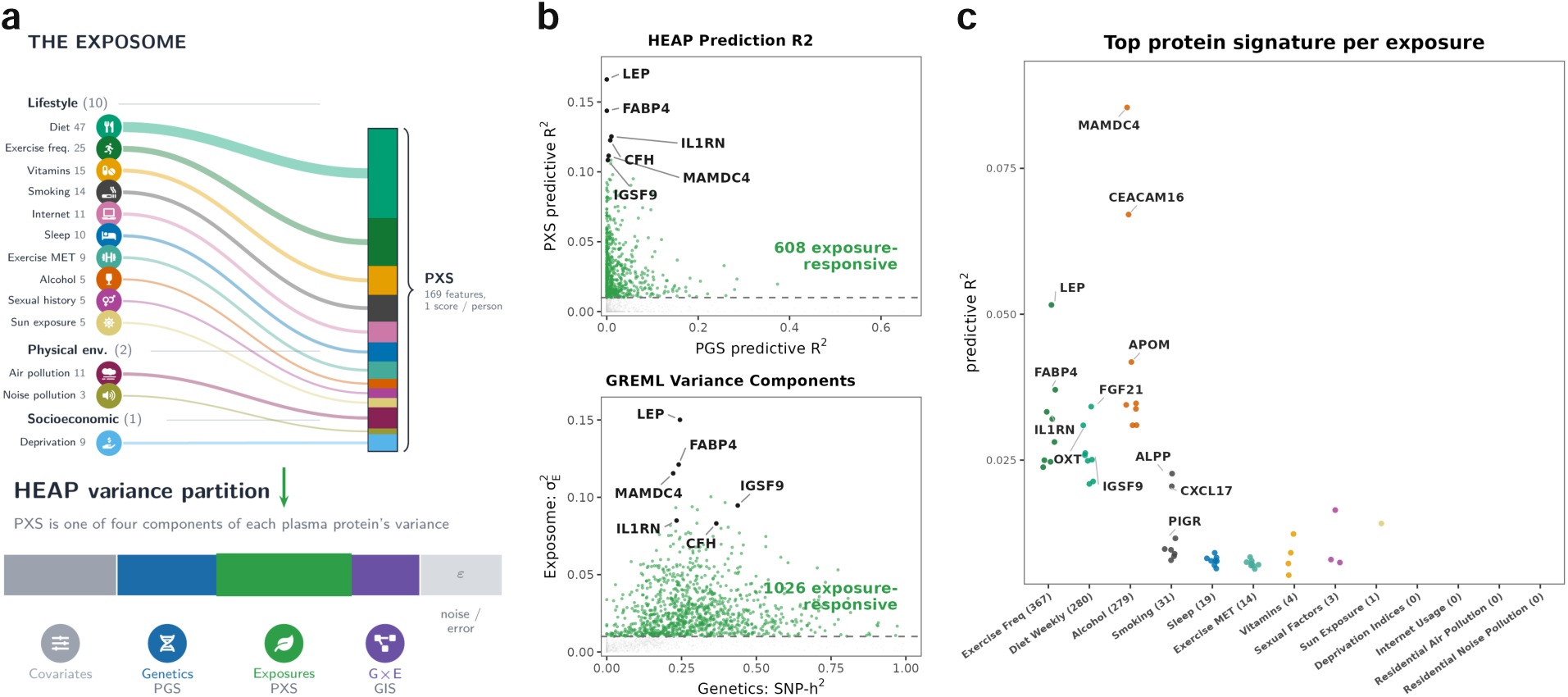
A spectrum of genetic-and exposure-driven plasma proteins. **(a)** HEAP variance partitioning framework. 13 exposure categories (lifestyle, physical-environment, and socioeconomic domains) combine into one per-protein polyexposure score (PXS). Each plasma protein’s variance is partitioned into covariates, a polygenic score (PGS), the exposomic score (PXS), a gene-by-environment term (GIS), and residual noise (*ε*). **(b)** Genetic versus exposomic contribution of each plasma protein, by HEAP cross-validated prediction *R*^2^ and by GREML variance components. Exposure-responsive proteins (exposomic *R*^2^ *≥* 1% of variance; dashed line) are in green with proteins labeled. **(c)** Per-protein exposure-responsive predictive *R*^2^ for each exposure category.

As an independent variance-based comparison, we used genome-based restricted maximum likelihood (GREML) ^18^ to estimate genetic, exposomic, and gene-by-environment contributions from genetic and exposure similarity between individuals (see Methods). GREML yielded concordant estimates of exposure-associated variance and identified 1,026 exposure-responsive proteins, supporting the HEAP framework (Fig. 1b; Supplementary Fig. 4; Supplementary Table 4). In contrast, gene-by-environment explained variation was consistently small, did not reproduce across both approaches, and showed no enriched pathways (Supplementary Fig. 5; Supplementary Fig. 4; Supplementary Table 5; Supplementary Table 3; Supplementary Table 1).

Exposure-responsive variance was concentrated in major lifestyle domains (Fig. 1c; Supplementary Table 2; Supplementary Table 6). Exercise, diet, alcohol, and smoking explained the largest number of proteins and generated distinct signatures (Supplementary Fig. 6). HEAP estimates generalized well to held-out data and remained robust across alternative model specifications and penalized estimators, with attenuation primarily among adiposity-associated proteins after BMI adjustment (Supplementary Fig. 7; Supplementary Table 7; Supplementary Fig. 8; Supplementary Data 1).

### The exposome shapes the proteome, largely independent of genetics

To map lifestyle effects across the proteome, we tested every exposure–protein pair under an 80/20 train/test split. We report associations that were Bonferroni-significant in both train and test subsamples (Supplementary Table 8). This identified 22,240 replicated exposure–protein associations spanning 127 exposures and 1,533 proteins, with exercise frequency, weekly diet, and smoking contributing the largest number of associations (Fig. 2a; Supplementary Fig. 9; Supplementary Table 8).

**Figure 2:**
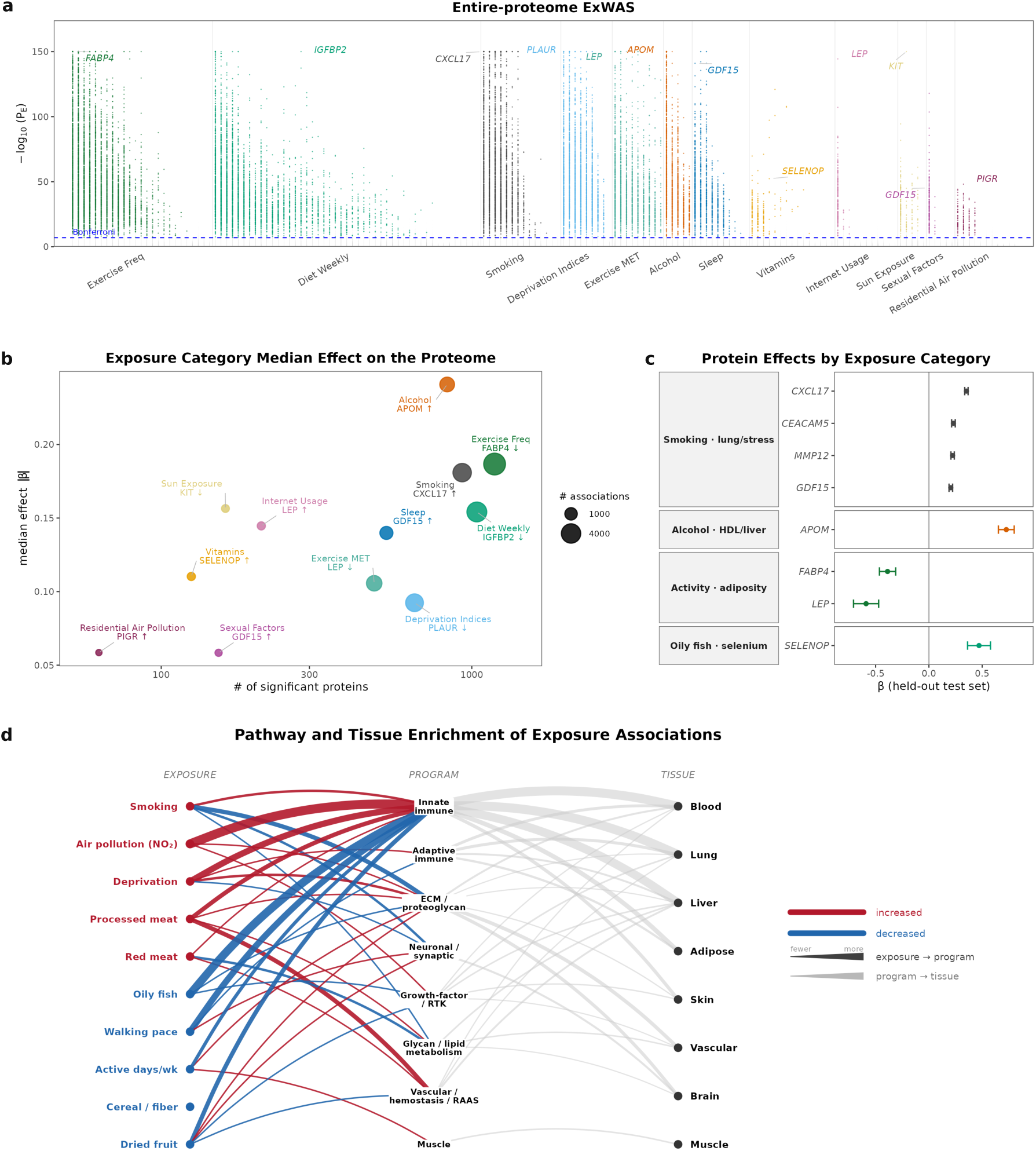
The exposome shapes the proteome, largely independent of genetics. **(a)** Entire-proteome ExWAS. Each point is one exposure–protein pair. 22,240 pairs replicated across 80/20 train/test splits are colored by exposure category and ordered left to right by number of associations. Dashed line marks the Bonferroni threshold. A lead protein is labeled per category. **(b)** Exposome Category Effects. Number of significant proteins versus median effect (*|β|*), with bubble size the number of replicated associations. **(c)** Example exposure–protein associations with smoking, alcohol, physical activity, and oily-fish intake (standardized *β*, 95% CI). **(d)** Exemplar exposure pathway program and tissue enrichments (FDR *q <* 0.05). Exposure*→*program edges are colored by direction (red, increased; blue, decreased) and weighted by number of enriched pathways. Gray program*→*tissue edges are weighted by the number of supporting exposures.

Exposure categories held differing proteomic associations. Across the exposome, lifestyle behaviors, exercise, diet, smoking, and alcohol impacted the greatest numbers of proteins and produced the largest median effect sizes (Fig. 2b). Additionally, effect sizes were dose-dependent. For instance, the mean absolute effect of alcohol on the proteome scaled monotonically with reported alcohol intake frequency (Supplementary Fig. 10).

The resulting exposure–protein associations recovered established exposure biology (Fig. 2c). Smoking was associated with lung and stress-response proteins including CXCL17, CEACAM5, MMP12, and GDF15; alcohol with APOM; physical activity with adiposity-related proteins LEP and FABP4; and oily-fish intake with the selenium carrier SELENOP. Beyond individual proteins, exposures converged on shared biological pathways (Fig. 2d). Putative “harmful” exposures (such as smoking, air pollution, deprivation, and red or processed meat) and hypothesized protective exposures (including physical activity, oily fish, cereal fiber, and dried fruit) were associated with overlapping immune, extracellular-matrix, growth-factor, glycan/lipid, and vascular–hemostatic programs in opposite directions (Supplementary Figs. 11–14; Supplementary Table 9; Supplementary Table 10). These divergent programs ultimately converged on a similar set of target tissues, including the blood, lung, liver, and brain.

Importantly, most exposomic proteomic signatures were robust to covariate adjustment scenarios. Attenuation of effect sizes, however, did occur among adiposity-linked proteins (e.g. LEP and FABP4) after BMI and medication adjustment. More globally, we found replicated exposure-protein associations across covariate specification, including exclusion of individuals with prevalent disease (Supplementary Fig. 15; Supplementary Data 2). Additionally, we tested for polygenic gene-by-environment effects, which were detectable but sparse with 108 replicated associations (Supplementary Table 11). These interactions were largely cis-dominated, concentrated at a few loci (e.g. FOLR3), and less extensive than the exposomic main effects (Supplementary Figs. 16–17; Supplementary Data 3). Together, these findings indicate widespread associations between the lifestyle exposome and the circulating proteome, largely independent of measured genetics.

### Exposure-responsive proteins connect lifestyle exposures to disease risk

To connect exposure-responsive proteins to disease, we developed Genetic and Exposomic Mediation (GEM) to quantify how much of the association between an exposure and incident disease is statistically accounted for by a measured protein (Fig. 3a). GEM uses g-computation over Cox models to decompose each exposure–disease association into an indirect effect through the protein (natural indirect effect, NIE) and a remaining direct effect (NDE) ^19–21^ (Fig. 3a, see Methods). Here, mediation refers to this statistical decomposition and does not establish that the protein is a causal intermediary.

**Figure 3:**
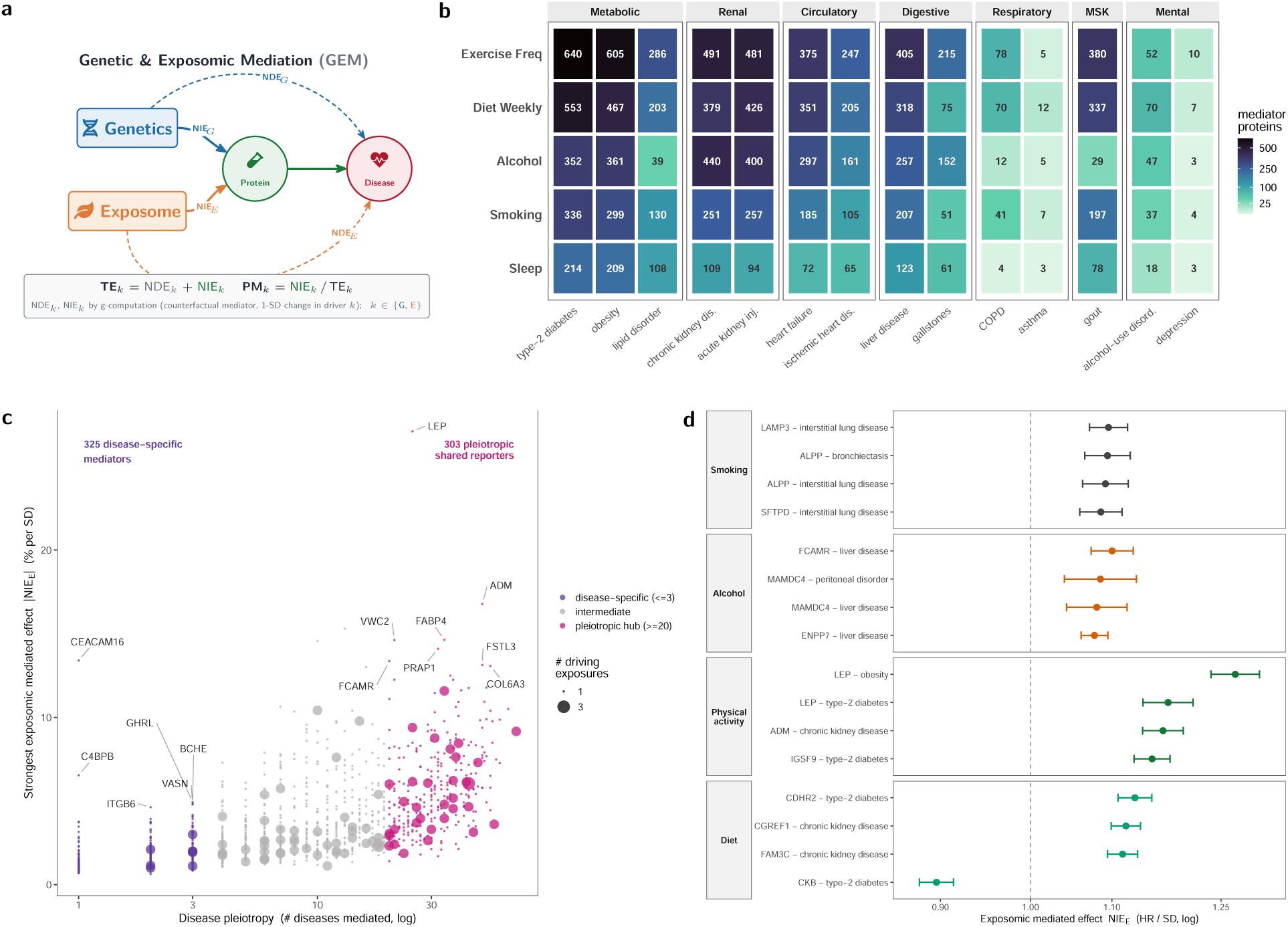
Exposure-responsive proteins connect lifestyle exposures to disease risk. **(a)** Genetic and Exposomic Mediation (GEM) decomposes each exposure–disease association into an indirect effect through a measured plasma protein (NIE), a remaining direct effect (NDE), a total effect (TE), and proportion mediated (PM) = NIE*/*TE. **(b)** Number of exposure-responsive proteins with FDR-significant mediated associations across exposure categories and diseases. **(c)** Proteins ranked by disease pleiotropy reveal a spectrum from disease-specific mediators to highly pleiotropic proteins shared across many diseases. Point size denotes the number of exposure categories associated with each protein. **(d)** Representative exposure–protein–disease triads across smoking–lung, alcohol–liver, physical activity–cardiometabolic, and diet–cardiometabolic axes. Points show mediated hazard ratios per SD with 95% confidence intervals; dashed line indicates HR = 1.

Across 174 incident diseases, 1,448 exposure-associated proteins mediated exposome–disease associations. These associations were densest in cardiometabolic, hepatic, renal, digestive, and respiratory disease (Fig. 3b; Supplementary Table 12; Supplementary Data 4). Individual proteins each accounted for only a small fraction of any one exposure–disease association, suggesting that lifestyle influences disease through shared proteomic responses rather than singular mediators (Supplementary Fig. 18).

This landscape was structured. Ranking proteins by disease pleiotropy revealed a spectrum from 325 disease-specific mediators, confined to one or a few outcomes, to 303 pleiotropic proteins acting across over 20 diseases (Fig. 3c). LEP, ADM, and FABP4 are a few examples of these hubs, linking exposure categories like physical activity to multiple metabolic, renal, and circulatory disease outcomes (Supplementary Fig. 19).

Representative exposure–protein–disease associations recovered expected organ biology (Fig. 3d): smoking through lung proteins such as SFTPD, alcohol through liver proteins such as MAMDC4, and physical activity and diet through cardiometabolic proteins such as LEP and ADM. Although no single protein explained a large proportion of any exposure–disease association, these proteins provided a prioritized set of exposure-responsive candidates linked to modifiable disease risk (Supplementary Fig. 20).

Mediation estimates were stable across penalized estimators and adjustment for blood-draw time (Spearman *ρ* 0.994–1.000), and attenuated under body-mass-index adjustment, the wider clinical covariate set, and exclusion of prevalent disease (*ρ* 0.84–0.87, with 40–49% of base-significant mediators retained). This attenuation is difficult to distinguish from confounding because BMI plausibly lies on the exposure-to-protein path (Supplementary Fig. 21; Supplementary Table 13; Supplementary Data 4).

### Mendelian randomization separates causal mediators from biological reporters

Observational mediation identified proteins associated with exposure and disease, but it cannot distinguish causal mediators from proteins that reflect exposure history, disease liability, or shared upstream causes. We applied bidirectional two-sample Mendelian randomization (MR) ^22^ across HEAP exposure–protein–disease (EPD) triads (Fig. 4a), evaluated all directed relationships, and graded evidence using increasingly stringent criteria incorporating instrument strengths, directionality, sensitivity analyses involving pleiotropy and heterogeneity, along with cross-platform pQTL replication (Fig. 4a, Methods, Table 1). In total we tested 199,646 triads spanning 122 exposures, 818 proteins, and 77 diseases, using 77,573 directed MR tests (Methods).

**Figure 4:**
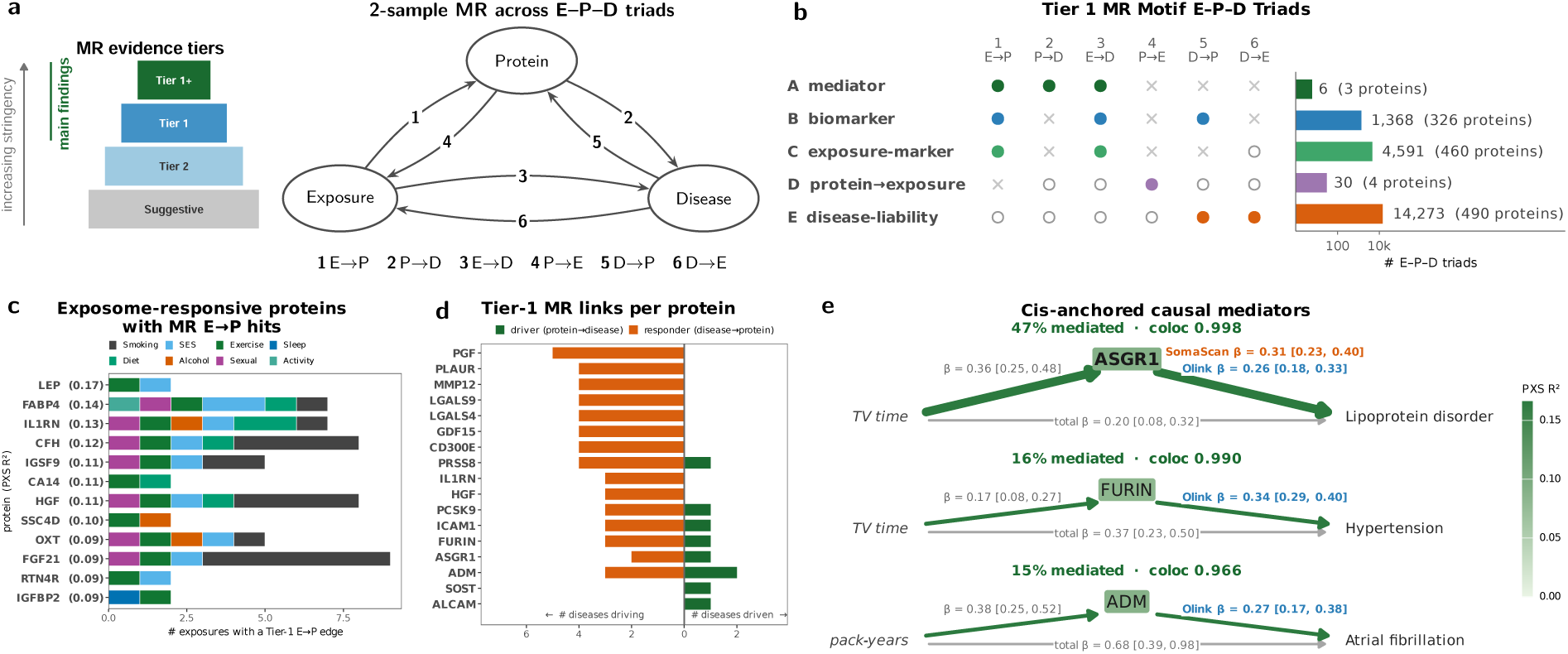
Mendelian randomization separates causal mediators from biological reporters. **(a)** Two-sample MR tests six directed edges among each exposure, protein, and disease (E–P–D triad) and grades the evidence on a stringency ladder: Suggestive (FDR-significant but weakly instrumented, not sensitivity-robust, or of unresolved direction), Tier 2 (FDR-significant), Tier 1 (adds sensitivity robustness and an established causal direction), and Tier 1+ (additionally replicated across the UKB Olink and deCODE SomaScan instruments). **(b)** Each of five MR motifs is a ✓/*×*/*◦* signature over the six edges (✓ supported, *◦* either direction, *×* absent). Bars count the Tier 1 E–P–D triads matching each motif. **(c)** The most exposome-responsive proteins (PXS *R*^2^) that carry MR support, each showing the number of exposures with a Tier 1 or Tier 1+ exposure*→*protein edge, colored by exposure category. **(d)** Example proteins with the number of diseases acting on it (disease*→*protein) versus the number it acts on (protein*→*disease). **(e)** Cis-anchored, colocalized causal mediators: the full exposure*→*protein*→*disease decomposition with *β* [95% CI] on each edge. The total exposure*→*disease effect, percent mediated, and colocalization (PP.H4). The protein*→*disease edge is shown for both instrument arms (UKB Olink, deCODE SomaScan).

**Table 1:** Mendelian randomization confidence tiers. Edges are assigned the highest tier whose criteria they meet, evaluated within each edge type. For exposure–protein–disease motifs, an edge is present only if it reaches Tier 1 or Tier 1+ criteria shown. All other tiers, Tier 2, Reverse, Suggestive, and Null edges are all treated as absent.

| **Tier** | **Criteria** | **Interpretation** |
|---|---|---|
| Tier 1+ | Reaches Tier 1 in UK Biobank and deCODE arms | Robust and cross-arm replicated |
| Tier 1 | FDR-significant, robust (no residual heterogeneity or pleiotropy, or rescued), and the causal direction is established by the Steiger test (directionality *p* < 0.05 in the forward orientation). For cis-pQTL, edge is colocalized (PP.H4 ≥ 0.8) | Robust MR hit |
| Tier 2 | trans-pQTL only instrument, or the cis-pQTL signal fails colocalization (PP.H4 < 0.8) | trans-pQTL MR or cis-pQTL LD confounding |
| Reverse | FDR-significant and robust, but the Steiger test establishes the reverse direction (directionality *p* < 0.05 against the assumed orientation) | Reverse evidence |
| Suggestive | FDR-significant, but fewer than three instruments (non-cis), heterogeneity / pleiotropy not resolved by MR-PRESSO or the weighted median, or a Steiger test that does not establish the causal direction (*p* ≥ 0.05) | Weak evidence |
| Null | Not FDR-significant (*q* ≥ 0.05) | No evidence |

Exposure instruments were derived from genome-wide association studies in UK Biobank participants independent of the pQTL discovery sample, following a sample-independent design ^6^. Instrument strength, single nucleotide polymorphism (SNP) heritability, genomic calibration, and genetic-correlation structure informed which exposures were suitable for MR. Many deprivation and pollution exposures measured on a group of individuals mapped few or no genome-wide loci and could not be instrumented (Supplementary Figs. 22–24). Only a minority of significant edges survived to the highest tier and replicated across both pQTL arms (Supplementary Fig. 25).

Tier 1 MR motifs indicated that causal mediation was rare. Only six triads across three proteins supported the mediator motif (exposure*→*protein*→*disease; Fig. 4b). Most triads supported reporter-like patterns, particularly disease-liability, 14,273 triads across 490 proteins, indicating that disease liability more often influenced protein abundance than proteins driving disease (Supplementary Table 14; Supplementary Table 15; Supplementary Data 5; Supplementary Fig. 26).

MR supported the exposure responsiveness of the proteins prioritized by HEAP. Among the top 50 proteins ranked by PXS *R*^2^, 96% had a supported exposure*→*protein edge (Fig. 4c), indicating potential biomarkers of lifestyle exposures without downstream causal effects on disease. Protein*→*disease effects were rare: eight proteins carried one at Tier 1, each acting on a single disease except ADM (Fig. 4d). Broad disease-responsive proteins included GDF15, LGALS9, IL1RN, and HGF, reinforcing that most mediation-prioritized proteins are biological reporters rather than causal intermediates.

Evidence for mediation were confined to a small set of cis-anchored, colocalized proteins. ASGR1 linked television-watching time to lipoprotein disorders (47% mediated, PP.H4 = 0.998); FURIN linked television-watching time to hypertension (16% mediated, PP.H4 = 0.990); and ADM linked smoking pack-years to atrial fibrillation (15% mediated, PP.H4 = 0.966) (Fig. 4e; Supplementary Table 16). Together, MR refined the observational mediation results into a small set of supported causal mediators and a much larger set of exposure-and disease-responsive biological reporters (Supplementary Figs. 27–29).

### Lifestyle signatures show concordance with exercise and GLP1RA-induced proteomic shifts

We next asked whether exposure-responsive proteins identified observationally reflected biology that changes following intervention (Fig. 5a). We hypothesized that if independent sources of evidence point toward a shared set of proteins, these proteins can contextualize disease links. Many lifestyle-associated proteins (UKB) were enriched for cardiometabolic disease, so we searched to see if these effects were concordant with interventional trials intended to alter cardiometabolic state.

**Figure 5:**
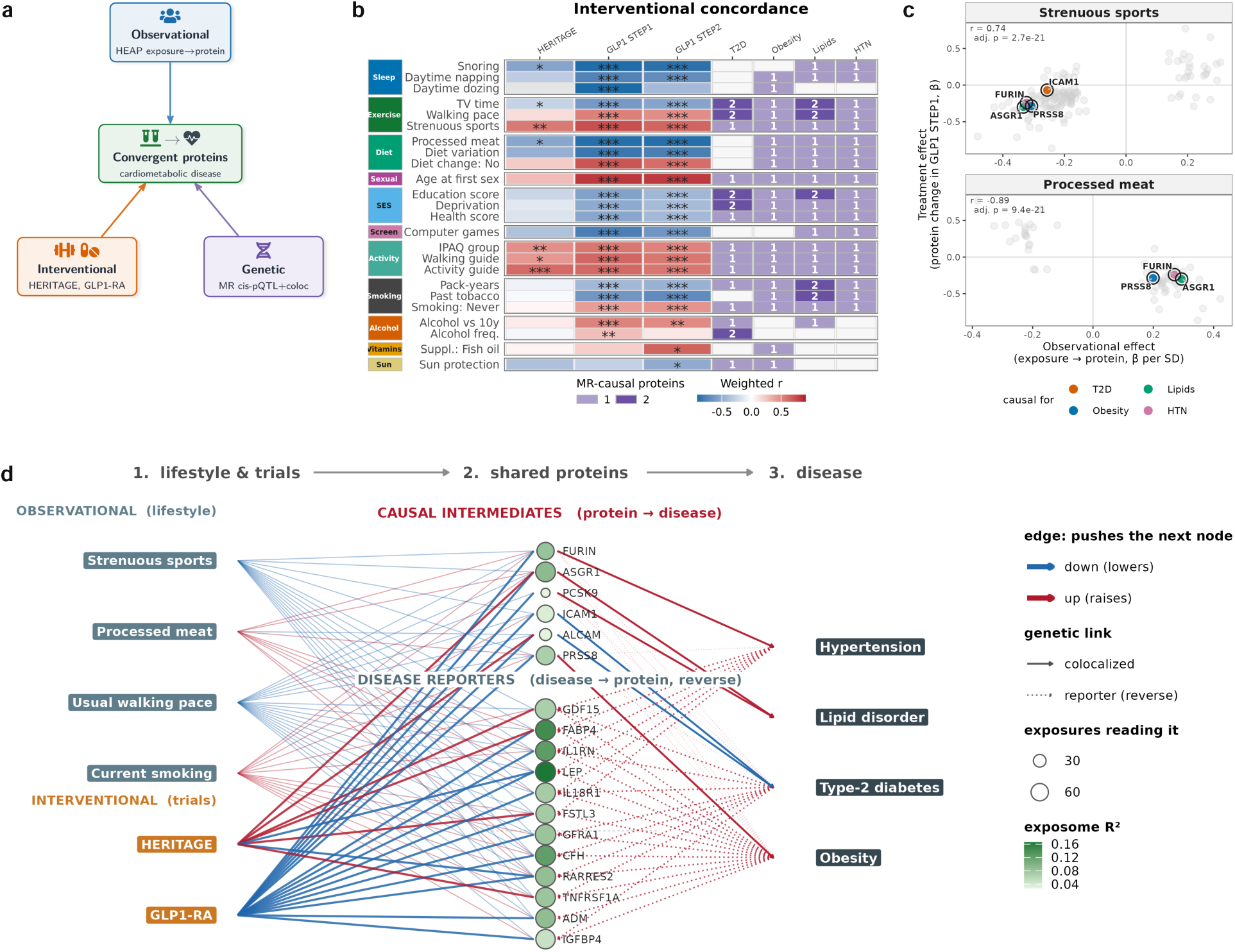
Lifestyle signatures show concordance with exercise and GLP1RA-induced proteomic shifts. **(a)** Analysis design comparing HEAP exposure–protein associations with HERITAGE endurance-training intervention, STEP1/STEP2 GLP1-RA randomized controlled trials, and MR/colocalization hits. **(b)** Concordance heatmap between HEAP exposure signatures and HERITAGE, STEP1, and STEP2 protein shifts. Colors show weighted correlations via Olink/SomaScan concordance adjustment; asterisks denote FDR-adjusted significance (* *P <* 0.05, ** *P <* 0.01, *** *P <* 0.001). Purple cells indicate the number of overlapping Tier 1 cardiometabolic MR proteins per disease. **(c)** Per-protein concordance between UKB exposure–protein associations and GLP1-RA protein shifts for strenuous sports and processed meat. Proteins are colored by Tier 1 MR hits on cardiometabolic disease. Rings mark cis-pQTL colocalization. **(d)** Lifestyle exposures and randomized trials (left) shift a shared set of plasma proteins (middle) linked to cardiometabolic disease (right). Node size = number of associated exposures; node fill = exposome *R*^2^. Proteins split into Tier-1 colocalized cis-pQTL MR with solid edges to disease and dotted edges for disease reporters detected by Tier1 reverse MR. Observational edges are thin, interventional edges are bold. Every edge is colored by the direction of its effect (red raises, blue lowers the node it points to).

We utilized plasma proteomic data from the HERITAGE^23,24^ cohort (n = 654), a 20-week endurance training intervention. Additionally, we incorporated serum proteomic data from 68-week GLP1 receptor agonist (GLP1-RA) treatment in STEP1 (n = 1133) and STEP2 (n = 595) randomized control trials (RCTs) ^25–27^. Across interventions, 97 exposure terms were significantly concordant with at least one trial signature (HERITAGE 31, STEP1 94, STEP2 78), 28 with all three (Fig. 5b; Supplementary Table 17). Physical activity traits, including strenuous sports, usual walking pace, and activity guideline measures, were positively concordant with HERITAGE, supporting the exercise-related HEAP signatures in an independent training intervention. These physical activity traits were also positively concordant with GLP1-RA signatures in STEP1 and STEP2. Diet, smoking, sleep, and screen-time traits also displayed additional concordance with STEP1 or STEP2. However, directional concordance depended on the exposure. For example, strenuous sports showed positive concordance with GLP1-RA protein shifts (*r* = 0.74), whereas processed meat showed inverse concordance (*r* = *−*0.89) (Fig. 5c; Supplementary Data 6). Thus, several observational lifestyle signatures overlapped with proteomic shifts observed after either behavioral intervention or randomized pharmacologic treatment.

We asked whether proteins shared across observational and interventional signatures represented candidate disease mediators or downstream consequences of disease. Overlaying cis-pQTL Mendelian randomization results identified six of the eight Tier-1 colocalized proteins with genetically supported effects on cardiometabolic disease: FURIN (hypertension), ICAM1 and ALCAM (type 2 diabetes), ASGR1 and PCSK9 (lipoprotein disorders), and PRSS8 (obesity). Other exposomic-responsive proteins, including GDF15, LEP, CFH, and FABP4, were downstream disease reporters, detected by reverse MR (Fig. 5d).

### Proteome-based exposure scores read and track lifestyle-associated disease risk

We asked whether proteomic signatures could be used as a quantitative molecular readout of the exposome. For each exposure, we trained a proteome-based exposure score (PES) from baseline plasma proteins and tested whether this PES could read exposure status, track within-person exposure change, and capture disease-relevant exposure biology (Fig. 6a, see Methods). PES were trained in 51,804 baseline participants using nested 5-fold cross-validation and evaluated in 1,210 held-out participants with repeat proteomic measurements.

**Figure 6:**
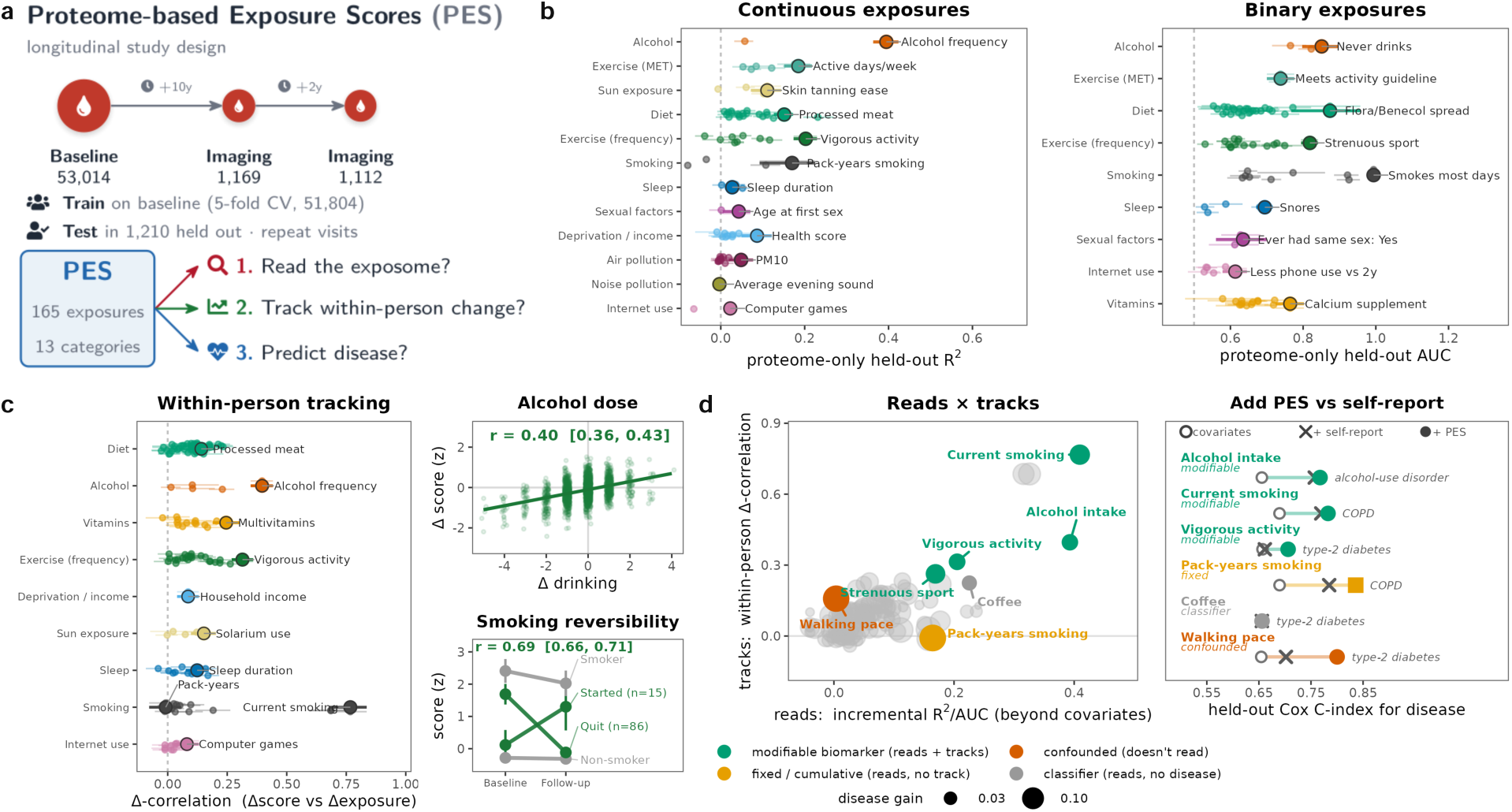
Proteome-based exposure scores read, track, and predict lifestyle-associated disease risk. **(a)** PES study design. Scores were trained from baseline plasma proteins and tested in held-out participants with repeat proteomic measurements. **(b)** Held-out PES performance for continuous and binary exposures, measured by *R*^2^ and AUC respectively (point = mean held-out repeat visits; error bars = 95% bootstrap confidence intervals). **(c)** Held-out within-person PES tracking of exposure change. Pairing each participant’s baseline visit with each repeat imaging visit. Examples of alcohol dose response (continuous) and smoking reversibility (binary) with number of transition counts. **(d)** Relationship between PES exposure reading, longitudinal tracking, and improvement in held-out Cox disease prediction beyond covariates and self-reported exposures.

PES captured distinct proteomic signatures of lifestyle exposures (Fig. 6b). Proteome-only models predicted multiple continuous and binary exposures, with the strongest performance for alcohol, diet, exercise, and smoking-related exposures, and the weakest performance for air and noise pollution. PES models improved exposure prediction beyond covariates alone, and this incremental performance was sustained over multiple covariate specifications and evaluation metrics including AUPR (Supplementary Figs. 30–31; Supplementary Table 18; Supplementary Data 7). PES scores were weakly correlated with one another (median *|r|* = 0.17) and were built from largely distinct proteins (Supplementary Figs. 32–33).

PES tracked longitudinal exposure change. In participants with repeat proteomic profiling, within-person changes in PES correlated with changes in self-reported exposures (Fig. 6c). This tracking reflected how participants varied in their exposures across assessment visits (Supplementary Fig. 34). Alcohol PES increased with alcohol dose (*r* = 0.40), and smoking PES captured reversible exposure dynamics, increasing among individuals who started smoking and decreasing among individuals who quit (*r* = 0.69). Tracking held between two visits, from baseline to imaging (10-year difference) and between the two repeat imaging visits (2-year difference), where the score followed within-person change for the majority of exposures (Supplementary Fig. 35; Supplementary Data 7).

Finally, PES captured disease-relevant information. Scores that read and tracked their exposure tended to improve prediction for fewer, more mechanistically related diseases (Fig. 6d). Adding the PES improved held-out prediction for exposure–disease pairs including smoking and COPD and physical activity and type 2 diabetes (Supplementary Table 19; Supplementary Data 7). Together, these results show that the reporter component of the exposure-responsive proteome can serve as a broad longitudinal biomarker of modifiable lifestyle biology.

## Discussion

These findings support a view of the plasma proteome as a molecular record of the cumulative lifestyle exposome. Across more than 50,000 UK Biobank participants, 169 exposures, and 2,686 circulating proteins, HEAP identified coordinated proteomic signatures of modifiable lifestyle that were linked to disease risk, intervention response, and longitudinal change.

Prior studies have cataloged genetic and non-genetic determinants of circulating proteins^9,28,29^. Here, we identify exposure-responsive proteins by their reproducibility across independent modeling frameworks. HEAP and GREML represent genetic and exposomic effects differently, yet converged on overlapping sets of exposure-responsive proteins. Across the proteome, covariates (e.g., age, sex, body mass index) and genetics explain most protein variation, leaving a smaller but highly reproducible component for the exposome. Notably, a substantial exposure-associated component was detected for 608 proteins.

In contrast, polygenic gene-by-environment effects were small and did not reproduce out-of-fold. These findings suggest that the exposure-responsive protein variation detectable at this sample size reflects additive exposure-associated effects rather than interactions between genotype and lifestyle exposure. However, GŒE can manifest through other forms of genetic heterogeneity across environmental contexts, including shifts in genetic correlation or genetic variance across exposure strata^30^.

Prior work has shown that plasma protein signatures can predict disease risk and health states, with a few proteins having causal effects on disease^6,31^. HEAP extends this perspective by evaluating how exposure-responsive proteins should be interpreted in the context of lifestyle–disease relationships. In mediation analyses, 1,448 proteins showed statistically mediated exposure–disease associations involving at least one incident disease, suggesting that lifestyle-associated proteins often lie along disease-relevant pathways. Genetic analyses, however, supported far fewer: only eight proteins had cis-anchored, colocalized support for a causal effect on disease, and only three of those showed evidence of causal mediation. The genetic evidence instead ran predominantly in the reverse direction, with Tier-1 support far more common for disease liability acting on protein levels than for proteins mediating exposure into disease. Most exposure-responsive proteins are therefore markers of exposure burden, residual confounding, disease liability, or downstream disease-associated processes rather than causal intermediates. As such, proteins without strong evidence for causal mediation may be less useful as therapeutic targets, but they can still be valuable molecular biomarker readouts of lifestyle-associated physiology, early disease processes, and modifiable biological state.

Two lines of evidence support this interpretation. First, proteomic signatures associated with lifestyle exposures in the UK Biobank were concordant with protein changes observed after structured exercise training in HERITAGE^24^ and GLP1 receptor agonist treatment^25–27^. These comparisons reveal that part of the observational lifestyle protein signature overlaps with molecular programs that shift after behavioral or pharmacological perturbation. Within that shared set, the causal minority is small but tractable: PCSK9 and ASGR1 were the only proteins supported by Mendelian randomization, colocalization, and replication across both pQTL platforms, and both are established or putative druggable targets^32,33^.

Second, proteome-based exposure scores (PES) show that protein signatures can be used as molecular readouts of exposure biology. Although prior studies have built protein scores for individual traits such as smoking^34,35^ or body mass index ^36^, HEAP extends this approach across the lifestyle exposome. These scores captured baseline exposure status, tracked within-person lifestyle change, and improved prediction for selected exposure-related diseases. They offer an objective, repeatable measure of whether a modifiable exposure has actually shifted. Plasma proteins therefore provide molecular measures of lifestyle-associated physiology that complement self-reported exposures and may prove useful for prevention research, where verifying adherence and detecting biological response before disease onset are the central measurement problems, and where measurement error of this kind has been proposed as a source of bias in the lifestyle literature^37^.

Several limitations should be considered. Many lifestyle exposures in the UK Biobank are self-reported, and most HEAP analyses used baseline exposure and proteomic measurements, limiting inference about exposure history, timing, and directionality. These limitations particularly affect the observational exposure–protein associations and mediation analyses. Statistical mediation assumes the specified temporal ordering and absence of unmeasured confounding among the exposure, protein mediator, and disease outcome ^19,20^; therefore, residual confounding, measurement error, and collider bias may influence the estimated mediated effects despite covariate adjustment and sensitivity analyses.

The genetic and exposomic components estimated by HEAP should not be interpreted as fully separable biological effects. Lifestyle behaviors can themselves be partially genetically influenced, creating indirect genetic effects on proteins that operate through measured exposures. Although the heritability of the exposures analyzed here was generally small to modest (*h*^2^: 0–0.13, Supplementary Fig. 22), such pathways may contribute to variation attributed to the exposomic component. Moreover, HEAP models genetic and exposure-associated effects primarily as additive and linear. Nonlinear genetic and exposomic effects or more complex genotype-by-environment relationships may therefore not be captured.

MR provided a genetically anchored test of causal mediation, but is limited by exposure coding, heritability, instrument strength, pleiotropy, and differences between Olink and SomaScan pQTL resources. Consequently, power to identify MR-supported exposure–protein–disease triads varies across exposures, which may partly influence which exposures are represented among supported pathways. Additionally, complete exposure–protein–disease triads are constrained by the sparsity of genetically supported protein–disease relationships in current pQTL and disease GWAS resources^38^.

The two protein assay platforms (SomaScan and Olink) correspond only modestly for many proteins^39,40^. Cross-platform analyses were therefore restricted to overlapping proteins, weighted by reported assay concordance, and while we used pQTL resources from both platforms when possible, platform-specific effects may remain.

UK Biobank has a low participation rate, and participants are healthier, more socioeconomically advantaged, and predominantly of European ancestry relative to the general population, which may introduce selection bias and limit generalizability ^41^. Finally, our definition of the exposome is limited to lifestyle, physical-environment, and socioeconomic features available in the UK Biobank and does not capture many chemical, occupational, microbial, and social exposures that may also shape protein expression, although new emerging biobank-scale data may soon be available to deploy these approaches^3^.

Together, HEAP defines a lifestyle-centered view of the plasma proteome. Rather than treating exposure-associated proteins only as biomarkers or candidate causal targets, this framework organizes them by their reproducibility, disease relevance, genetic support, intervention concordance, and longitudinal behavior. The resulting architecture provides a resource for studying how modifiable exposures are represented in the plasma proteome and how those representations relate to disease risk.

## Methods

### Datasets

#### UK Biobank Cohort

The UK Biobank (UKB)^42^ contains 500k individuals with genetic, exposomic, and omics data. Individuals range from 40-70 years of age with genetic, health-record, exposomic, imaging, clinical, metabolomic, and proteomic measurements. We used UKB plasma proteomics for this study. Approximately 50k individuals had measurements across 3000 protein biomarkers that ranged across disease phenotypes: cardiometabolic, neurological, inflammation-related, and oncology-related ^42^. Normalized protein expression (NPX) values were obtained from UKB which were built from selected quality control measures and normalization procedures that accounted for batches^9^. We used exposomic measurements from the UKB from individual-level survey data. The measurements include lifestyle factors such as alcohol consumption, diet, exercise, smoking usage, internet usage, and vitamins. We incorporated neighborhood-level data of income and multiple deprivation indices which aggregate local income, employment, education, and housing. We used imputed genotypes from UKB as previously described ^43^ to generate genetic scores.

#### HERITAGE Cohort

We used plasma proteomic profiling from the HERITAGE cohort, which included 654 individuals of both European and African ancestry, to correlate findings with UKB exposure-protein associations ^24^. The HERITAGE cohort is a longitudinal intervention study, in which participants completed a 20-week endurance training program, exercising three times per week under supervision ^23^. Inclusion criteria required participants to be healthy (i.e. no cardiometabolic disease) and sedentary for three months prior to the exercise intervention. The study profiled 4,914 proteins using the SOMAscan platform and collected data at two time points: one before the program began and one 24 hours after the program ended^24^. These measurements were recorded at a fasted and rested state. We utilized the summarized statistics of relative changes in protein concentration from before to after the exercise regimen.

#### GLP1 Receptor Agonist Randomized Control Trials: STEP1 and STEP2 Cohorts

We used serum proteomic profiling from the STEP1 and STEP2 phase 3 randomized, double blind, placebo controlled clinical trials ^27^. STEP1 included participants with obesity, while STEP2 included participants with type 2 diabetes ^25,26^. Both trials had participants take 2.4 mg of semaglutide, a GLP1 receptor agonist, for 68 weeks to assess the safety and utility of semaglutide for weight loss and glycemic control in these respective populations. The fasting serum samples of 1956 individuals were profiled for approximately 6400 proteins using the SomaScan platform at baseline and after treatment^27^. Summarized statistics of proteomic effects were utilized to capture the relative change in protein concentration from baseline to week 68 with semaglutide 2.4 mg treatment compared to placebo.

### Preprocessing

#### Exposomic Features

We preprocessed 169 exposomic features (88 binary, 50 continuous, 31 ordinal) across the 13 categories: diet, exercise frequency, metabolic equivalent task (MET), vitamins, smoking, internet usage, sleep, alcohol usage, sexual history, sun exposure, air pollution, noise pollution, and socioeconomic deprivation. These features included a mixture of continuous, ordinal, unordered categorical, and binary features from survey questionnaire data. Ordinals are handled with contrasts, while unordered categorical and binary features are one-hot (dummy) encoded. Exposomic measurements with at least 80% complete cases were considered for the analysis, and any participants with missing data were excluded (Supplementary Table 20).

#### Plasma Proteomics

We utilized normalized protein expression accounting for batch effects and technical factors. For full details on quality control measures and repeatability of plasma protein expression, refer to Sun et al., 2023.

For the analysis involving predictive *R*^2^ decomposition, exposomic and gene-by-environment associations, and mediation analysis, we excluded proteins that did not have a built genetic score and utilized complete cases. For variance decomposition through GREML, we utilized all 2,923 proteins with complete cases. For proteome-based exposure scores (PES), we excluded proteins with greater than 20% missing and median imputed using the training set.

#### UK Biobank Disease Phenotyping

We utilized first-occurrence records with ICD-10 disease codes defined in UKB to assign disease outcomes in our analysis^44^. We defined prevalent disease to be any ICD-10 code time marked before or at baseline assessment. Incident disease is defined as an ICD-10 code marked after baseline assessment. To ensure stable estimates, we restricted analyses to diseases with at least 100 incident cases. 181 ICD-10 disease codes met this threshold when considering overlap with samples containing proteomics, covariate, and exposure data (Supplementary Table 21). These disease codes were used in both mediation and proteome-based exposure-score analyses. Additionally, for sensitivity analyses we constructed an exclusion set of individuals based on prevalent disease, in the following eight major chronic-disease categories: diabetes (E10–E14), ischaemic heart disease (I20–I25), heart failure (I50), stroke/cerebrovascular disease (I60–I64), COPD (J43–J44), chronic kidney disease (N18), liver cirrhosis (K70, K74), and dementia (F00, F01, F03, G30).

#### Covariates

We utilized a base covariate specification throughout all analyses for consistency: age+ sex +age^2^ +age*×*sex +age^2^*×*sex + assessment center + 20 genetic PCs. This base specification accounts for biological factors, age and sex, that influence protein expression along with adjustments for population structure, using assessment center and 20 genetic PCs. For sensitivity, we added the following additions to the covariate set. BMI inclusion (+BMI), additional factors related to drawing of blood (+fasting time, season), features related to co-morbidities mentioned as the “clinical” set (+BMI, fasting, season, common medications such as cholesterol lowering drugs, insulin, blood pressure lowering drugs, and hormone replacement therapy), and exclusion of prevalent disease cases (*∼*15% of participants with proteomics). We excluded people with prevalent disease in the following eight major chronic-disease domains (as described in “UK Biobank Disease Phenotyping”). Additionally, a batch effect covariate was not included for this analysis since the UKB proteomics were normalized with batch correction ^9^.

#### Protein Polygenic Score Utilization

We used polygenic scores (PGS) for each plasma protein in the UKB. A comprehensive study of genetic scores across the proteome, metabolome, and transcriptome has been conducted across multiple cohorts^45^ These polygenic scores, defined by calculated weights, were stored in the OmicsPred ^45^ database which includes scores built for UKB participants of multi-ancestry. These polygenic scores were constructed using a Bayesian ridge regression on individual-level genotype data under a train-test split. The scores were validated on external cohorts, such as the INTERVAL^46^ cohort, which profiled the same proteins as the UKB. To partition local (cis) from distal (trans) genetic regulation, we partitioned a protein’s polygenic score into cis and trans components where variants within *±*1 Mb of the protein-coding gene were aggregated into cis genetic score, *G*_cis_, and the rest of the variants aggregated into a trans genetic score, *G*_trans_.

### Variance Decomposition

#### GREML

We partitioned protein abundance variance into genetic (G), environmental (E), and gene-by-environment (G*×*E) components by multi-kernel GCTA-GREML ^18^, representing each component as an *n × n* similarity kernel across individuals. The genetic kernel, G, was a genetic relatedness matrix built from LD-pruned, genotyped SNPs. To restrict inference to unrelated participants we retained a maximal set with no pairwise relatedness above 0.025 (excluding relatives closer than first cousins), keeping 44,514 individuals. The environmental kernel was *E* = *XX*^T^*/M*, where *X* is the matrix of 169 exposures, continuous exposures centered and scaled to unit variance, ordinal exposures one-hot encoded, and *M* is its number of encoded columns. The gene-by-environment kernel was the element-wise product of the genetic and environmental kernels, (G *×* E)*_ij_* = *G_ij_ E_ij_*. For each protein we jointly fit the three kernels in a single REML model, taking the variance explained by each as its share of total phenotypic variance, and adjusted for the base covariate set (see Covariates). Each protein was analyzed on its own complete-case sample, non-missing protein, genotype, covariates, and exposures, among unrelated participants. Of 2,918 proteins, 2,238 converged and were retained.

#### Polyexposure Score Creation

We built polyexposure scores (PXS) using a lasso model. For each normalized plasma protein *P*, we fit a single lasso-penalized regression jointly on the protein’s cis and trans genetic scores (*G*_cis_, *G*_trans_), all exposure features (*E*), their gene-by-exposure interactions, and the base covariates (*C*): *P ∼ C* + *G*_cis_ + *G*_trans_ + *E* + *G*_cis_:*E* + *G*_trans_:*E*. The model was evaluated under 5-fold nested cross-validation. An outer 5-fold split estimated held-out predictive *R*^2^, and within each training fold an inner 5-fold cross-validation selected the penalty *λ* minimizing cross-validated error. To prevent leakage, centering and scaling parameters were learned on each training fold and applied to its held-out fold. From non-zero lasso weights, *β*, we formed per-protein polyexposure score PXS = ∑*_i_ β_i_E_i_* and a gene-by-exposure interaction score 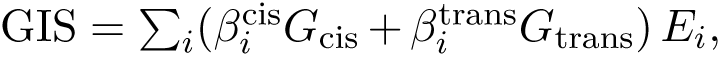 together with genetic scores from *G*_cis_ and *G*_trans_. Out-of-fold scores were constructed for all individuals, minimizing overfitting in downstream use of the PXS.

#### Predictive *R*^2^ decomposition

We partitioned each protein’s held-out predictive *R*^2^ into contributions from genetics (*G*), exposures (*E*), gene-by-environment (*G×E*), and covariates (*C*) by a leave-one-out drop-decomposition on the learned scores. Predicting *P* from the composite scores, each component’s unique contribution was the reduction in held-out *R*^2^ when its score was removed from the full model (e.g. unique *E* = *R*^2^(*C*+*G*+*E*+*G×E*) *− R*^2^(*C*+*G*+*G×E*)), averaged over the outer folds. Because the genetic score was partitioned into cis and trans, we applied the same procedure to decompose *G* into cis-or trans-genetic contribution. We considered this held-out predictive *R*^2^ over the base covariate adjustment along with alternative covariate specifications mentioned in the Covariates section. For a per-category decomposition, we recomputed exposure scores to predefined categories (e.g. diet, exercise, smoking, alcohol), PXS*_c_* = ∑*_i_*_∈_*_c_ β_i_E_i_*, reusing the full-model lasso weights, and applied the same drop procedure to obtain each category’s unique *R*^2^.

#### Exposomic and Polygenic GxE Associations

We assessed the effects of individual exposures via univariate E and polygenic GxE association tests to obtain effect sizes and significance of individual exposures. We constructed univariate associations under a 80/20 train-test split using a linear model for each protein: *P ∼ C*_1:_*_k_* + *G*_cis_ + *G*_trans_ + *E_i_* + *G*_cis_:*E_i_* + *G*_trans_:*E_i_*. We report the standardized beta’s for *E_i_*, the effect of an exposure on a protein, and *G*_cis_:*E_i_*, *G*_trans_:*E_i_*, the GxE interaction effect. Their respective p-values were computed with a nested-model *F*-test comparing the full model against one with the relevant term block removed: the *E_i_*block for the exposure effect, and the joint *G*_cis_:*E_i_, G*_trans_:*E_i_* block for the GxE effect. For multiple-hypothesis testing, a Bonferroni correction was applied based on the number of preprocessed exposomic features and proteins tested (see above Methods, “Preprocessing Exposomic Features”). We declared an effect to be validated if we found an association with a significant p-value in both the train and test sets. We checked the stability of estimates by running models under several covariate specifications (see “Covariates”). We performed Spearman correlation tests of validated effects observed across the specifications. From this, we assessed the effect size, direction, significance, and pattern of exposure-protein associations.

### Mediation Analysis

#### Motivation

We quantified the extent to which proteins may mediate the relationships between exposure, genetics, and disease. We assumed the following temporality for mediation analysis where genetics and exposures impact circulating proteins leading to development of disease. The genetic component is defined since birth and the exposure component is defined by past self-reported events recorded in surveys. The proteins are measured at baseline and incident (future) disease is captured via ICD-10 codes following this protein measurement. Therefore, each protein may act as a mediator between the genetic and exposomic determinants of disease. We use mediation analysis to quantify the indirect and direct effects. Where indirect effects are relationships between an exposure/genetics and disease outcome that occur through the mediator, protein. While direct effects are relationships that link exposures/genetics to the disease outcome independent of the mediator, protein. Mediation analysis comes with assumptions of no confounding or unmeasured confounding which can distort effects captured. We utilized the base covariate set and utilized additional specifications for sensitivity reporting.

#### Framework

We utilized the genetic (PGS) and constructed polyexposure score (PXS) to capture aggregate genetic and exposomic components and obtain stable mediation estimates. Mediation analysis involved two regression models ^20^: a mediator model and outcome model. We used a linear regression to define the mediator model: *P ∼* PGS + PXS + *C*. The mediator model regressed the protein, P, onto predictors, PGS and PXS, and covariates, C. We used a cox proportional-hazards regression to capture the time-varying disease outcomes defined by: *D ∼ P* + PGS + PXS + *C*. This outcome model regressed the disease outcome, D, onto P, PGS, PXS, and C. To resolve contributions of genetics and exposures, we additionally fit partitioned models that split genetics *G_cis_*, *G_trans_* into cis and trans scores and exposures into predefined categories. We refit all models under several covariate specifications to test the stability of estimates against confounding. We analyzed only diseases with at least 100 incident cases and required at least 500 overlapping participants for reporting estimates.

#### G-computation: Estimation of Indirect and Direct Effect

We decomposed each predictor’s effect into direct and indirect components by g-computation ^19,20,47^. For each predictor (PGS, PXS) we predicted the protein expression from the mediator model under a one-standard-deviation shift in that predictor (0.5 SD below to 0.5 SD above its mean), with all other predictors held at their observed values. This predicted protein expression value is then propagated through the outcome model. The natural indirect effect (NIE; referred to as mediation strength) is the change in disease hazard produced by this shift acting through the protein, and the natural direct effect (NDE) is the change with the protein held fixed. Both estimates are expressed on the hazard-ratio scale. We obtained multiple-testing corrected p-values by Benjamini–Hochberg and confidence intervals for the NIE and NDE using the delta method^48^. We summarized the proportion mediated (PM) as 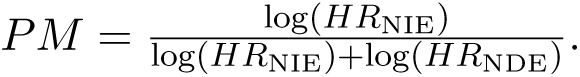

### Genetics and Heritability of Exposures

#### GWAS on exposures

We performed a genome-wide association study (GWAS) for each exposure using REGENIE^49^, under a two-step whole-genome regression procedure. The first step fit a whole-genome ridge model on directly genotyped, LD-pruned variants to capture polygenic background and sample relatedness. The second step tested imputed variants for association conditional on the first-step predictions. Quantitative exposures were rank-inverse-normal transformed, and binary exposures were analyzed on the binary scale with approximate Firth correction ^49^. To preserve a two-sample design for the downstream Mendelian randomization, the GWAS was run in UK Biobank participants without proteomic measurements (excluding the proteomics cohort), adjusting for the base covariates (see Covariates). We analyzed imputed variants with minor allele frequency *≥* 1%, genotype missingness *≤* 2%, and Hardy–Weinberg equilibrium *p ≥* 10^−5^.

#### Heritability of exposures

We estimated the SNP-based heritability of each exposure by LD score regression ^50^, using precomputed European-ancestry LD scores restricted to HapMap3 variants^51^. Heritability was reported for each exposure, and the regression intercept was retained as a model-based estimate of confounding from population structure, distinct from polygenicity.

### Mendelian Randomization

#### Framework and setup

We utilized Mendelian randomization (MR) to test causal relationships among the exposure–protein–disease triads found through mediation analysis (see Mediation Analysis) and exposomic associations (see Exposomic and Polygenic GxE Associations) of proteins. We formed these triads by matching each of the 6,683 Bonferroni-significant mediated protein–disease pairs to the replicated exposure–protein associations involving the same protein (17,505 of 22,240), giving 199,646 triads. Edges shared by multiple triads were estimated once, giving 77,573 directed MR tests. Genetic variants were utilized as instruments to estimate directed edges: exposure*→*protein, protein*→*disease, and exposure*→*disease. Along with testing reverse causation through edges: protein*→*exposure, disease*→*protein, disease*→*exposure. We estimated every edge in two arms. A split-sample UK Biobank arm, in which exposure instruments (from the non-proteomic GWAS section above) and UKB Olink pQTLs (measured only in the proteomics cohort) ^9^ are drawn from non-overlapping participants. We adopt this split sample MR approach, from the Deng et al., 2025 UKB proteomics analysis^6^. Additionally, we utilized an external replication arm using deCODE SomaScan pQTLs ^40,52^. We used two pQTL summary stats to account for potential differences in genetic variants tied to each platform, SomaScan vs Olink ^39,40^. Disease instruments were drawn from FinnGen (version Release 12) ^53^.

#### Obtaining instruments

For all genetic instruments we selected genome-wide significant variants (*p <* 5 *×* 10^−8^) with an instrument-strength F-statistic above 10, and LD-clumped them (*r*^2^ *<* 0.001 within a 10 Mb window) against a European reference panel ^22,51,54^. Protein instruments were defined separately as cis (within *±*1 Mb of the coding gene region) and trans (all remaining variants), and the two sets were tested as distinct instruments.

#### MR pipeline and sensitivity analyses

For each edge we harmonized the instrument–outcome data and selected the estimator by the number of available instruments: a Wald ratio for a single instrument, inverse-variance weighting (IVW) for two, and IVW, MR-Egger, the weighted median, and the weighted mode for three or more^55–58^. We then applied a fixed sequence of sensitivity checks: Cochran’s *Q* for heterogeneity, the MR-Egger intercept for directional pleiotropy, Steiger filtering for causal direction ^54^, and MR-PRESSO for outlier detection and correction ^59^. All MR estimators and tests were computed with the TwoSampleMR framework^60^. Edges were multiple-testing corrected by Benjamini–Hochberg FDR within each edge type (*q <* 0.05). We consolidated these results into a tiered confidence system (Table 1): an edge advanced from significance, through instrument robustness (no heterogeneity, pleiotropy, or rescue by MR-PRESSO or weighted median), to a confirmed causal direction (via steiger filtering), and for cis-pQTL edges, a colocalization test. Edges reaching the top tier in both arms were labeled cross-arm replicated Tier1+, but we report in the main analysis any MR hits that are Tier1 or above.

#### Tier system

Table 1 defines the MR confidence tiers. A null edge fails FDR, a suggestive edge is significant but has too few instruments or unresolved heterogeneity or pleiotropy. A reverse edge is robust but points against the hypothesized direction (the tested direction). Tier 2 collects robust, correctly directed edges whose pQTL instrument is trans only or the cis signal fails to colocalize. Tier 1 is the high-confidence set (including colocalized cis-pQTL edges). Tier 1+ marks Tier 1 edges replicated across both the UK Biobank and deCODE arms.

#### Colocalization

For cis-pQTL edges (protein*→*disease and protein*→*exposure), we assessed whether the protein and outcome signals share a causal variant using Bayesian colocalization ^61^. Within *±*500 kb of the cis lead variant, we computed the posterior probability of a single shared causal variant (PP.H4) under the method’s default priors, requiring at least 50 shared variants per locus. An edge was declared colocalized at PP.H4 *≥* 0.8. This threshold served as a hard gate on the tier system. A cis-pQTL edge that failed to support colocalization was demoted to Tier 2 and was not interpreted as evidence of a shared causal variant. Colocalization was applied only to cis-pQTL edges, where the single-shared-variant assumption is appropriate.

### Observational and Interventional Cohort Comparison

We compared the effect sizes of exposure-protein associations with circulating proteomics of intervention studies. Exercise intervention study HERITAGE^23,24^ and GLP1 agonist randomized control trials (RCTs) STEP1 and STEP2^25–27^. For the HERITAGE study, reported log fold change (logFC) differences between pre-vs post-training proteomic signatures were used for comparison with UKB effect sizes. Approximately 250 plasma proteins were able to be cross-referenced between the HERITAGE SomaScan and UKB Olink platforms. For the STEP1 and STEP2 RCTs, the summary statistics of the serum protein longitudinal effects from both the STEP1 and STEP2 trials were used for comparison with UKB effect sizes. Approximately 2.5k proteins were cross-referenced between the STEP1/2 SomaScan and UKB Olink platforms. We correlated the exposure-protein effect sizes found in UKB to effects observed in each intervention study. We calculated Pearson correlations and p-values adjusted for FDR using the Benjamini-Hochberg procedure for each intervention separately. We integrated correlations between SomaScan and Olink platforms for proteins described in Eldjarn et al., 2023^40^ to reference potential transferability. Additionally, MR evidence (Tier 1 and above) were integrated to assess which proteins were putatively causal for type 2 diabetes and its comorbidities: obesity, lipoprotein dysfunction, and hypertension.

### Proteome-Based Exposure Scores

We built proteome-based exposure scores (PES) that predict an individual’s exposures from their proteome and evaluated its ability to predict exposures that changed longitudinally and its prognostic value in predicting incident disease outcomes. We utilized participants with complete proteomic data and split participants based on if they had repeated followup. We trained and validated the PES based on 51,804 participants with only-baseline measurements and held-out test on 1,210 participants with repeated proteomic measurements at imaging and repeat-imaging visits in the UKB. We trained and tested these scores in this fashion, to make sure the held-out test set is never introduced during model training.

### Training and PES construction

We utilized the full set of approximately 2,900 proteins from UKB to predict each of the 169 exposures using lasso regression. We used median imputation for the proteomics utilizing the training samples from each fold. For model selection we used 5-fold nested cross-validation, the outer 5-fold loop produces out-of-fold predictions and the inner 5-fold cross-validation tuned the lambda penalty within each fold. The out-of-fold predictions constructed the per-participant PES for cohort, preventing overfitting when the score is used in downstream analysis for disease prediction. A final model, refit on the full training cohort, is applied to the held-out repeat-visit participants for evaluation. We trained models with only-proteomics, proteomics + covariates, and covariates only to assess the predictive performance of the constructed PES compared to covariates.

### Disease prediction with PES

To assess if the PES has prognostic value, we modeled incident disease with cox proportional-hazards regression, using the out-of-fold PES as a predictor. For each exposure–disease pair we compared these nested models: (i) covariates, (ii) covariates and self-reported exposure, and (iii) covariates, self-reported exposure, and PES. We report the hazard ratio (HR) of the PES along with its concordance (C-index). We fit these Cox models in the 51,804 baseline-only training participants using the out-of-fold PES, froze the resulting coefficients, and applied them to the 1,210 held-out repeat-visit participants. We report the incremental C-index in that held-out set, with out-of-bag bootstrap for variability. We ran the disease predictions covering 181 conditions with at least 100 incident cases, and corrected associations using the Benjamini–Hochberg FDR across all exposure-disease pairs.

### Evaluation Metrics

For exposure prediction, we reported the area under the ROC curve (AUC) and area under the precision–recall curve (AUPR) for binary exposures and the *R*^2^ for continuous exposures on the held-out test set at each visit. For temporal tracking of exposures via an individual’s PES, we computed the change in correlation, Δ correlation, between the within-person change in the observed self-reported exposure and the change in the predicted PES across the held-out test set in repeated visits. Disease prediction is summarized by the concordance (C-index) on bootstrapped held-out test set.

### Enrichment Analysis

#### Pathway enrichment

We applied enrichment analysis to identify pathways and tissues impacted by the lifestyle exposome. We used rank ordered significant exposure-protein association effect sizes (i.e. t-statistic) to find biological pathways and tissues upregulated or downregulated for each exposure (e.g. alcohol, diet, exercise, smoking, etc.). We used the gene set enrichment analysis, GSEA^62^, approach with parameters, minGGsize = 10, maxGGsize = 500, and FDR correction to obtain pathways and tissues significantly enriched (FDR < 0.05) in exposure-protein associations. We utilized resources GO ^63,64^ and Reactome ^65,66^ for pathway enrichment, and GTEx^67^ for tissue enrichment.

#### Tissue specific enrichment

We defined tissue-specific protein sets to tissue enrichment analysis using GTEx. GTEx is a resource measuring gene expression from 838 deceased donors with genotype information across 54 tissues, to assign plasma proteins to specific tissues ^67^. Using the gene expression defined by GTEx, we classified a gene in a tissue set if the fold change for a specific tissue is 4 times that of the mean expression of other tissues. This was determined through referencing ways plasma proteins are defined to organ sets^68,69^. We extended this analysis to retain tissue-specific results and to balance out organs that have multiple tissues, such as the brain which is divided into multiple brain regions.

## Data and Code Availability

Individual-level UK Biobank data are available to approved researchers through the UK Biobank Access Management System (application 52887) and cannot be redistributed by the authors. Curated summary tables are provided as Supplementary Tables (HEAP_Supplementary_Tables.xlsx), and large article-specific result tables are provided as Supplementary Data (HEAP_Supplementary_Data.zip). PES weights are released as an open resource in the Supplementary Data. These resources are also available through https://heap.bio, which provides access to materials and links associated with this study. Analysis code is available at https://github.com/shakson-isaac/HEAP.

## Data Availability

All data produced are available online at https://heap.bio.

## Acknowledgements

This work was supported by grants from the National Institutes of Environmental Health Sciences (NIEHS) R01ES032470, National Institute of Diabetes and Digestive and Kidney Diseases (NIDDK) R01DK137993. National Institutes of Environmental Health Sciences (NIEHS) U24ES036819, Advanced Research Projects Agency for Health ARPA-H D24AC00345.

## Author Contributions

S.I. developed the study concept and methodology, performed the computational and statistical analyses, interpreted the results, created the visualizations, and contributed to manuscript preparation and editing. R.J.E., V.L.M., and M.S.U. contributed to study design, statistical methodology, and interpretation of results. Y.H.J., A.G., S.S., B.M.N., and A.R.M. contributed to genetic and statistical methodology and interpretation of results. C.J.P. contributed to the study concept, methodology, supervision, and interpretation of results. All authors contributed to manuscript preparation and editing and approved the final manuscript.

