## Supplementary Document for "Human Plasma Proteomics Links Modifiable Lifestyle Exposome to Disease Risk"

#### Contents


**1 Supplementary Figures** **1**

**2 Supplementary Tables** **37**

**3 Supplementary Data** **38**


#### 1 Supplementary Figures

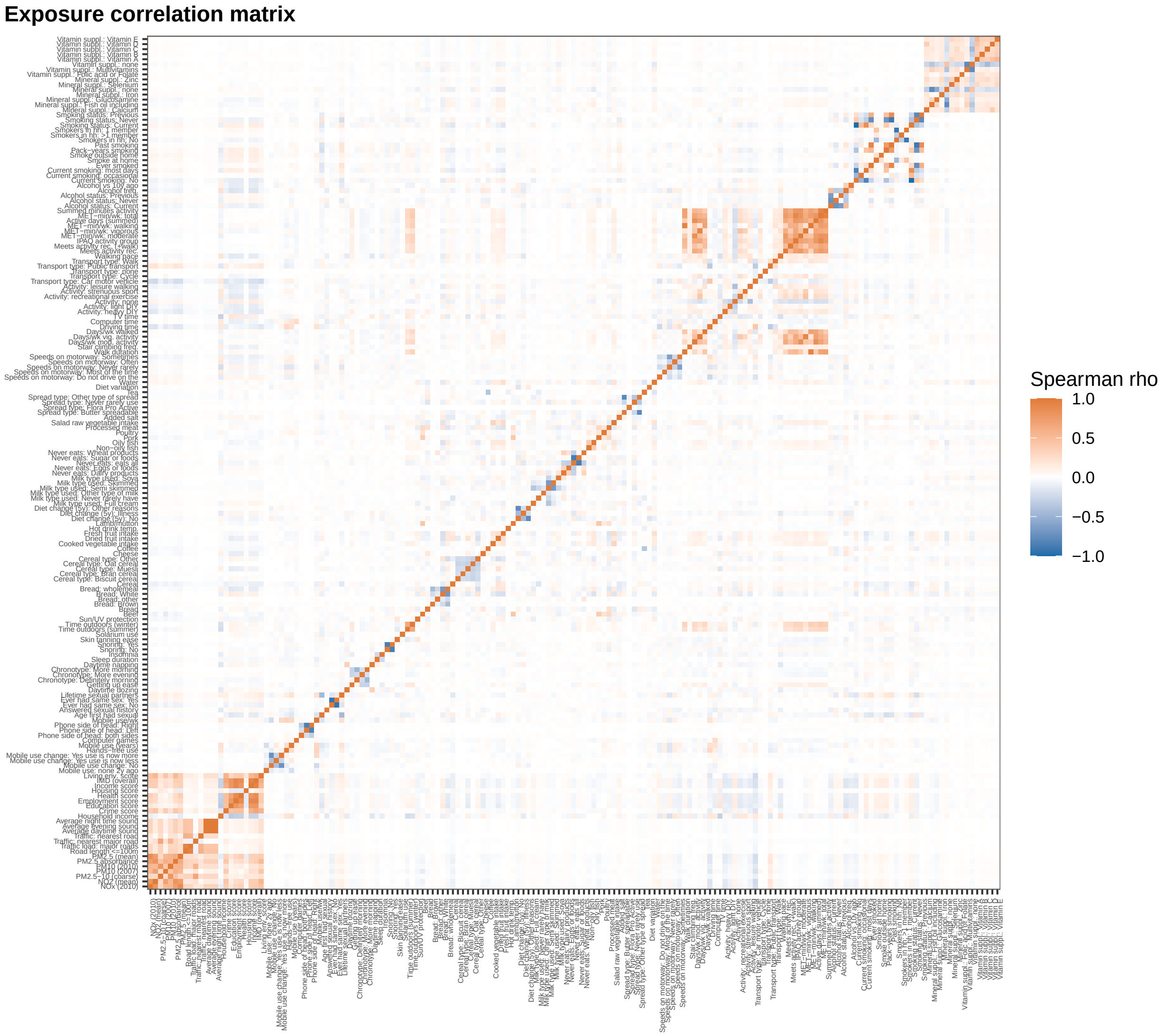


**Supplementary Figure 1: Correlation structure of the exposome.** Spearman rank correlations among the 169 exposure features across 502k participants.

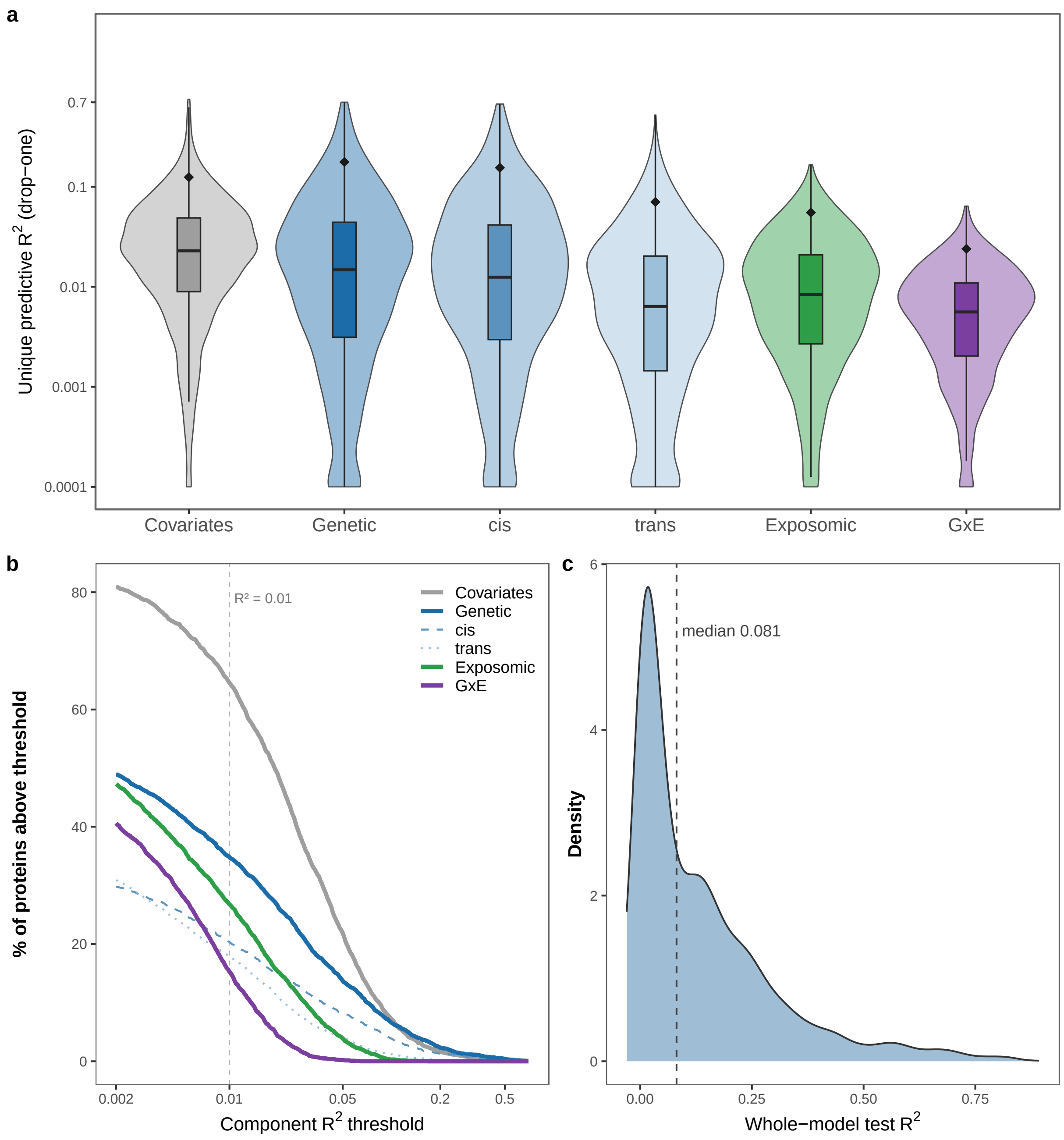


**Supplementary Figure 2: Per-protein variance architecture.** Decomposition of out-of-fold predictive $R^2$ across 2,686 proteins using base covariate set and lasso penalized regression. Every value is the unique (drop-one) out-of-fold $R^2$ of that component given the others; cis and trans subdivide the genetic component. **(a)** $R^2$ distribution by each component, showing only non-zero components. Median and IQR, with diamond showing the 95th percentile. **(b)** Number of proteins reaching over specific $R^2$ thresholds. $R^2 = 0.01$ line defines exposure-responsive proteins. **(c)** Whole-model out-of-fold $R^2$, all components combined.

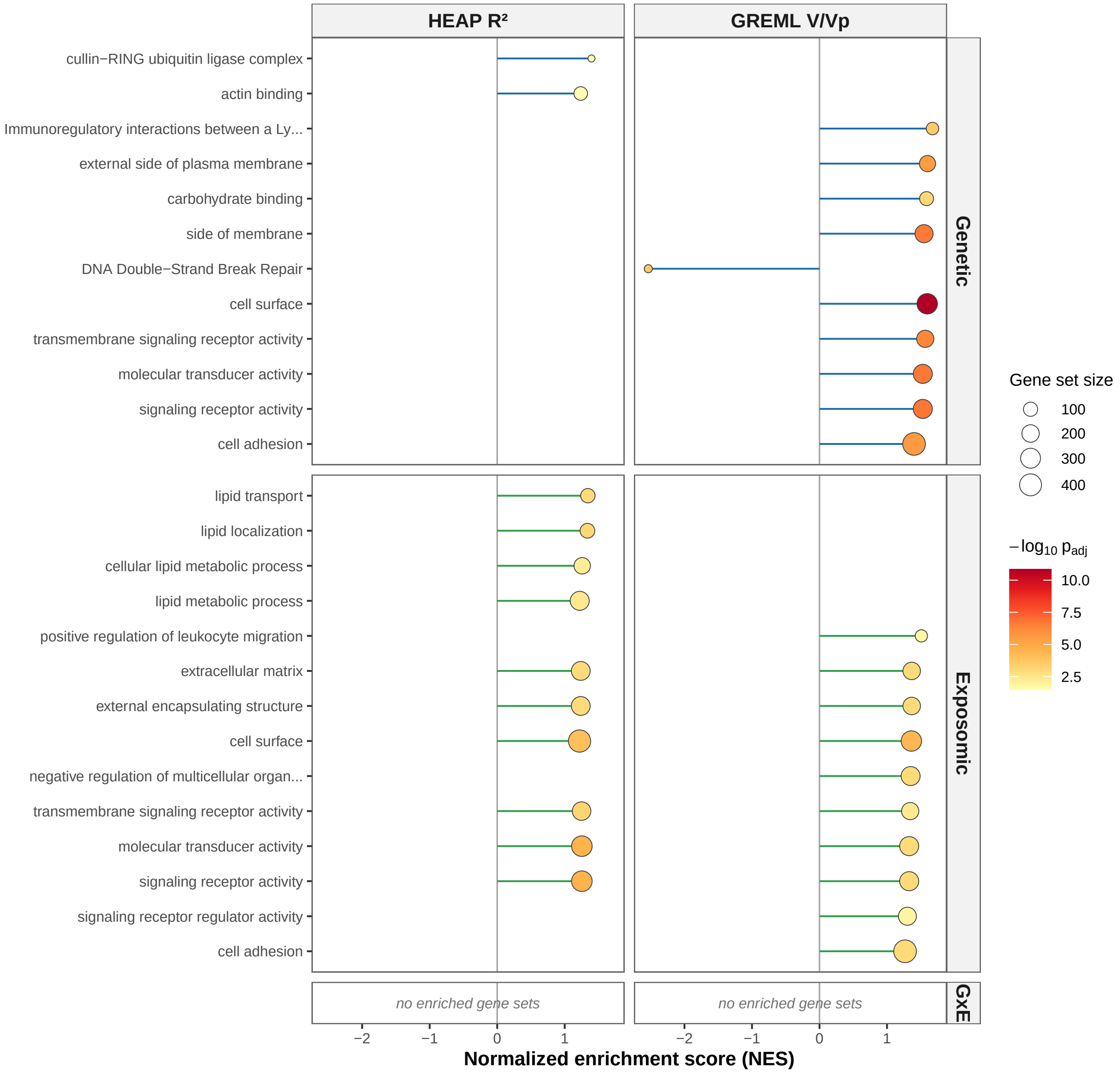


**Supplementary Figure 3: Gene-set enrichment of variance component methods.** GSEA (GO and Reactome; BH-FDR $q < 0.05$) of the 2,686 proteins, ranked two ways: by HEAP's unique (drop-one) out-of-fold $R^2$ and by the independent GREML variance component $V/V_p$. Top 10 enriched sets with the smallest adjusted $p$-value are shown.

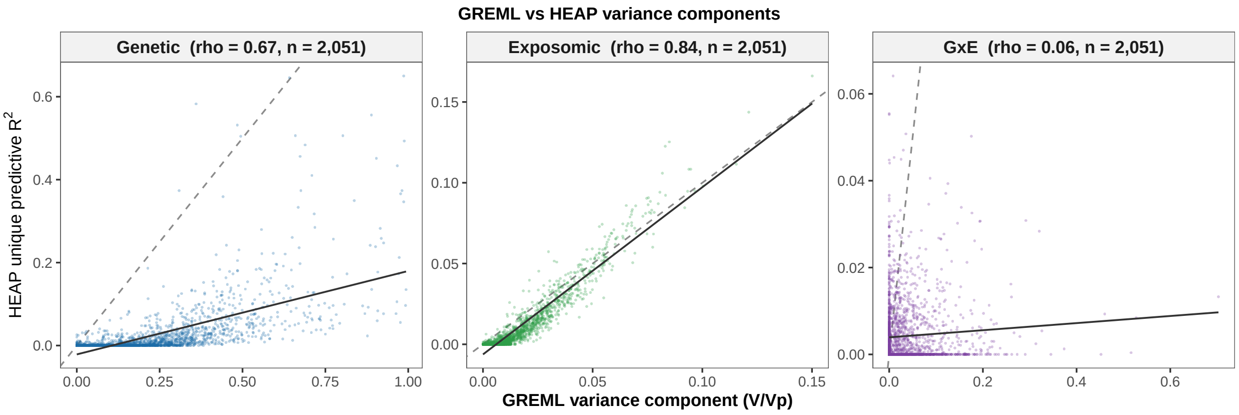


**Supplementary Figure 4: GREML versus HEAP variance components.** Concordance between the whole-genome GREML variance component ($V/V_p$) and the HEAP unique (drop-one) out-of-fold $R^2$, over the 2,051 proteins with a GREML estimate for all three components.

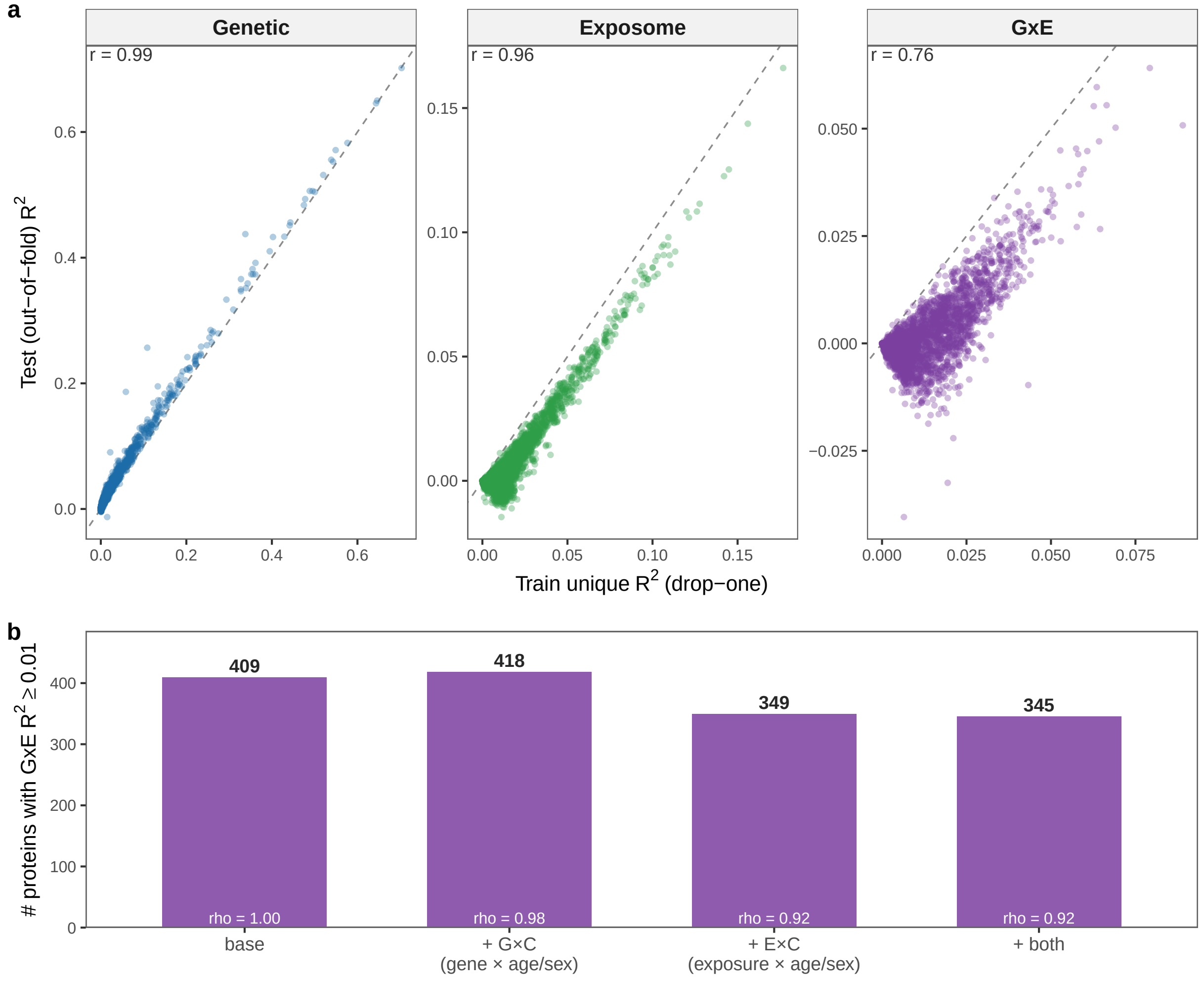


**Supplementary Figure 5: Stability of the gene-by-environment component. (a)** Training versus held-out unique (drop-one) $R^2$ per protein (2,686 proteins). **(b)** Proteins with GxE $R^2 \geq 0.01$ when re-fit with explicit age/sex interaction terms. Gene $\times$ covariate (G$\times$C), exposure $\times$ covariate (E$\times$C), or both compared to the base model; $\rho$ is the Spearman correlation of that model's per-protein GxE $R^2$ with the base model.

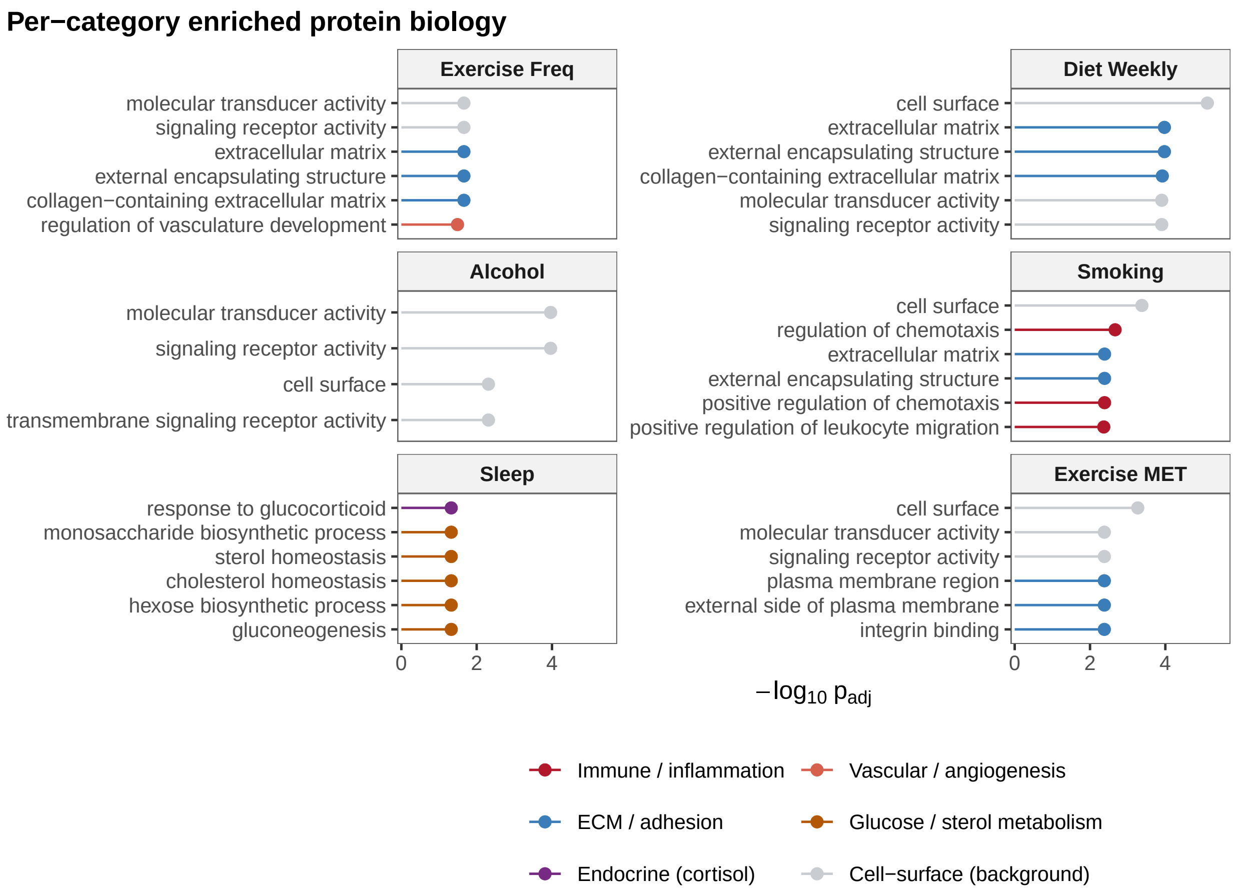


**Supplementary Figure 6: Enriched protein biology by exposure category.** GSEA (GO and Reactome; BH-FDR $q < 0.05$) of the 2,686 measured proteins, with the proteome re-ranked for each exposure category by that category's unique out-of-fold exposomic $R^2$. Shown for six lifestyle categories. Up to six positively enriched sets with the smallest adjusted $p$-value are shown.

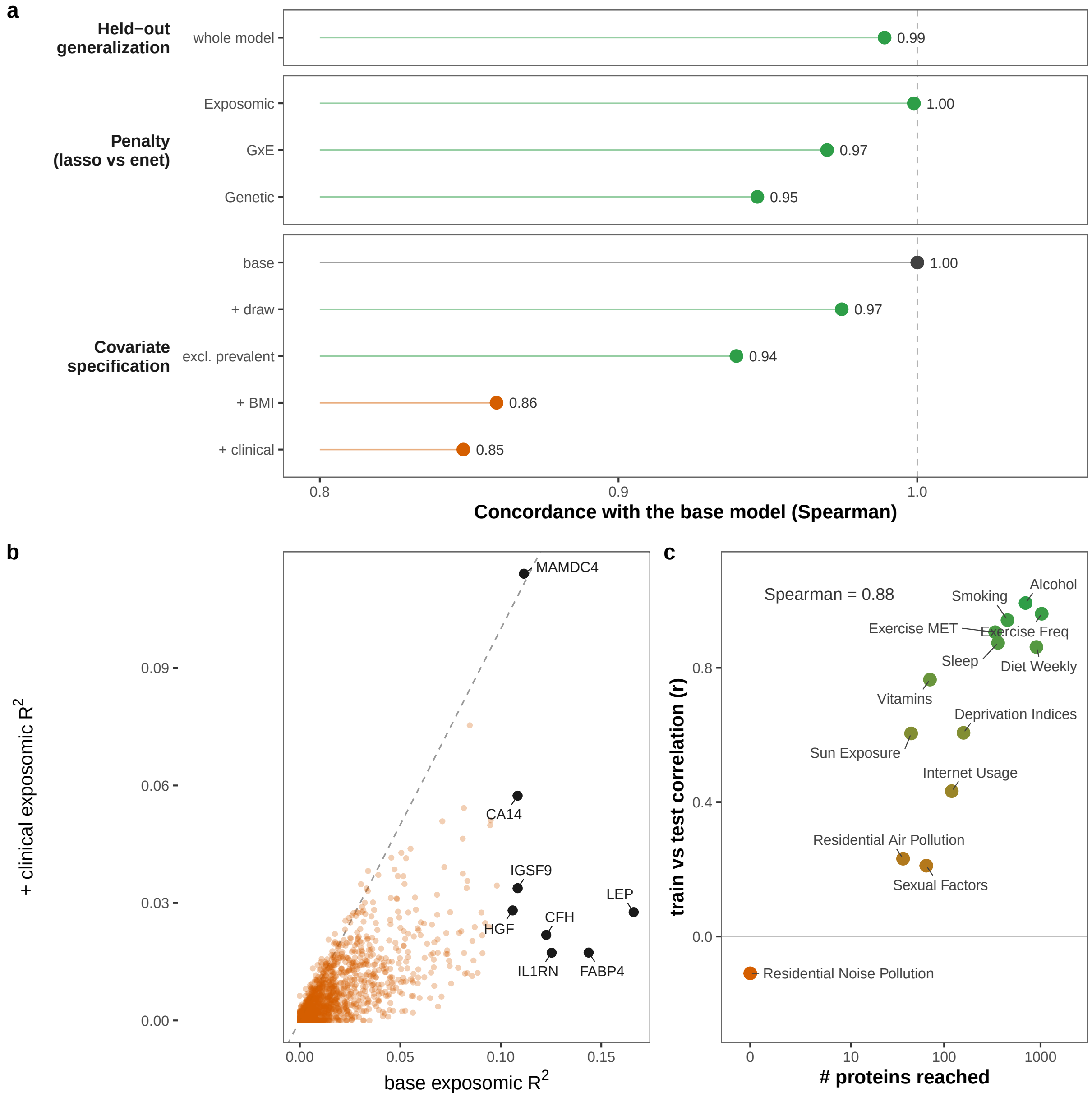


**Supplementary Figure 7: Robustness of HEAP estimates.** **(a)** Spearman concordance with the base model of the per-protein estimate under each perturbation. The held-out train/test split (whole-model $R^2$), lasso versus elastic-net penalty (shown per component), and five alternative covariate specifications (exposomic $R^2$). **(b)** Per-protein exposomic $R^2$, base versus clinical covariate specification. Eight adiposity-linked exemplars are labeled. **(c)** Per-category train versus held-out correlation of the per-protein category $R^2$, against number of proteins with $R^2 \geq 0.001$, for the 13 exposure categories.

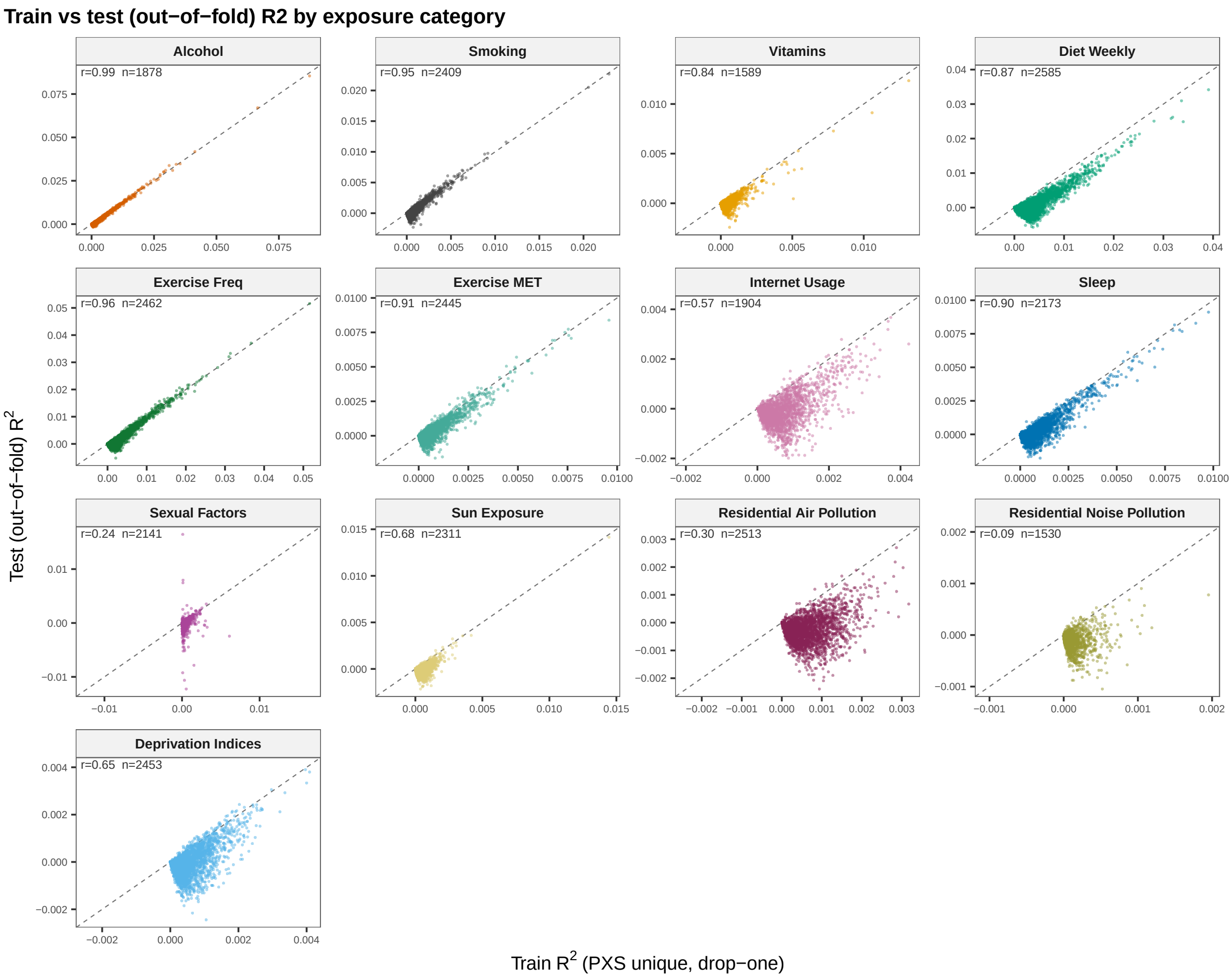


**Supplementary Figure 8: Train versus out-of-fold unique $R^2$, by exposure category.** Training versus held-out unique (drop-one) $R^2$ (base specification, lasso penalty) per exposure category.

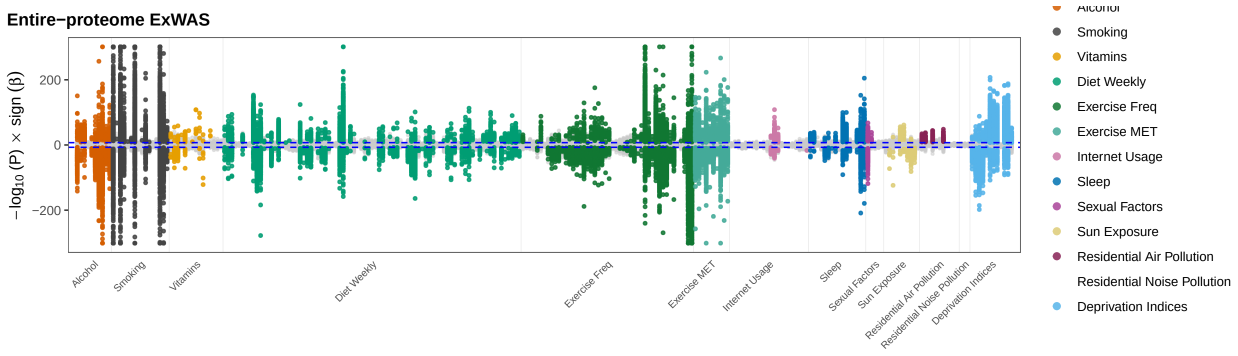


**Supplementary Figure 9: Signed exposome-wide (ExWAS) Miami plot.** One point per exposure term–protein pair, plotted as the training-split $-\log_{10} p$ signed by the direction of $\beta$. Colored pairs are Bonferroni-significant in both the training and the held-out split; gray pairs are not.

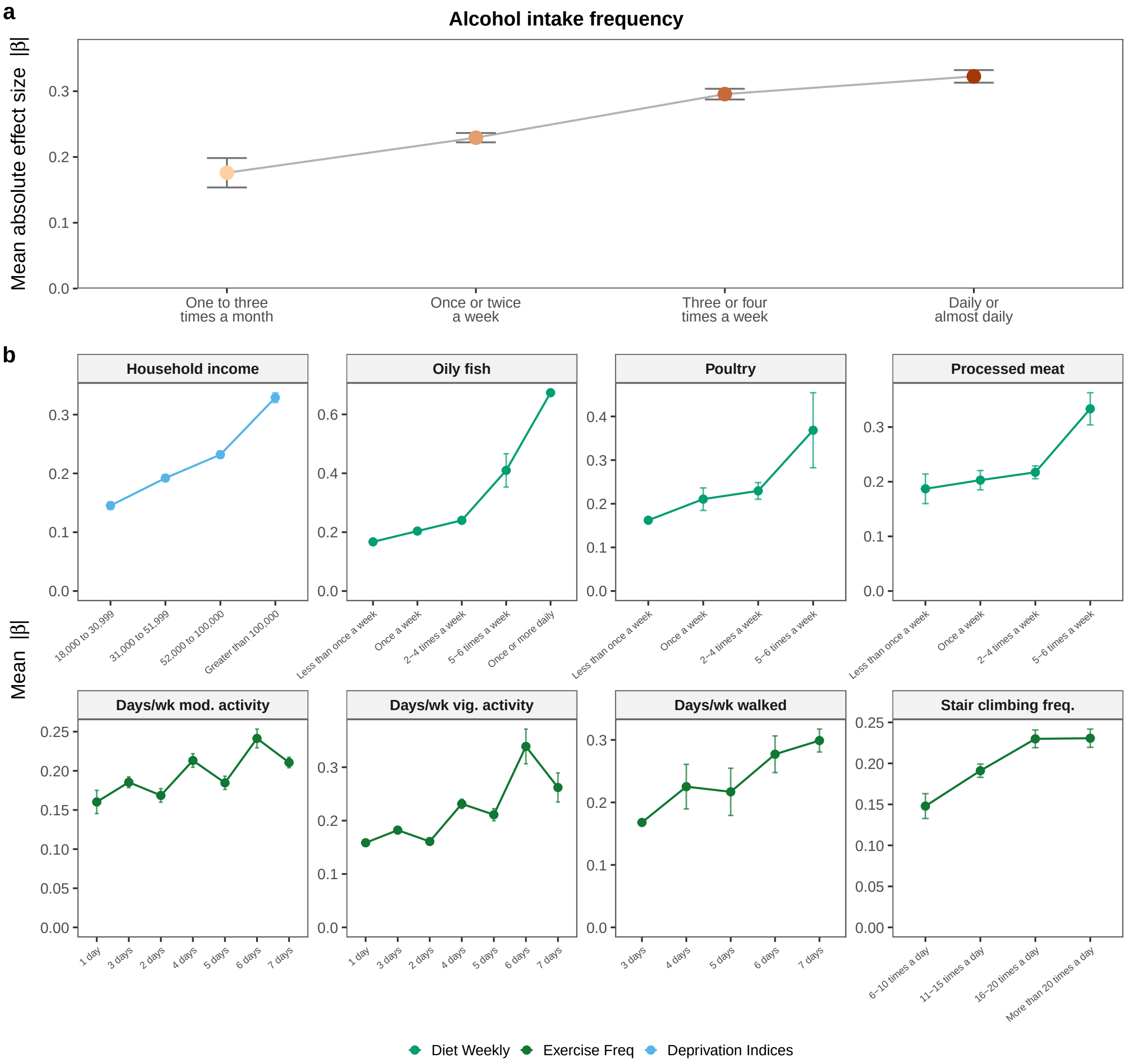


**Supplementary Figure 10: Dose-dependence of exposure–protein effect sizes.** Mean absolute effect size over every replicated protein term at each reported level of an exposure. Error bars are 95% confidence intervals on that mean, taken from the spread of $|\beta|$ across proteins. Levels are the reported response categories and are compared to the reference (lowest category). **(a)** Alcohol intake frequency. **(b)** Example exposures across diet, exercise, and deprivation indices categories.

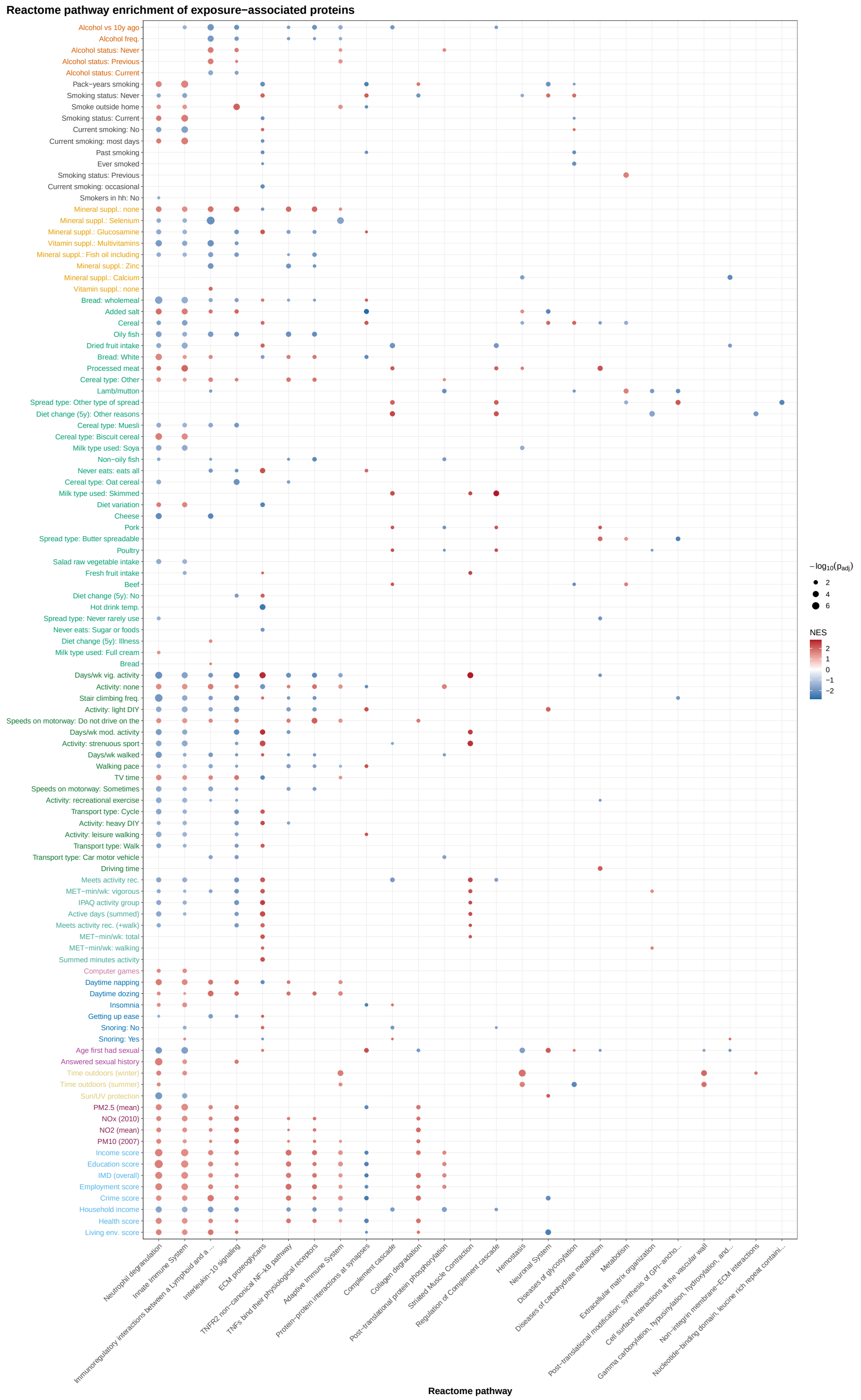


**Supplementary Figure 11: Reactome pathway enrichment per exposure.** GSEA of exposure–protein associations, ranked by association $t$ statistic, against Reactome gene sets; BH-FDR $q < 0.05$. Of all pathways reaching FDR significance, the 25 with the strongest peak significance across exposures are shown.

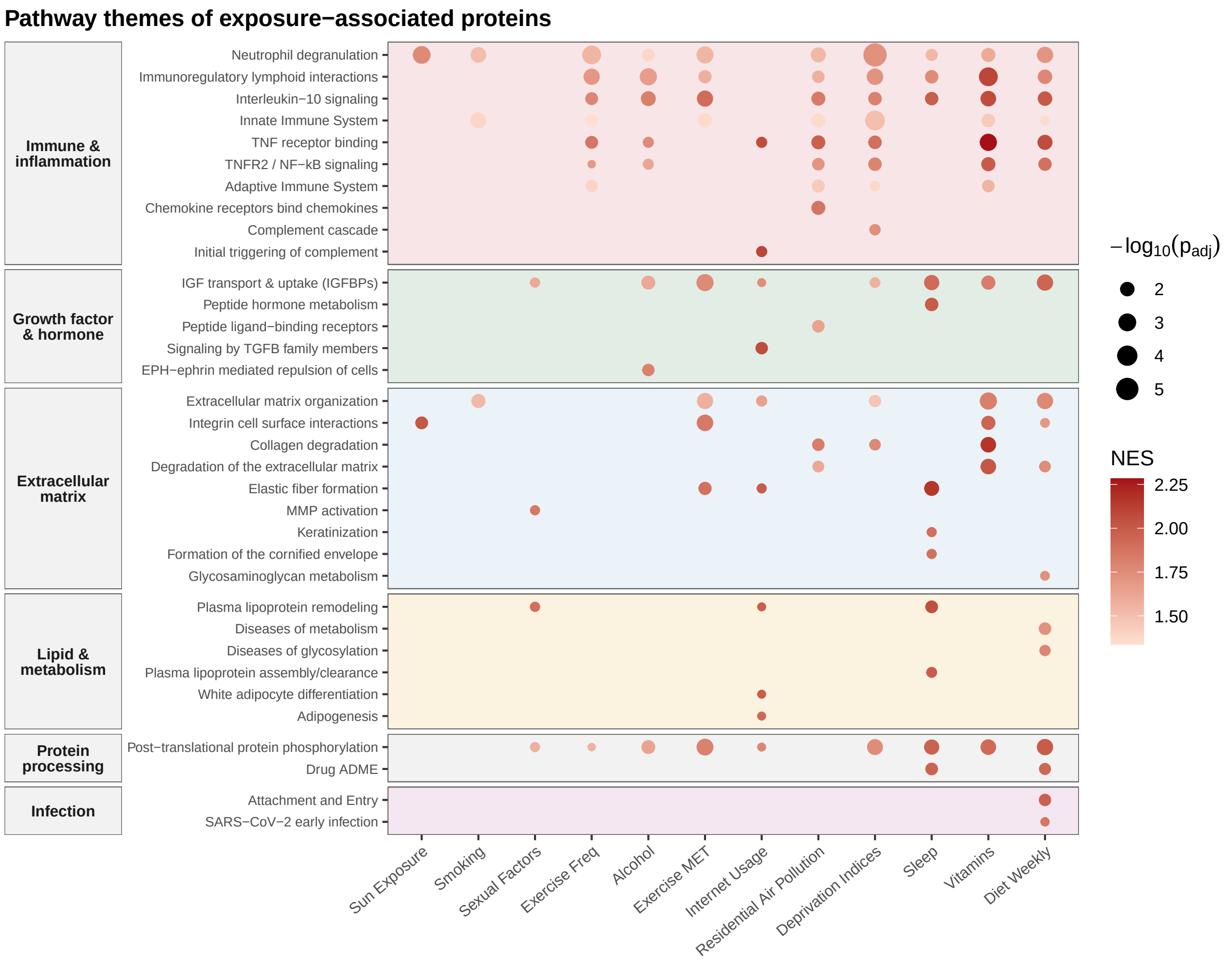


**Supplementary Figure 12: Reactome pathway enrichment per exposure category.** GSEA of the proteins associated with each exposure category, ranked by association $t$ statistic, against Reactome gene sets. Only positive enrichments surviving multiple-testing correction are drawn (NES $> 0$, BH-FDR $q < 0.05$).

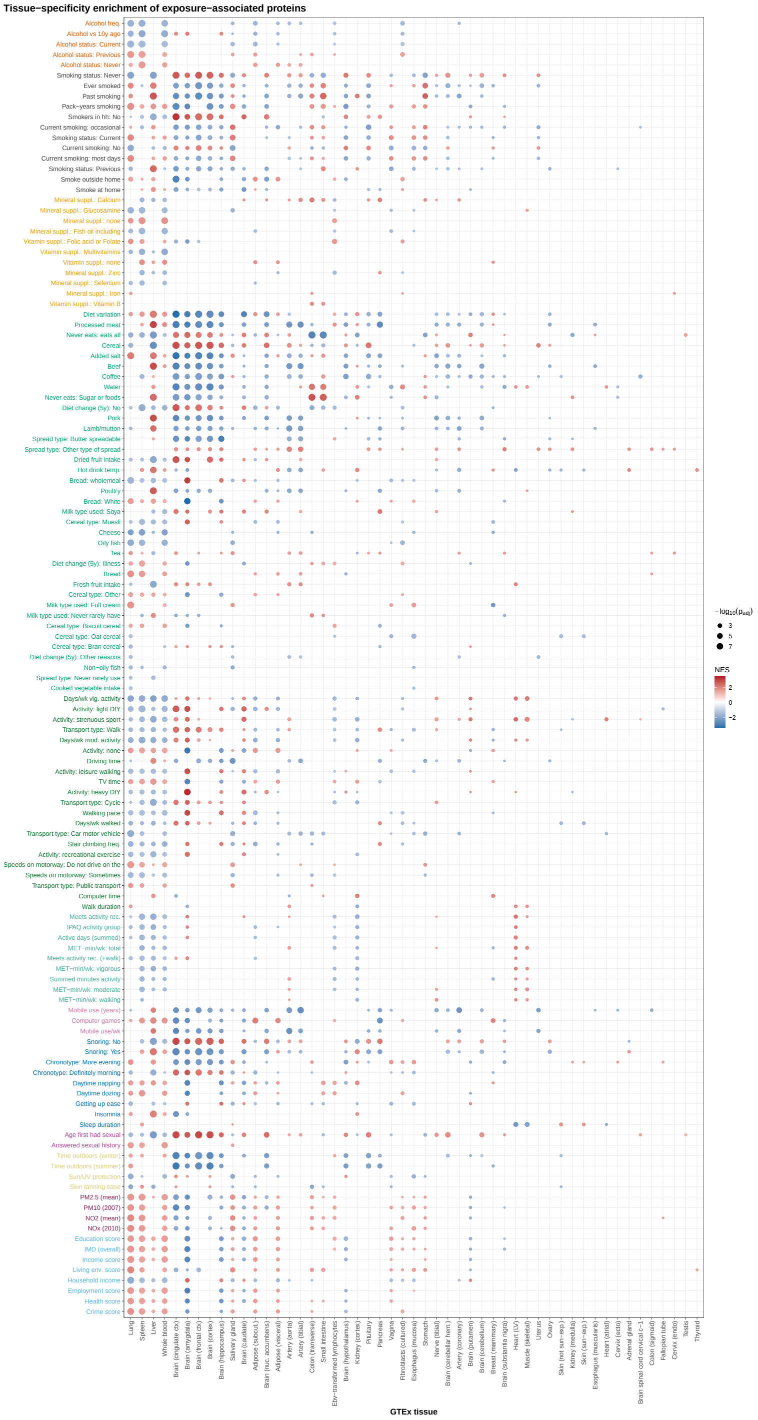

**Supplementary Figure 13: Tissue-specificity enrichment per exposure.** GSEA of exposure–protein associations, ranked by association $t$ statistic, against GTEx tissue-specific gene sets; BH-FDR $q < 0.05$. Every significant enrichment is shown.

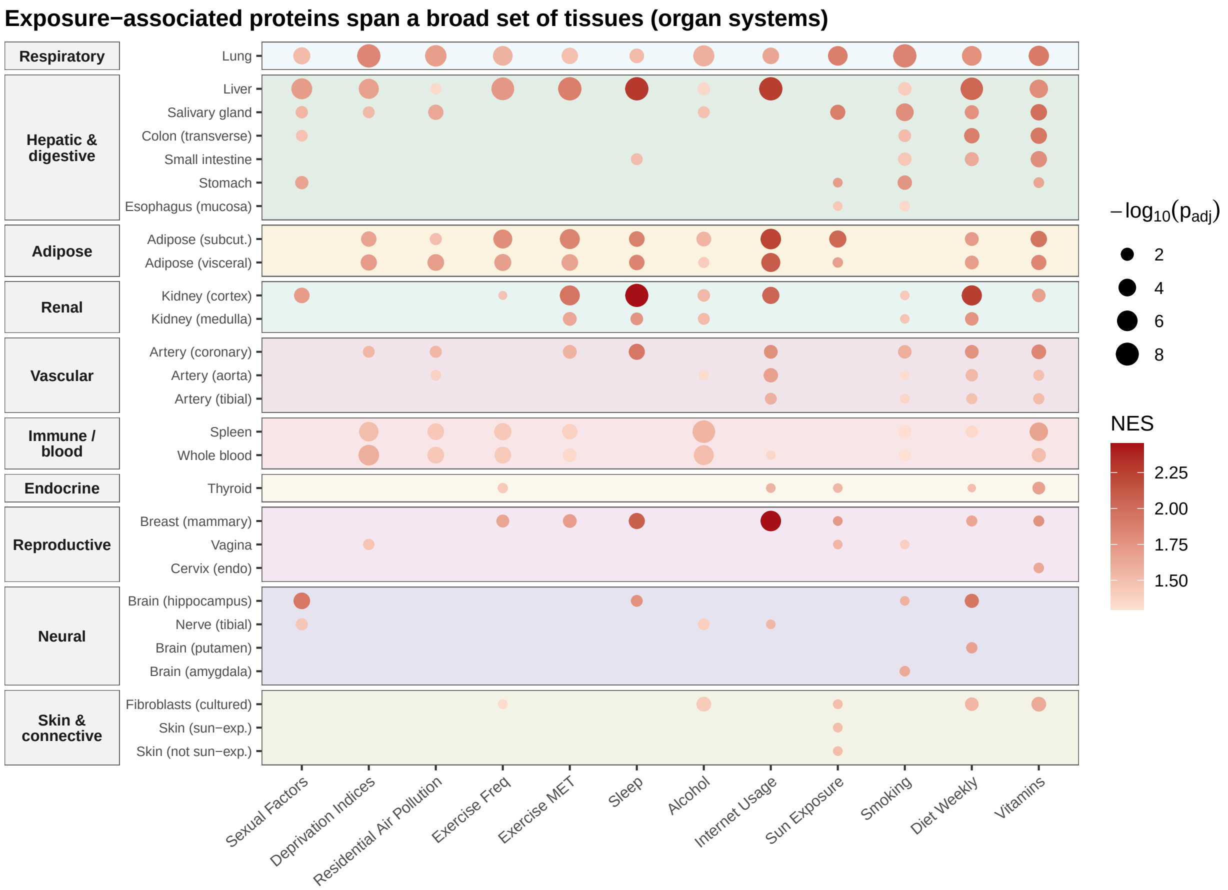


**Supplementary Figure 14: Tissue-specificity enrichment per exposure category.** GSEA of the proteins associated with each exposure category, ranked by association $t$ statistic, against GTEx tissue-specific gene sets. Only positive enrichments surviving multiple-testing correction are drawn (NES $> 0$, BH-FDR $q < 0.05$).

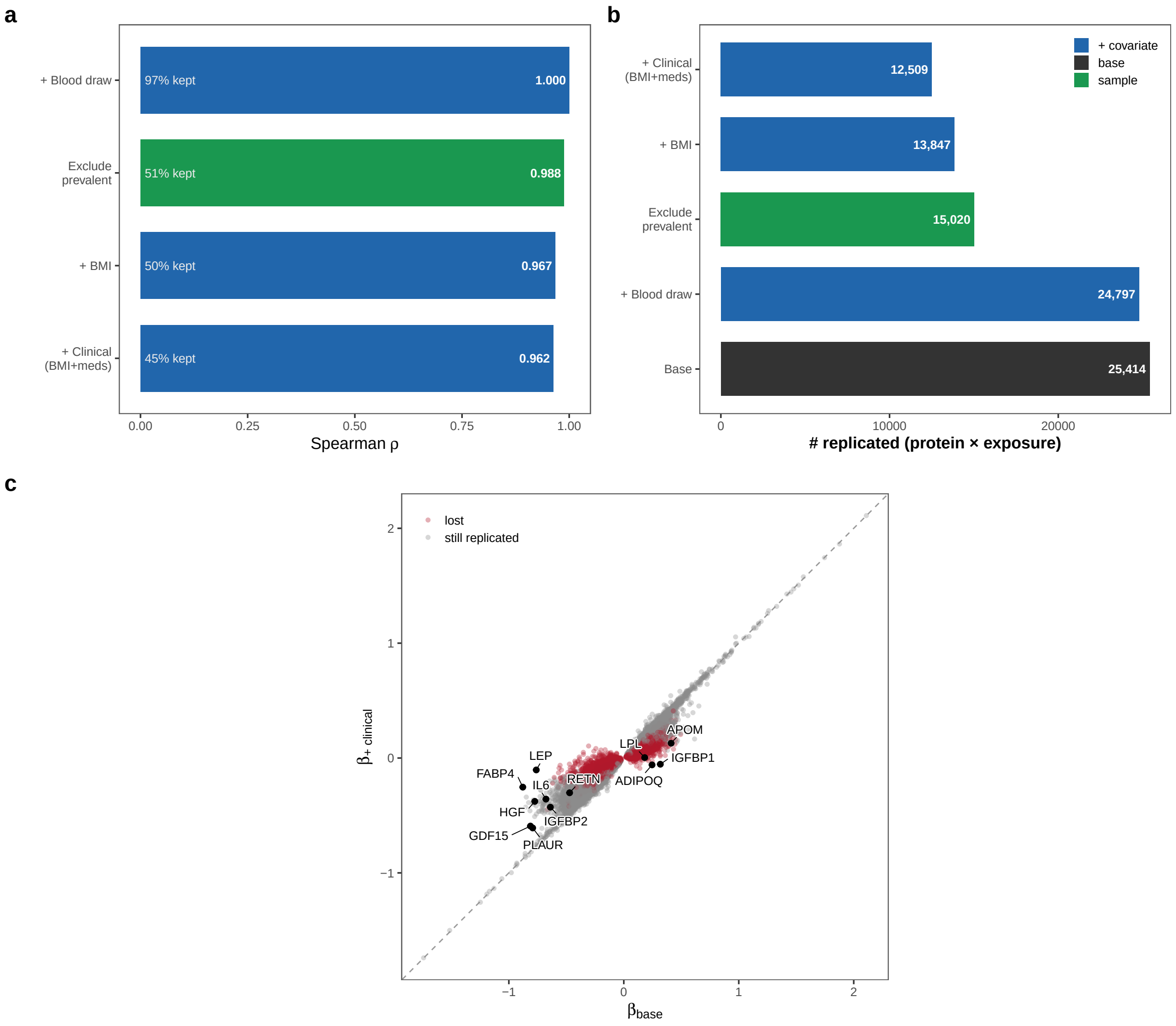


**Supplementary Figure 15: Covariate-specification sensitivity of exposure–protein associations.** Base-replicated pairs (Bonferroni-significant in both 80/20 splits) were re-estimated under each alternative specification. **(a)** Concordance with base specification effects, $\beta$. **(b)** Number of replicated pairs per specification. **(c)** Clinical (BMI + medication classes) versus base $\beta$. Black points show the twelve largest $|\beta|$ drops among adiposity- and inflammation-linked proteins.

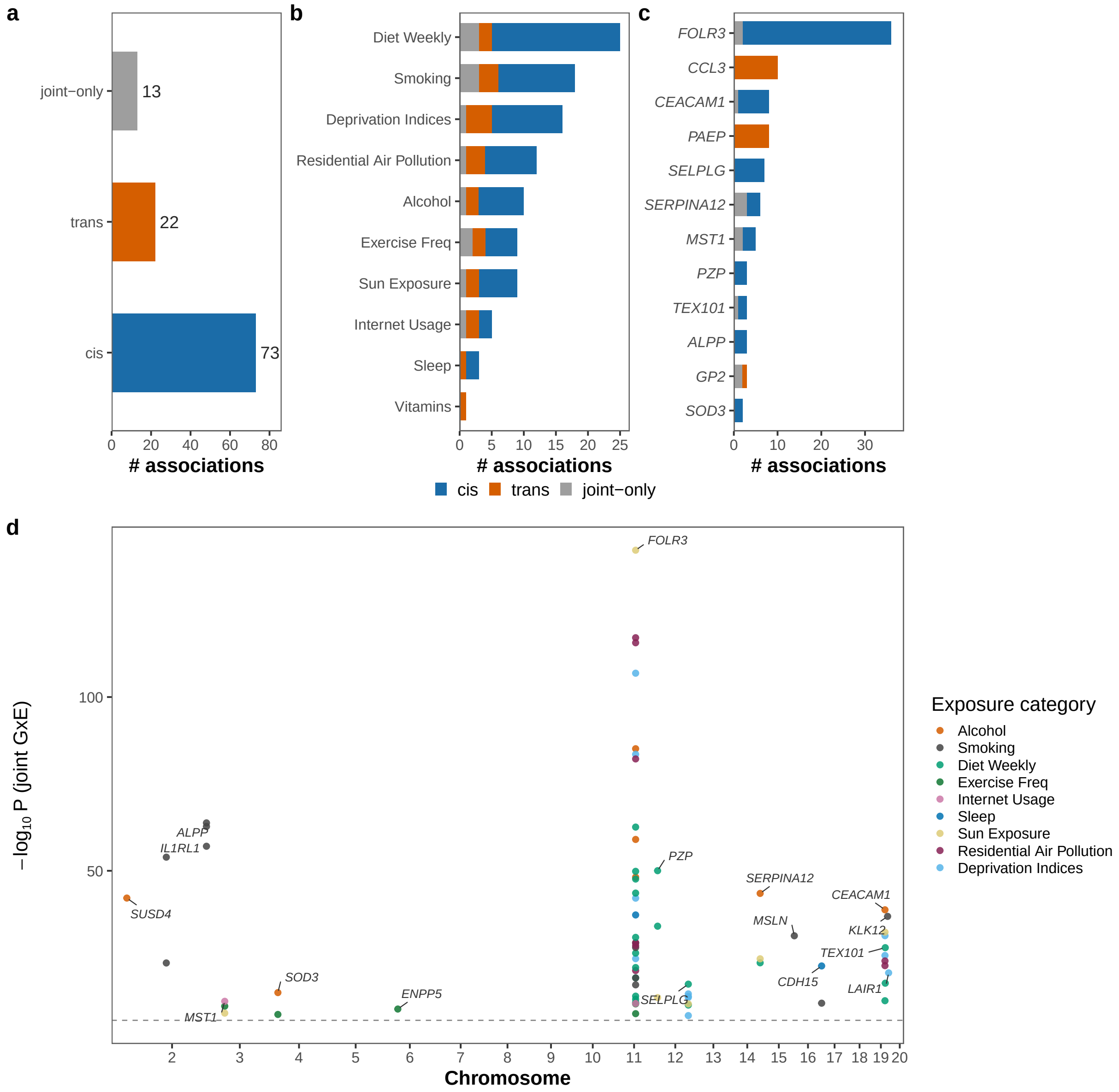


**Supplementary Figure 16: Architecture of replicated polygenic gene-by-environment interactions.** Polygenic G×E was tested per protein×exposure pair with a joint $F$-test; the interactions shown are Bonferroni-significant in both 80/20 splits. **(a)** Number of replicated associations for cis-, trans- and joint tests, **(b)** by exposure category, and **(c)** by protein, for the twelve proteins carrying the most interactions. **(d)** Genomic map of the replicated cis interactions, each point is a protein×exposure pair placed at the coordinate of the protein-coding gene. Colored points are Bonferroni significant.

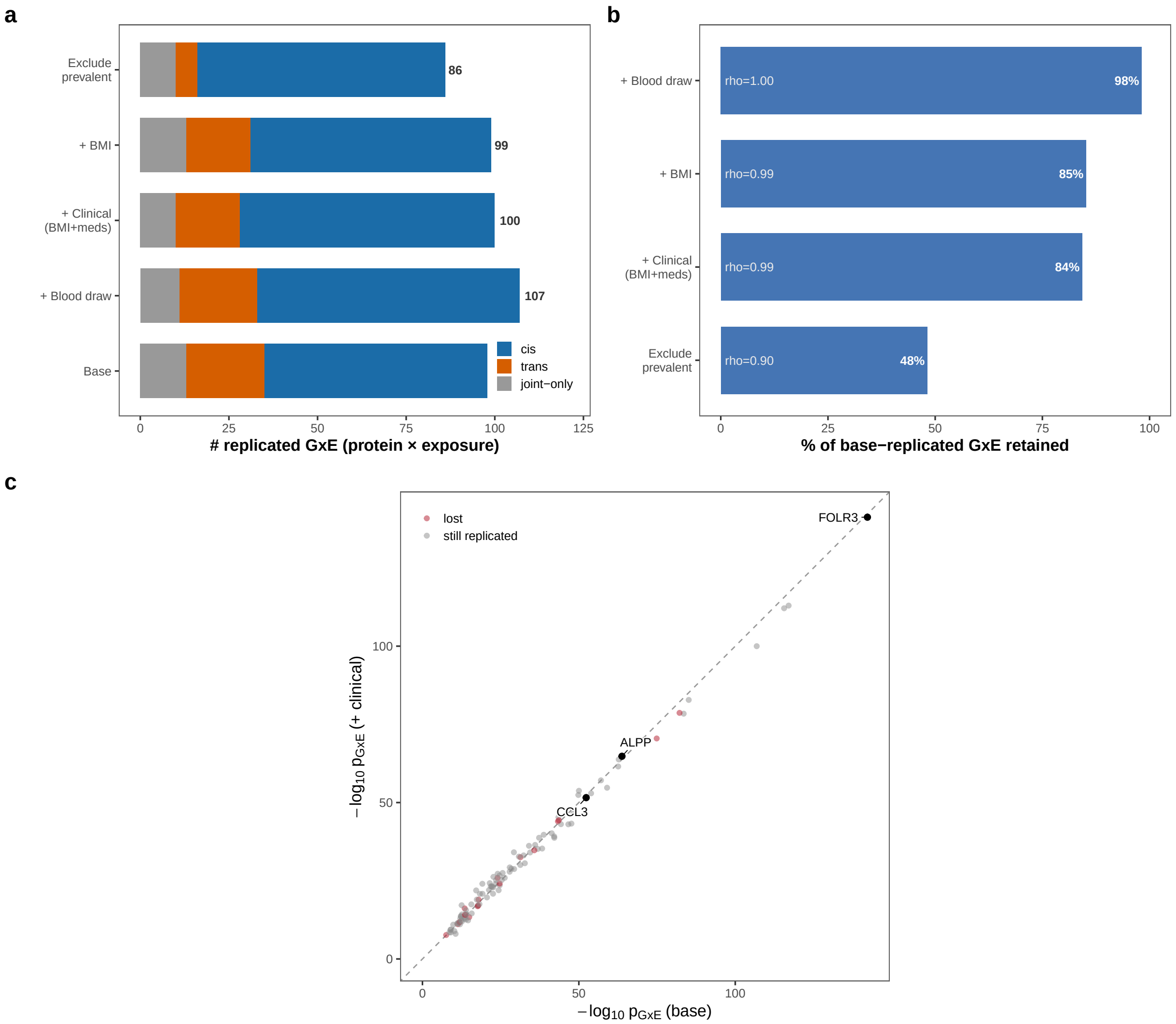


**Supplementary Figure 17: Covariate-specification sensitivity of replicated polygenic G×E interactions.** Base-replicated interactions (joint $F$-test, Bonferroni-significant in both 80/20 splits) were re-estimated under the covariate ladder used for the main effects. **(a)** Replicated interactions per specification. **(b)** Retention of base-replicated interactions, annotated with the Spearman $\rho$ of $-\log_{10} p$ against base. **(c)** Clinical (BMI + medication classes) versus base. Black points denote examples of the most significant pairs.

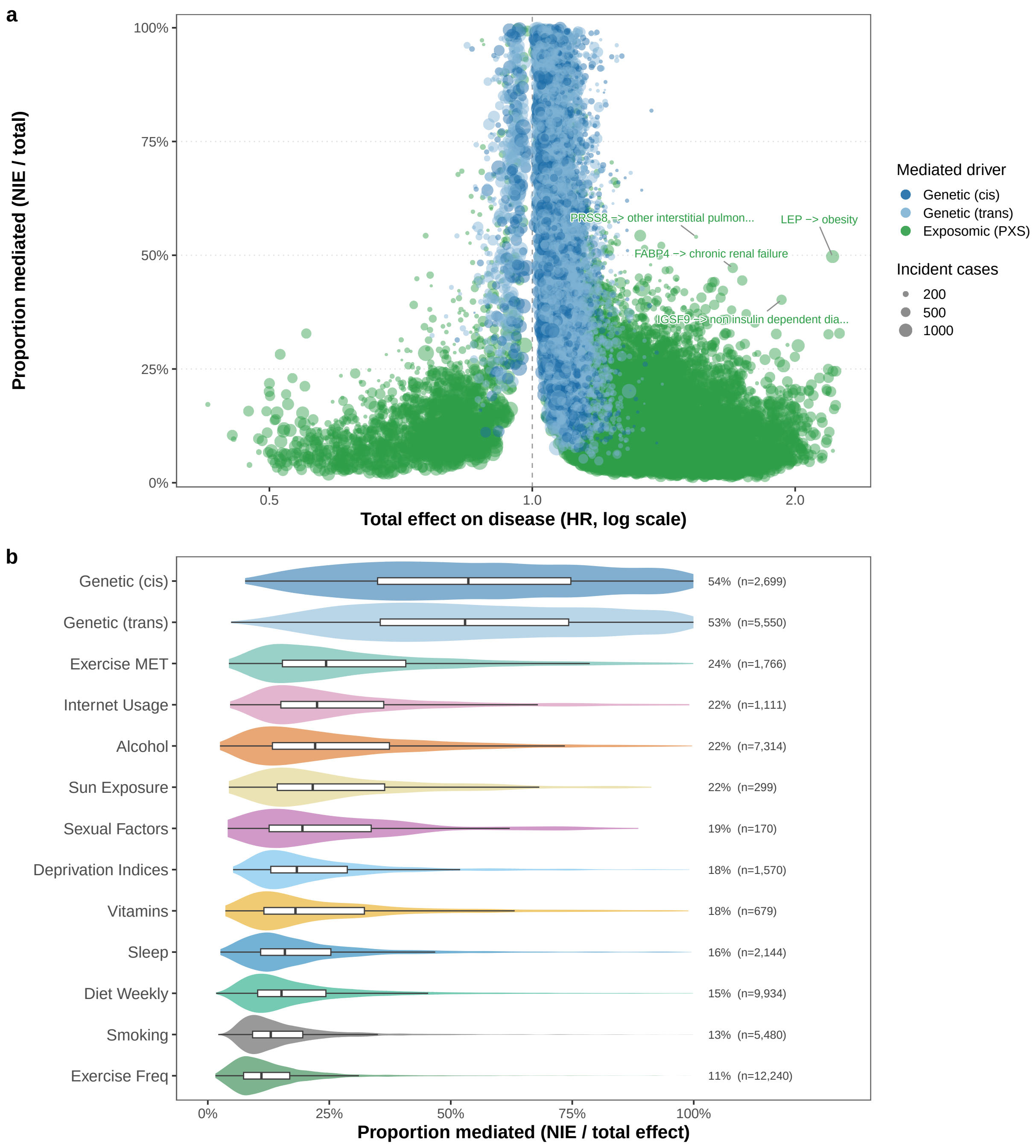


**Supplementary Figure 18: Proportion of a driver's effect on incident disease mediated by a single plasma protein.** GEM splits a driver's (e.g. genetic or exposomic) total effect (TE) into the measured protein's natural indirect effect, NIE and the proportion mediated PM = NIE/TE. The effects are shown in Cox hazard ratios per SD. Links with an FDR-significant NIE ($q < 0.05$) sign-consistent with TE are shown. **(a)** Each point is one driver–protein–disease link; dashed line, HR = 1. Four exemplar links are labeled. **(b)** Proportion mediated by a given driver.

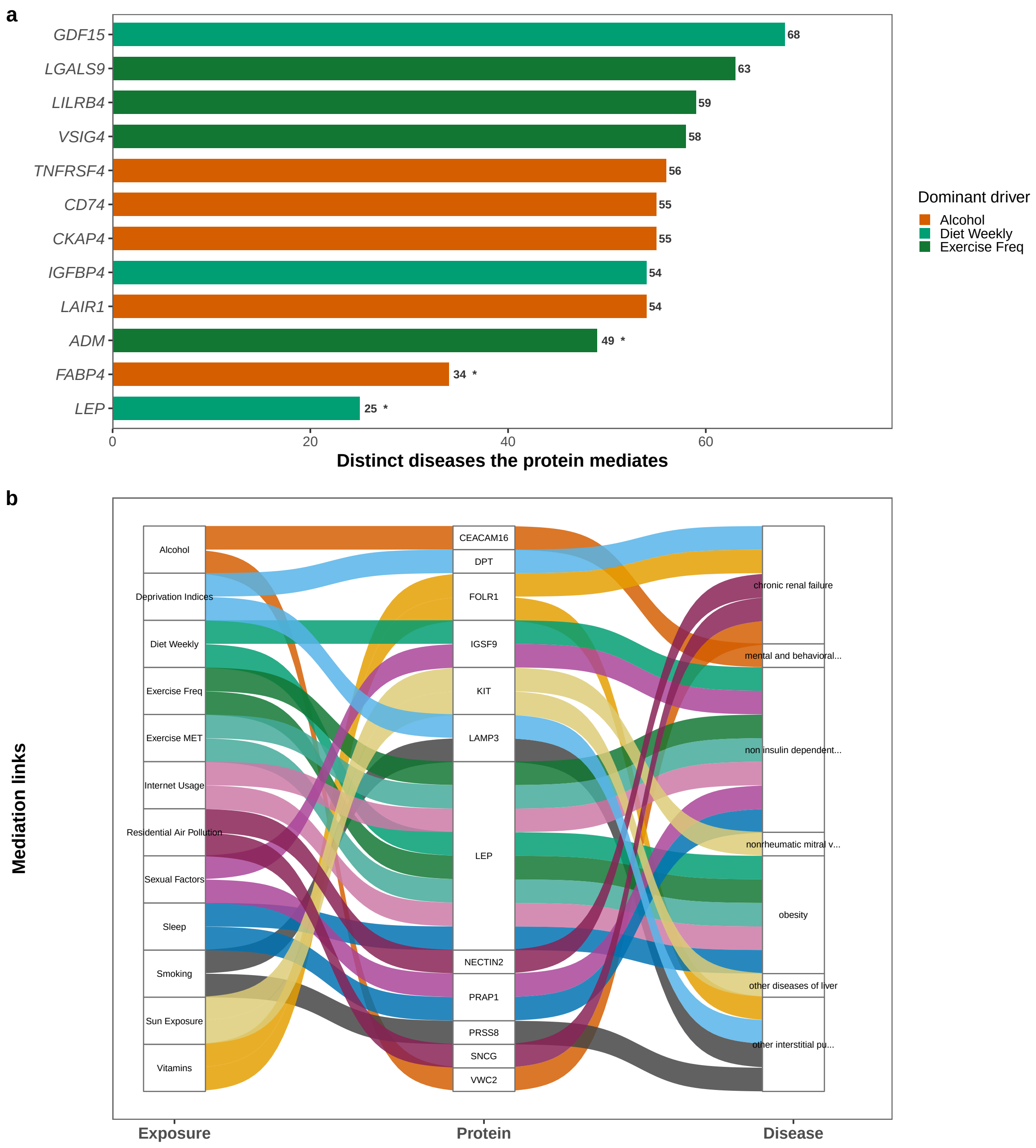


**Supplementary Figure 19: Breadth and structure of protein-mediated exposure–disease links.** A link counts when the Cox GEM natural indirect effect (NIE) from a modifiable exposure to an incident disease through the protein is FDR-significant ($q < 0.05$) and sign-consistent with the total effect. **(a)** Mediator breadth, for the nine broadest mediators plus the three proteins named in the main text (asterisked). **(b)** Exposure $\rightarrow$ protein $\rightarrow$ disease flows for each exposure's two strongest links, ranked by $|\log \mathrm{HR}|$ of the NIE.

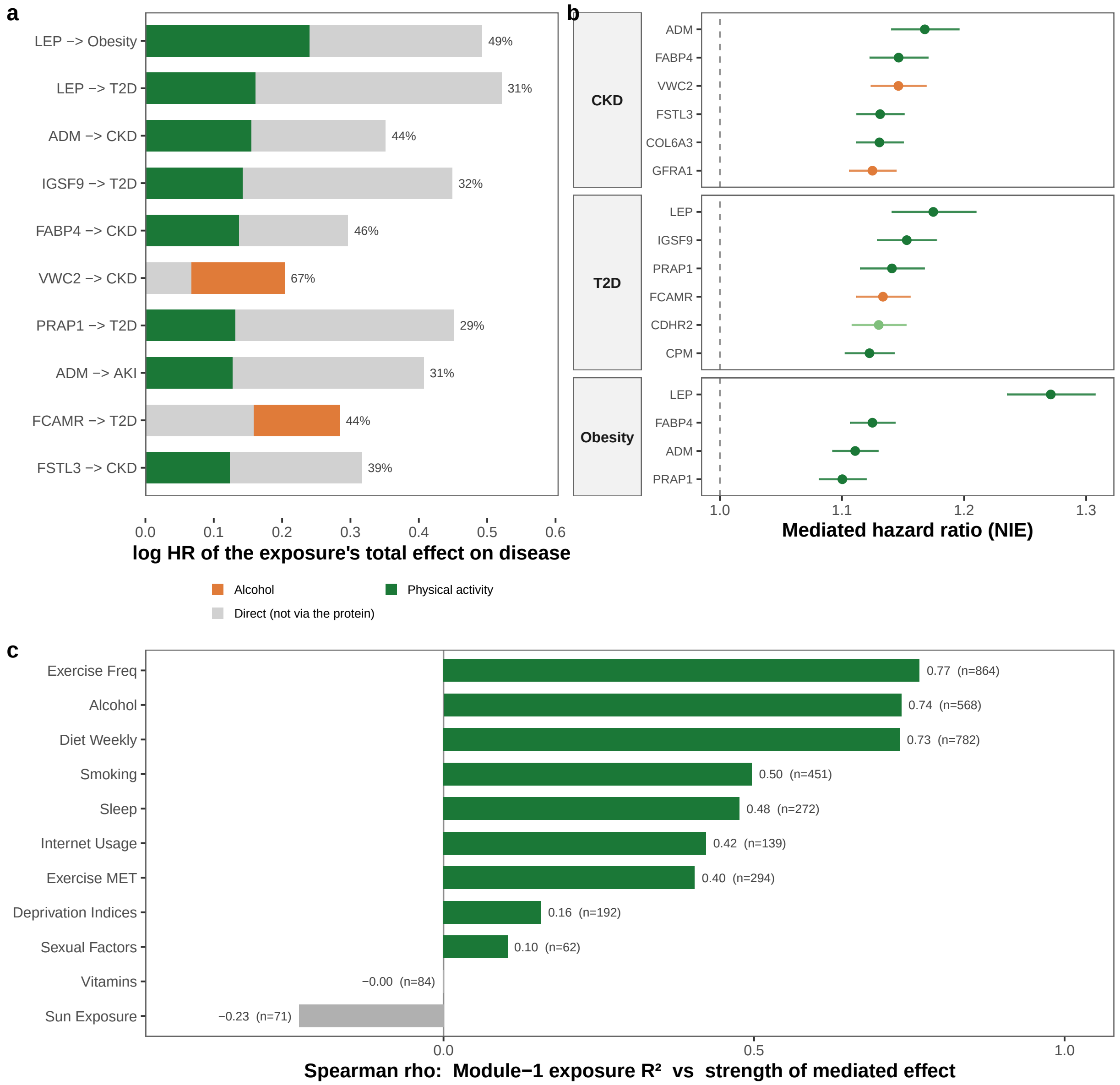


**Supplementary Figure 20: Prioritization of protein intermediaries of modifiable exposures.** All links are FDR-significant ($q < 0.05$), sign-consistent GEM mediations; effects are Cox hazard ratios per SD. **(a)** The exposure's total effect split into its mediated and direct parts, for the ten links with the largest mediated $|\log \mathrm{HR}|$ among those with at least 300 incident cases, one per protein–disease pair; annotations, proportion mediated. **(b)** Mediated hazard ratio (NIE) with 95% confidence interval for up to six intermediaries in each of the three diseases carrying the most strong mediations. **(c)** Spearman $\rho$ across proteins between exposomic $R^2$ and mediated effect strength.

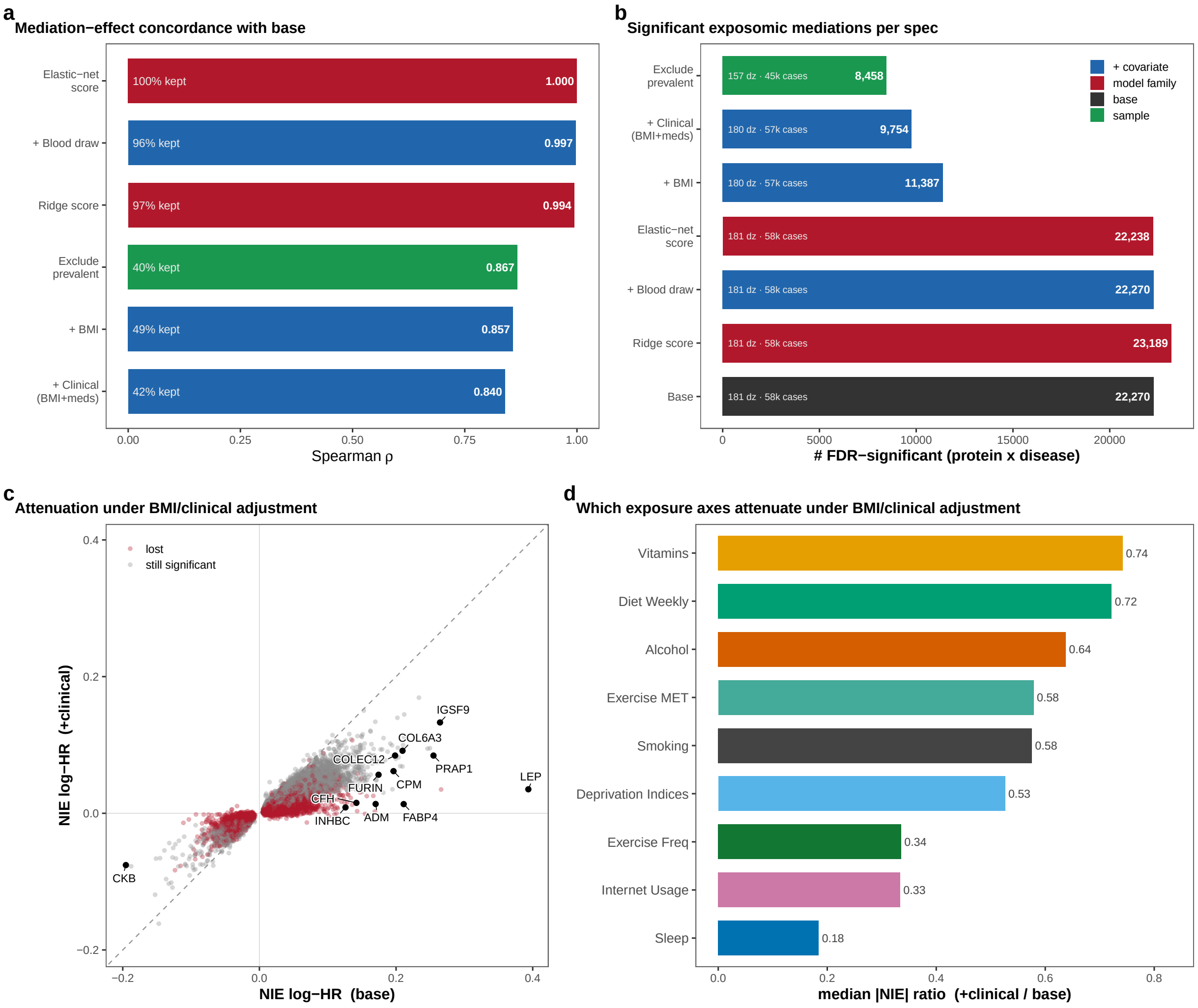


**Supplementary Figure 21: Covariate-specification sensitivity of GEM mediation estimates.** Robustness of the Module 3 mediation natural-indirect-effect (NIE) estimates to the covariate specification, modeling family and analysis sample, evaluated over the exposomic mediations that are FDR-significant under the base/lasso GEM model. **(a)** Spearman $\rho$ of the NIE log-hazard ratio against the base specification, with the fraction of base-significant pairs retained for each specification. **(b)** Number of FDR-significant protein×disease mediations recovered under each specification (bar labels give the number of diseases tested and incident cases), colored by whether the variant alters the covariate set, the modeling family or the analysis sample. **(c)** Per-pair NIE log-HR under the full clinical specification versus base, with representative attenuated mediators highlighted. **(d)** Median |NIE| ratio (clinical/base) per exposure category (<1 indicates attenuation).

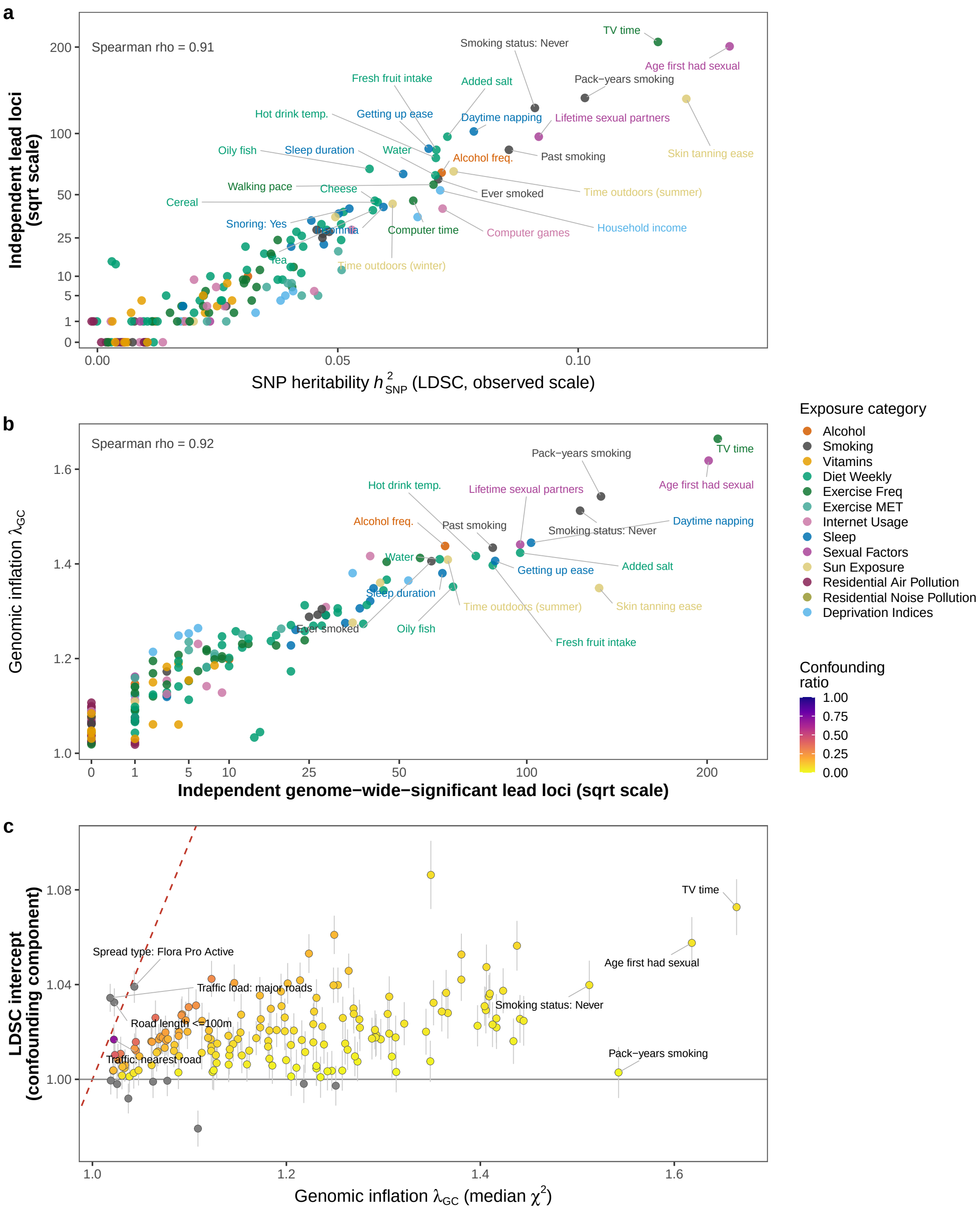


**Supplementary Figure 22: Instrument diagnostics for the exposure GWAS.** Each point is one of the 169 exposure GWAS with an LD Score heritability estimate, from which the Mendelian-randomization instruments were drawn. Independent lead loci are counted by 500 kb clumping at $p < 5 \times 10^{-8}$. **(a)** SNP heritability against lead loci. **(b)** Lead loci against genomic inflation. **(c)** Genomic inflation against the LDSC intercept; bars are ±1 standard error and the dashed red diagonal marks inflation attributable entirely to confounding.

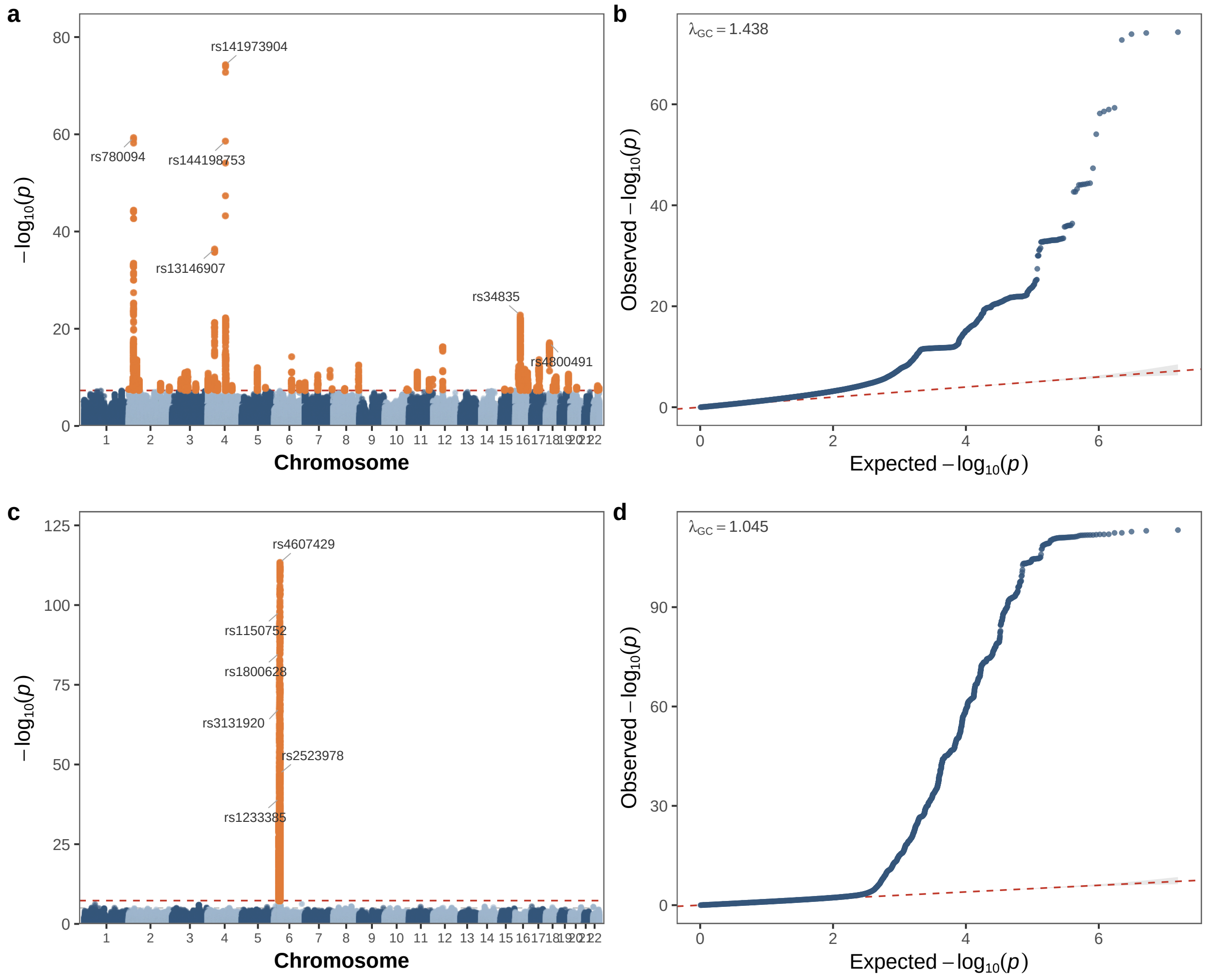


**Supplementary Figure 23: Manhattan and quantile–quantile plots for two exposure GWAS.** Two of the 169 exposure GWAS are shown. In the Manhattan panels the dashed red line is the genome-wide threshold ($p < 5 \times 10^{-8}$). In the quantile–quantile panels the dashed red line is the null expectation. **(a,b)** Alcohol-intake frequency. **(c,d)** Wheat avoidance.

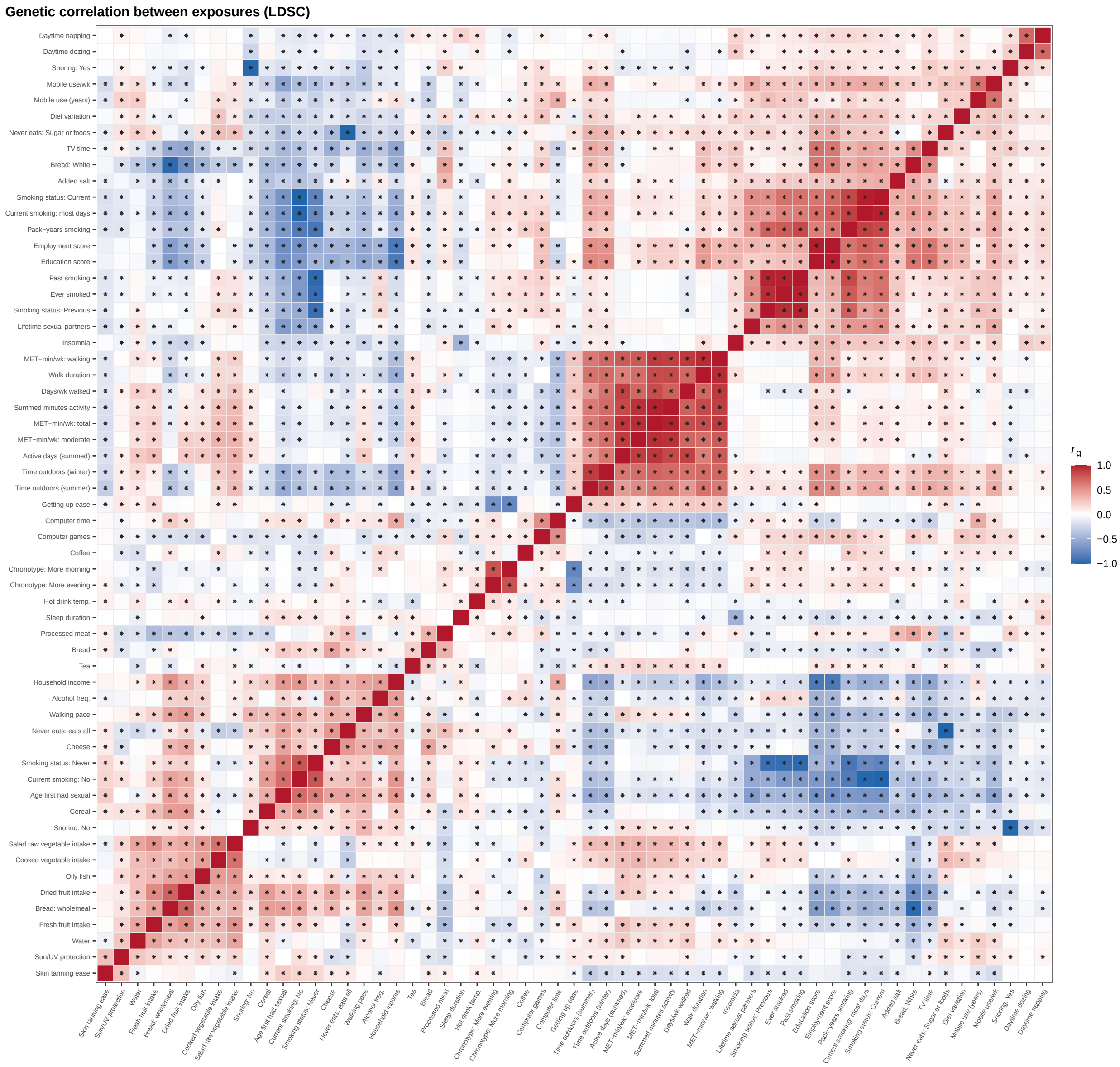


**Supplementary Figure 24: Genetic correlation between exposures.** Pairwise $r_g$ from bivariate LD Score regression, restricted to the 59 exposures whose heritability is estimated to support a bivariate fit. Asterisks mark pairs significant at FDR $q < 0.05$.

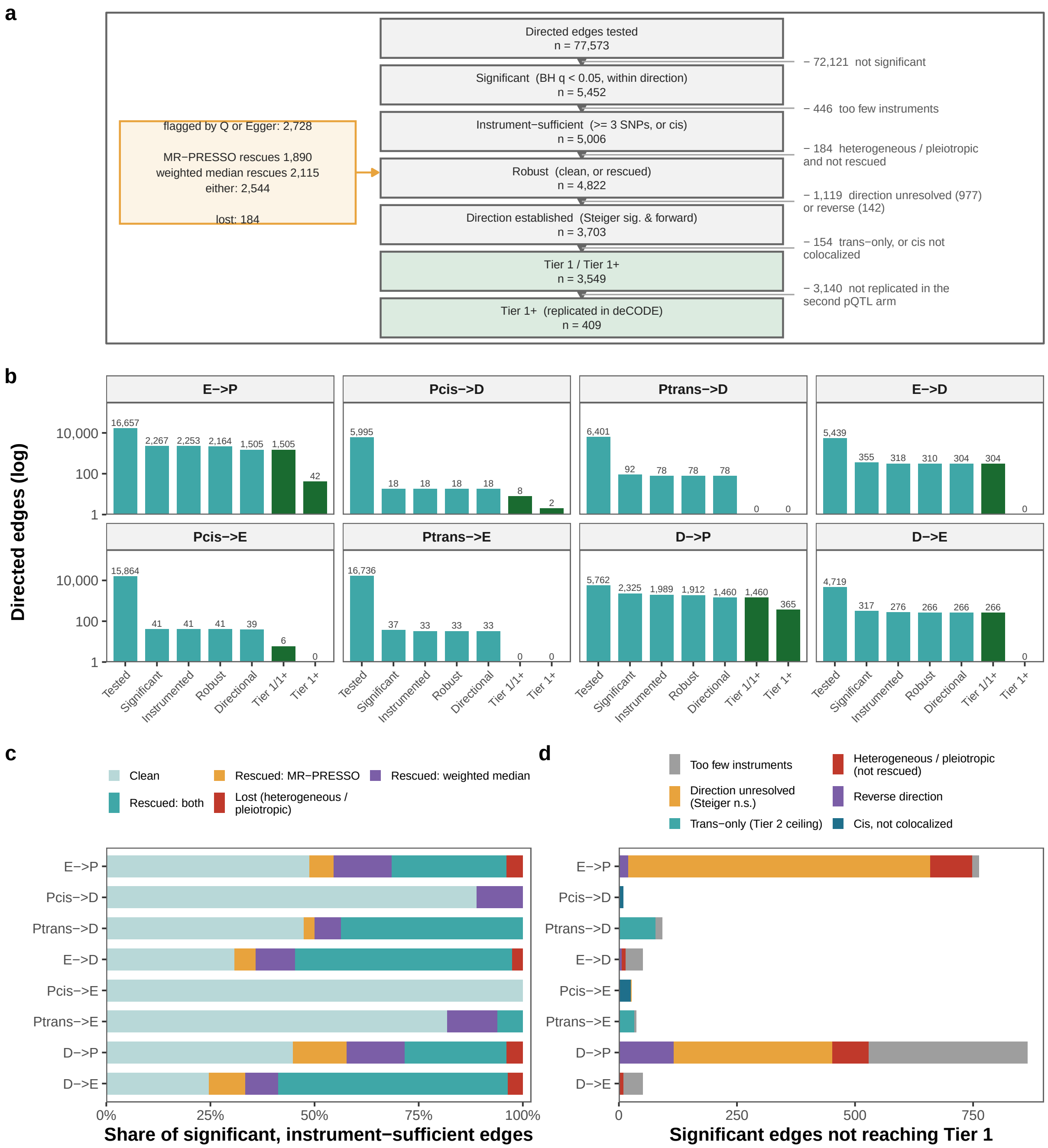


**Supplementary Figure 25: Mendelian randomization edge filtering and tier assignment.** All panels show the UK Biobank arm across eight directed edges among exposure (E), protein (P), and disease (D); $P_{cis}$ and $P_{trans}$ denote the pQTL instrument class. **(a)** Sequential filtering of tested edges by significance (BH $q < 0.05$), instrument sufficiency ($\geq$ 3 SNPs; cis exempt), robustness, Steiger directionality, Tier 1 criteria, and deCODE replication (Tier $1^+$). **(b)** Edge counts through each filtering step, shown on a log scale. **(c)** Robustness outcomes among significant, instrument-sufficient edges, including rescue by MR-PRESSO or weighted median. **(d)** Reasons significant edges did not reach Tier 1.

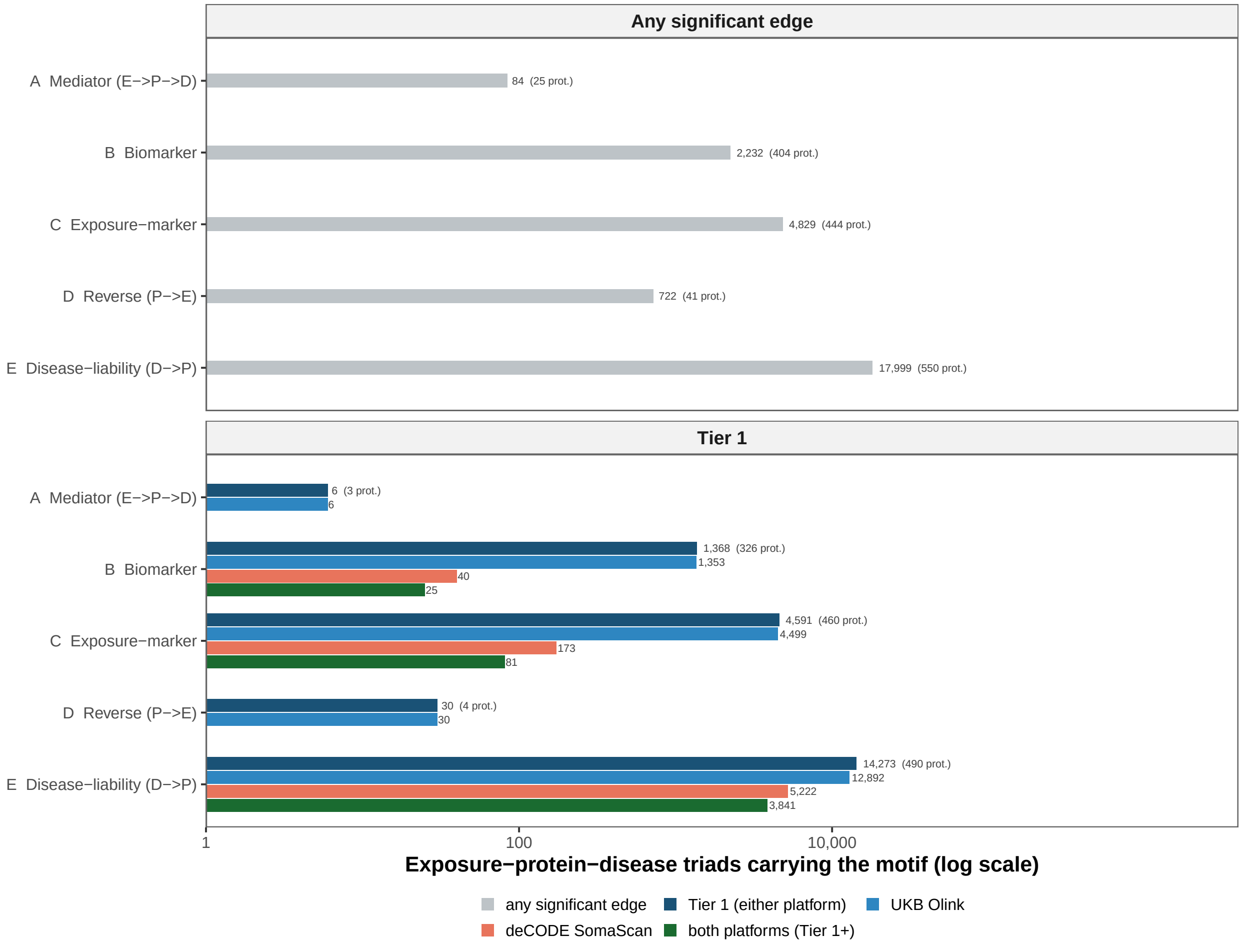


**Supplementary Figure 26: Exposure–protein–disease motif counts under two evidence rules and by pQTL platform.** The top panel counts motifs supported by significant (FDR $< 0.05$) edges, whereas Tier 1 requires all defining positive and negated edges to satisfy Tier 1 criteria. Protein-involving edges are evaluated separately within each pQTL platform. Tier 1 denotes support from either platform and Tier $1^+$ support from both; exposure–disease edges are shared across platforms. Numbers in parentheses indicate the number of unique proteins.

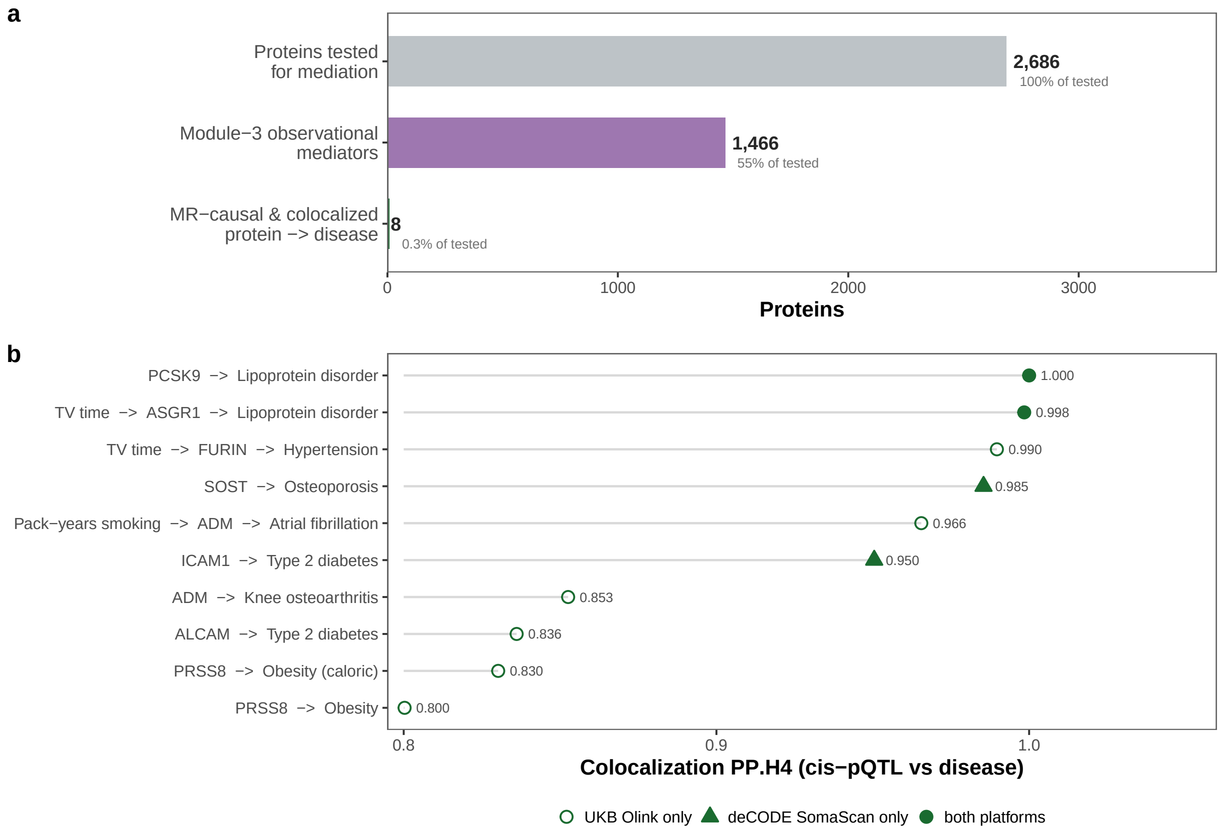


**Supplementary Figure 27: Genetic filtering of the observational mediators. (a)** Proteins retained at each stage. Tested for mediation, observational exposome→protein→disease mediators (FDR-significant, sign-consistent natural indirect effect), and protein→disease Tier-1 cis-pQTL Mendelian-randomization that colocalizes with the disease signal (PP.H4 $\geq$ 0.8). **(b)** Each row is one surviving protein→disease link, ranked by colocalization posterior. An exposure is shown only where that exposure→protein→disease triad is a Tier-1 mediator. Rows without an exposure have no Tier-1 exposure→protein edge.

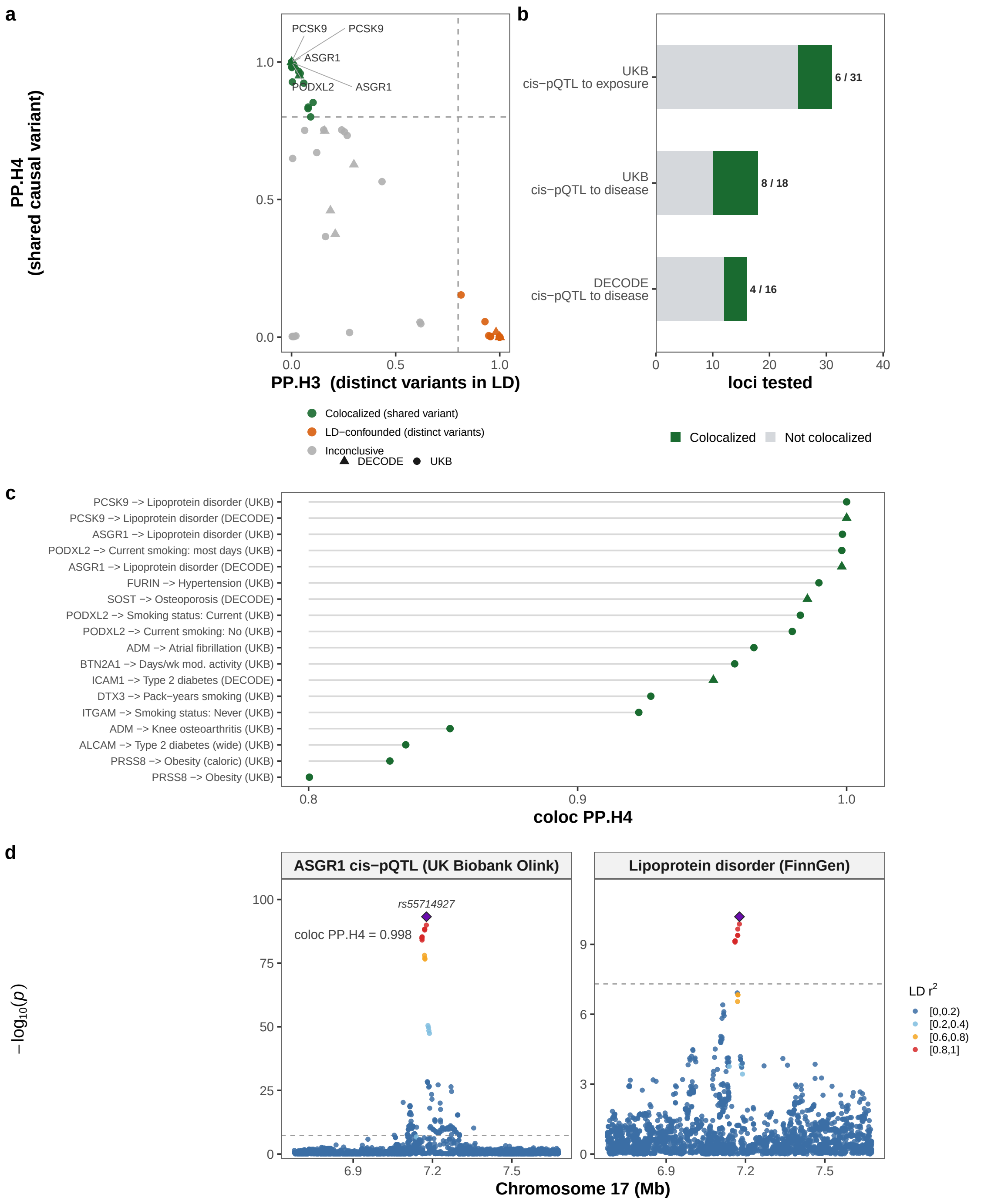


**Supplementary Figure 28: Colocalization of cis-pQTL and outcome signals.** Colocalization (PP.H4 $\geq$ 0.8) one causal variant per trait. **(a)** Each point is one cis-pQTL $\times$ outcome locus test. The five strongest colocalizations are labeled. **(b)** Loci tested and colocalized, by arm and edge type. **(c)** Every colocalized hit, ranked by PP.H4. **(d)** The ASGR1 cis-pQTL, in the UK Biobank Olink arm, and the FinnGen lipoprotein-disorder GWAS over the same chromosome-17 window; the shared lead variant is a diamond and the dashed line the genome-wide threshold ($p < 5 \times 10^{-8}$).

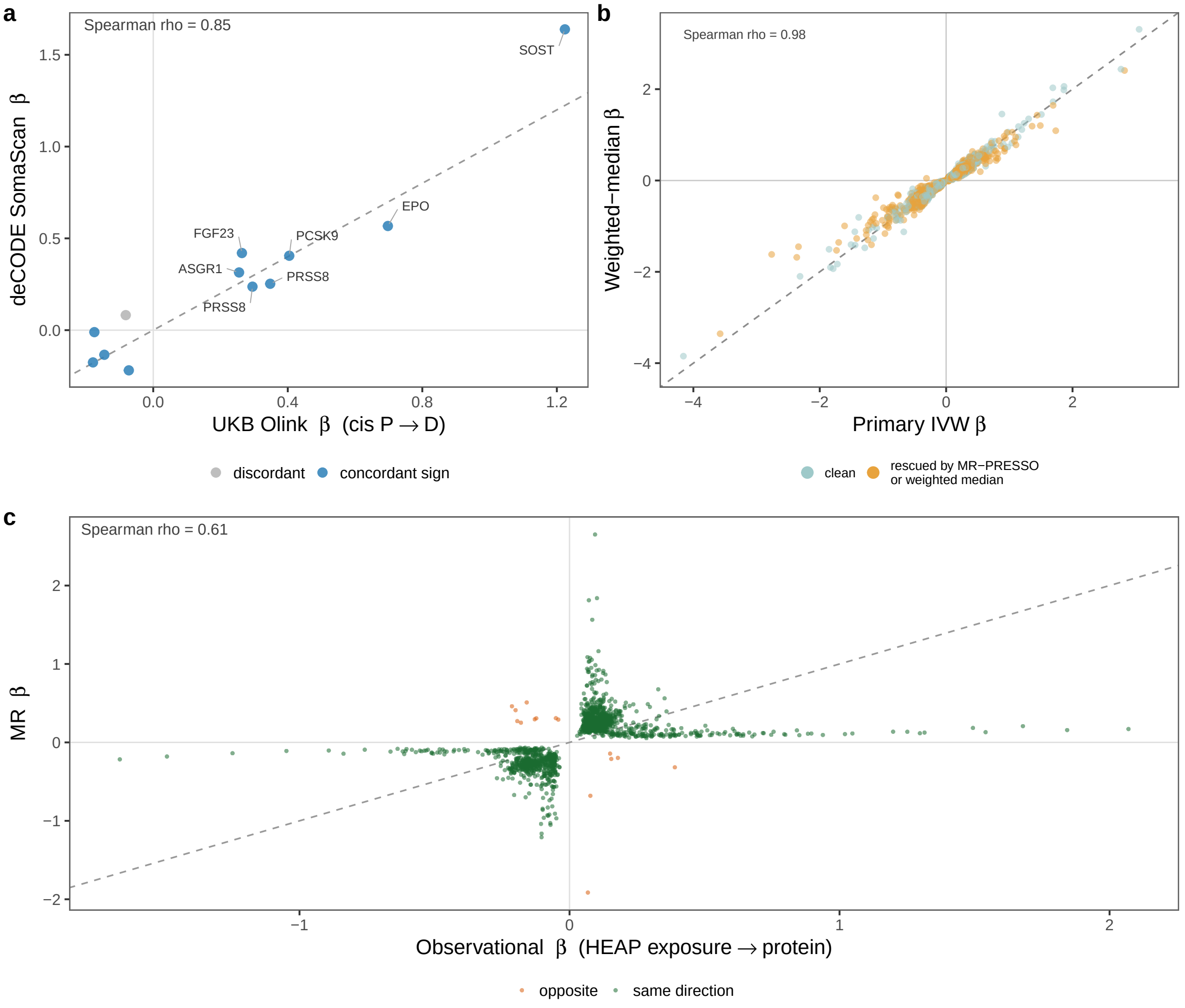


**Supplementary Figure 29: Replication, sensitivity, and observational agreement of the Mendelian-randomization estimates.** **(a)** Two-arm replication of the cis-pQTL protein→disease edges that are FDR-significant in the UK Biobank Olink arm. **(b)** Agreement between the primary inverse-variance-weighted estimate and the weighted median over the Tier-1 hits. **(c)** Exposure→protein pairs with FDR-significant MR support, plotted against the corresponding HEAP observational effect estimated in the held-out split.

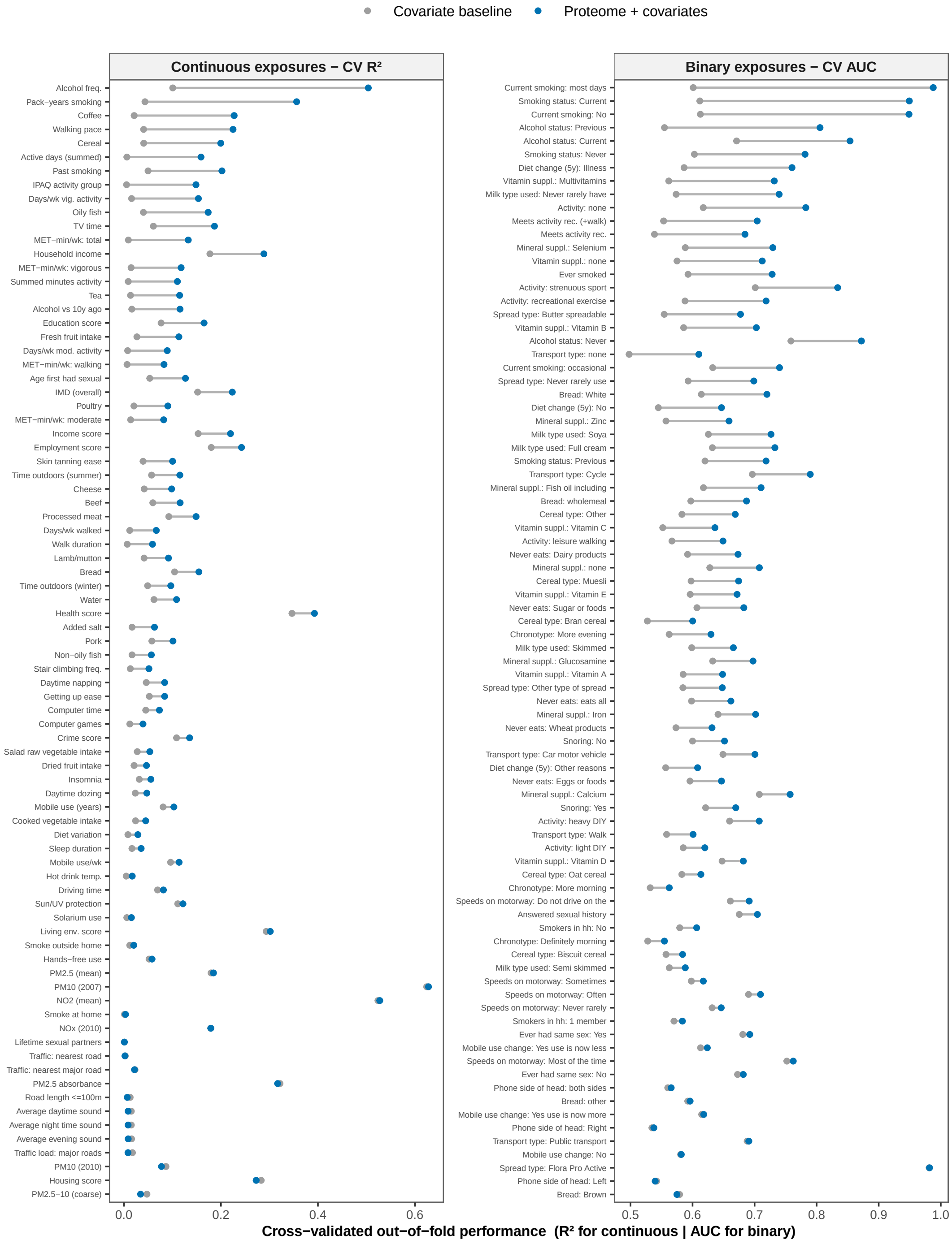


**Supplementary Figure 30: Held-out prediction of exposures from covariates alone versus covariates plus the proteome-based exposure score (PES).** Cross-validated out-of-fold performance is shown as $R^2$ for continuous exposures (left) and AUC for binary exposures (right). Segments show the performance gained by adding PES to the base covariate specification.

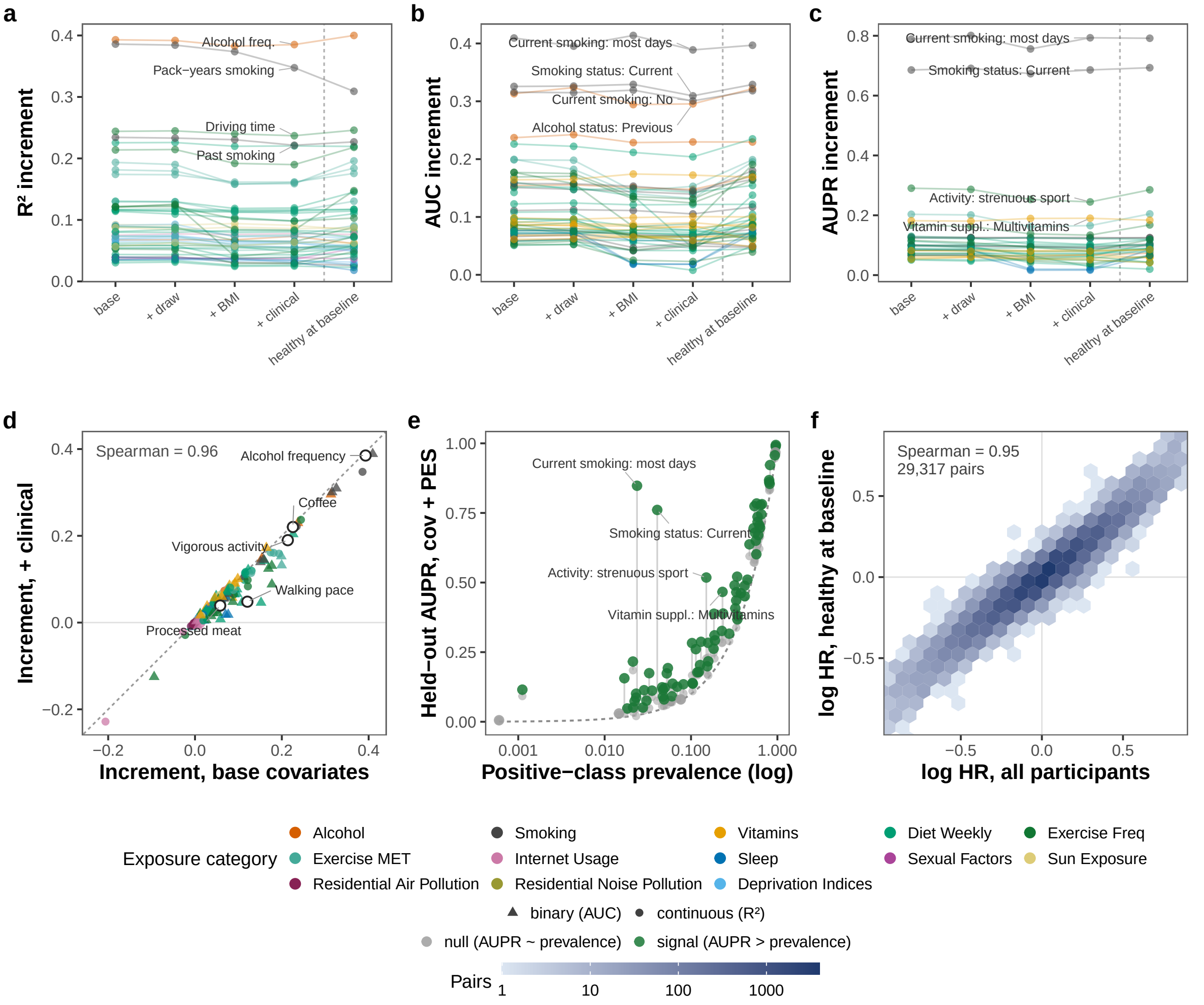


**Supplementary Figure 31: Held-out performance of proteome-based exposure scores (PES) across covariate specifications.** **(a–c)** Incremental held-out performance from adding PES to covariates alone, measured by **(a)** $R^2$, **(b)** AUC, and **(c)** AUPR; lines connect estimates for each exposure across covariate specifications. **(d)** Concordance of performance increments under the base and clinical covariate specifications. **(e)** Held-out AUPR after adding PES versus positive-class prevalence; the dashed line denotes the prevalence null and segments show improvement over covariates alone. **(f)** Concordance of PES–disease associations, shown as log hazard ratios per SD of PES, in all participants versus those healthy at baseline; hexagons summarize exposure–disease pairs common to both analyses.

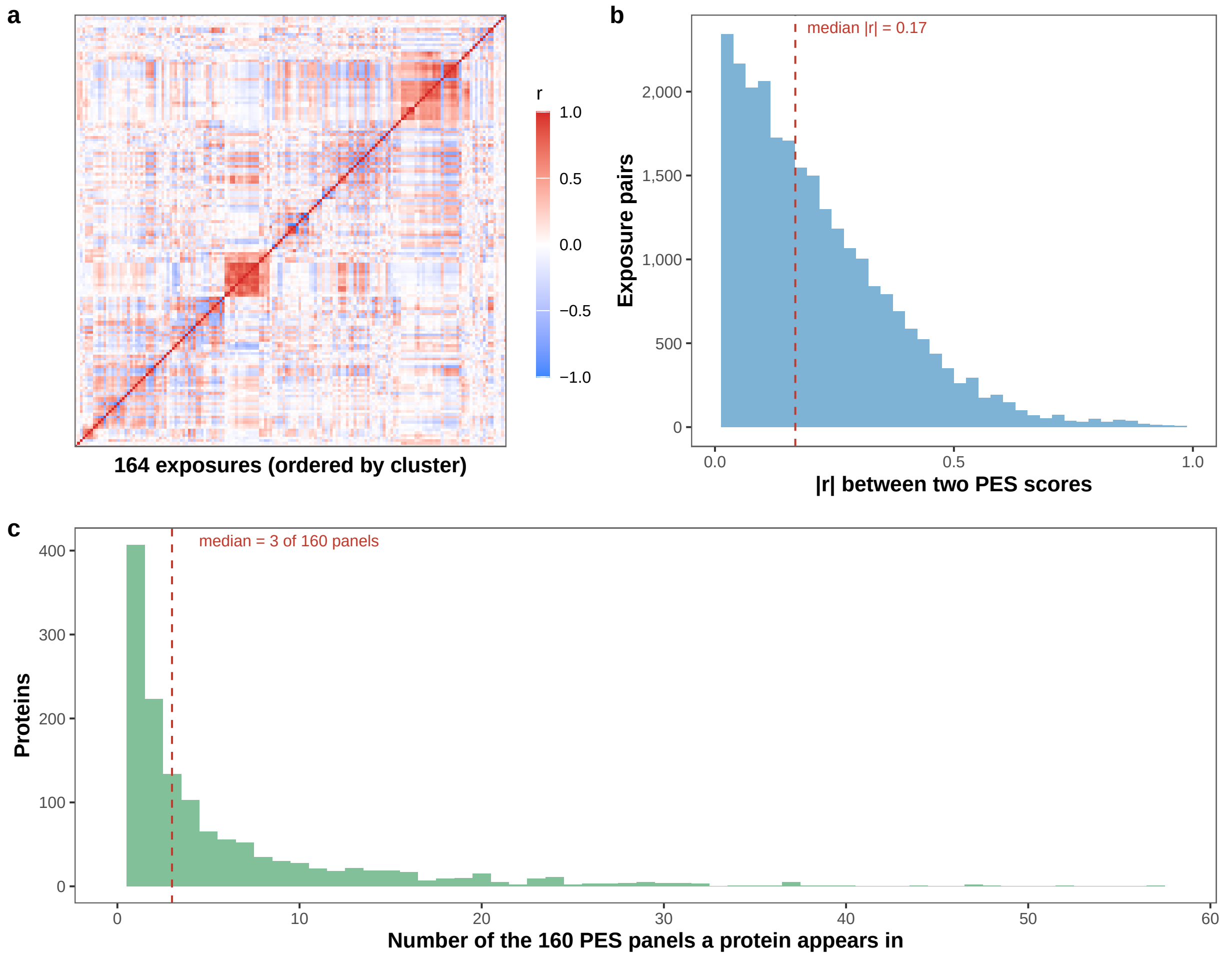


**Supplementary Figure 32: Correlation among proteome-based exposure scores (PES) and protein sharing across scores.** **(a)** Correlation matrix of proteome-only PES in the held-out sample, with exposures ordered by hierarchical clustering. **(b)** Distribution of absolute correlations ($|r|$) across exposure pairs. **(c)** Distribution of the number of PES in which each protein appears, based on the top 50 proteins contributing to each score.

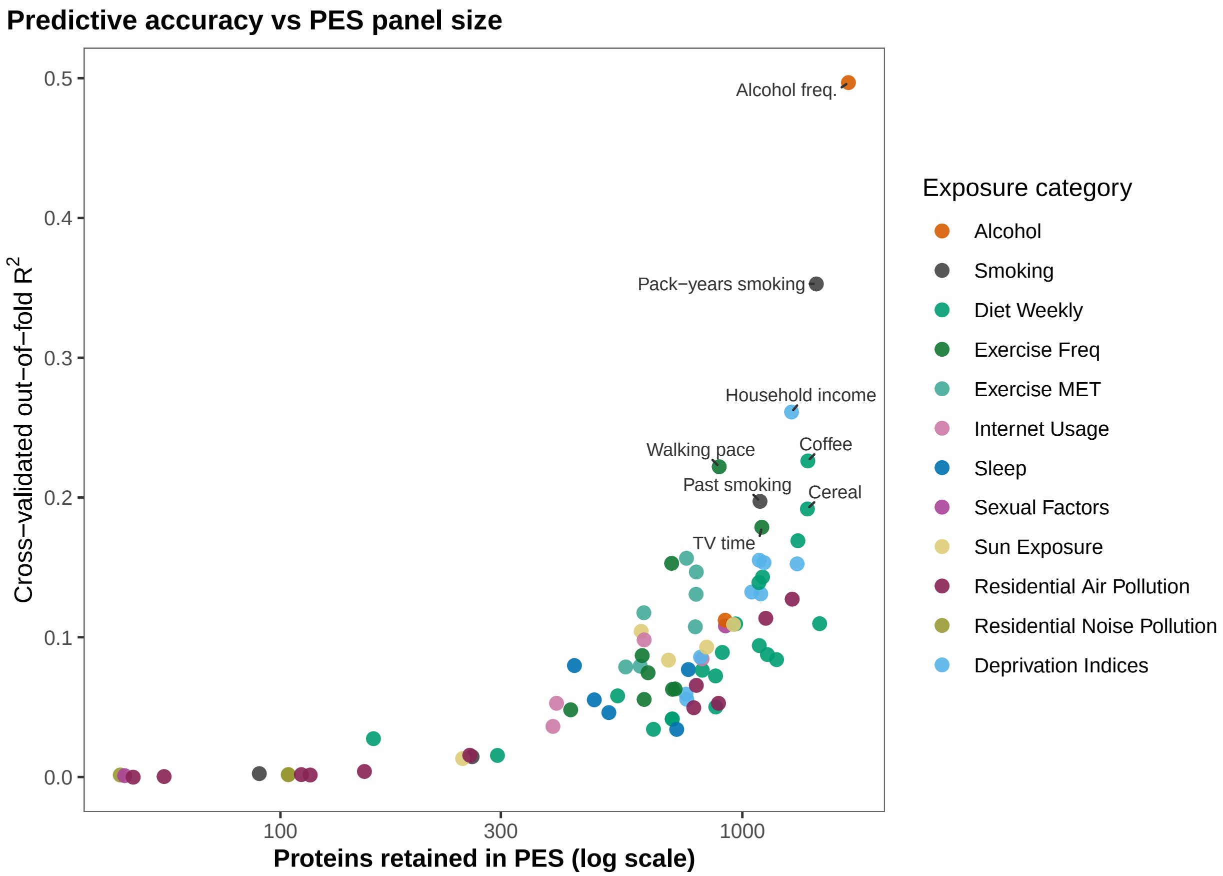


**Supplementary Figure 33: PES accuracy versus retained panel size.** Cross-validated out-of-fold $R^2$ is shown against the number of proteins retained in each proteome-only PES for continuous exposures. Proteins are those retained by lasso at the cross-validation-selected penalty. The eight exposures with the highest $R^2$ are labeled.

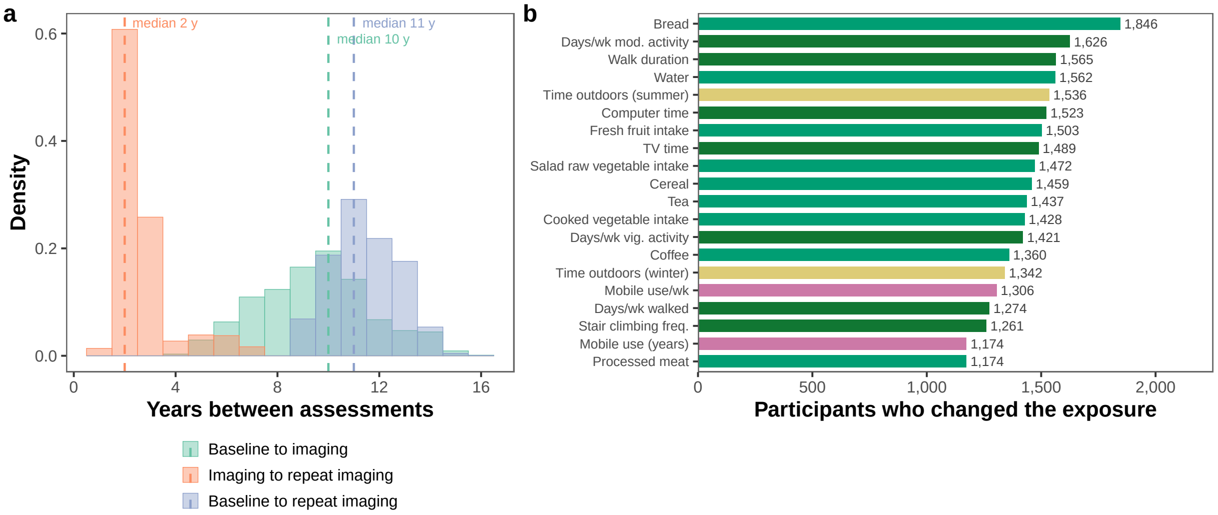


**Supplementary Figure 34: Repeat-visit design behind the within-person tracking estimates.** **(a)** Time between repeat assessments. **(b)** Number of repeat-visit participants who changed their exposure, shown for the top 20 exposures that changed the most.

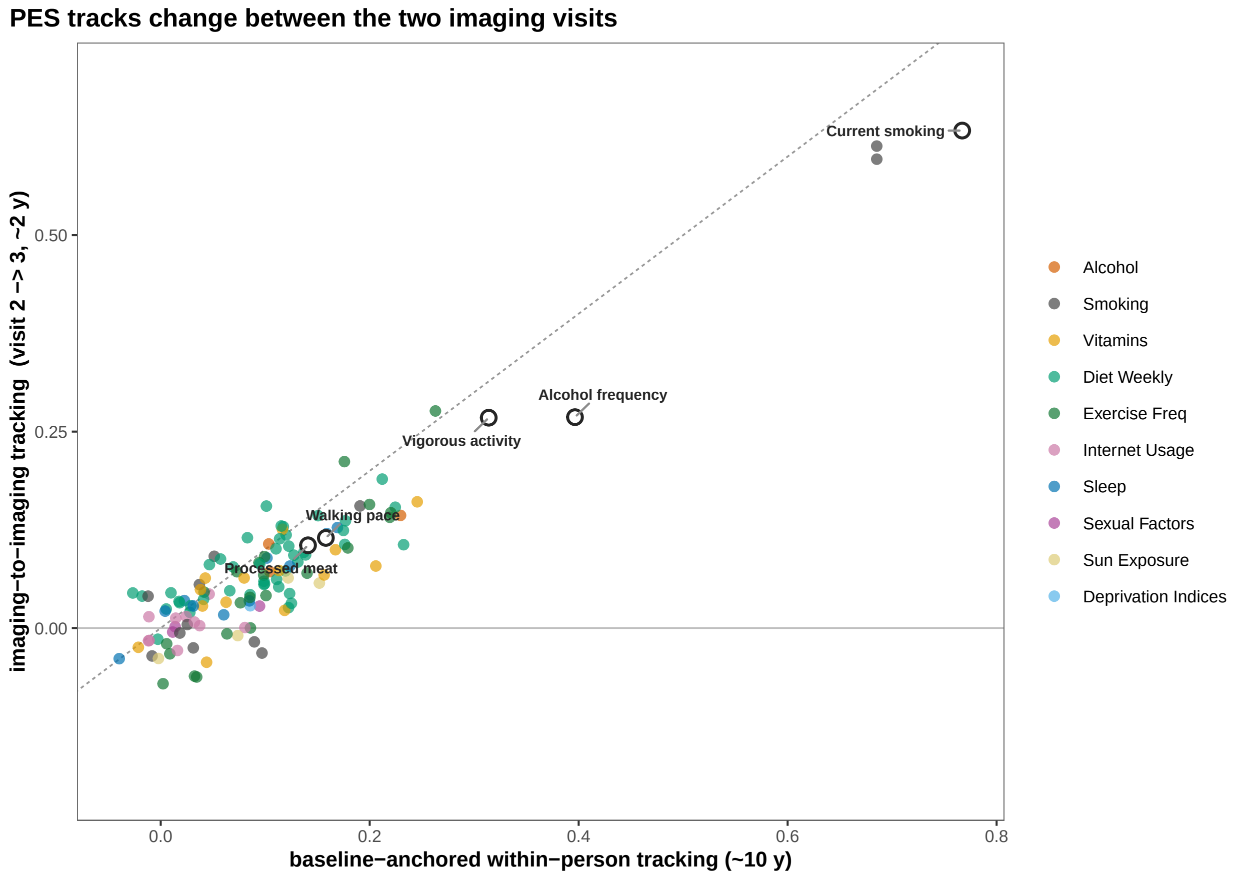


**Supplementary Figure 35: Within-person PES tracking across two longitudinal intervals.** Tracking is the Pearson $r$ between within-person change in reported exposure and change in held-out PES, compared between the two imaging visits (∼2 years) and the baseline-to-imaging interval (∼10 years). Exposures are shown when at least 15 participants with held-out PES at both imaging visits changed the exposure.

### 2 Supplementary Tables

#### Variance decomposition of the proteome

**Supplementary Table 1.** Held-out incremental variance ($R^2$) attributable to covariates, genetics, the exposome and gene-by-environment for each of the 2,686 analyzed plasma proteins, under the base covariate specification.

**Supplementary Table 2.** Distribution across the proteome of each variance component, genetic sub-block, exposure category and gene-by-environment category.

**Supplementary Table 3.** Gene-set enrichment of proteins ranked by their exposomic and genetic signal, against GO and Reactome gene sets.

**Supplementary Table 4.** GREML whole-genome variance components per protein, genetic, exposomic and gene-by-environment, estimated by multi-kernel GCTA-REML on an unrelated subset under the base covariate specification.

**Supplementary Table 5.** Gene-set enrichment of proteins ranked by their GREML variance component, reported separately for the genetic, exposomic and gene-by-environment components.

**Supplementary Table 6.** The per-protein decomposition of Supplementary Table 1 showing cis- and trans-genetic scores separately, and the exposomic and gene-by-environment components split across the 13 exposure categories.

**Supplementary Table 7.** Per-protein training versus held-out out-of-fold $R^2$ for the whole HEAP model, with the fold-to-fold standard deviation and the train-minus-test gap.

#### Exposure–protein associations and enrichment

**Supplementary Table 8.** Replicated exposure–protein associations, Bonferroni-significant in both the training and held-out test partitions.

**Supplementary Table 9.** GTEx tissue enrichment of exposure-associated proteins, by exposure.

**Supplementary Table 10.** Reactome pathway enrichment of exposure-associated proteins, by exposure.

**Supplementary Table 11.** Replicated polygenic gene-by-environment associations, Bonferroni-significant in both the training and held-out test partitions.

#### Mediation

**Supplementary Table 12.** Per-disease counts of significant protein mediators of incident disease, for the exposomic and genetic drivers.

**Supplementary Table 13.** Stability of the mediation results across alternative specifications. Each row re-estimates every mediation link under one alternative and compares it with the base specification.

##### Mendelian randomization and colocalization

**Supplementary Table 14.** Counts of exposure–protein–disease Mendelian-randomization triads carrying each motif, and the number of distinct proteins involved, under both the Tier-1 and nominal evidence bars.

**Supplementary Table 15.** Every exposure–protein–disease triad carrying at least one motif under the Tier-1 definition, with its assigned motif and the supporting Mendelian-randomization estimates.

**Supplementary Table 16.** Colocalization of cis-pQTL Mendelian-randomization edges for the UK Biobank and deCODE instrument arms, each locus tested against the GWAS of its target.

##### Interventional Concordance

**Supplementary Table 17.** Concordance between HEAP exposure–protein effects and the proteomic shifts observed in the HERITAGE, STEP1 and STEP2 trials, for all 169 exposure signatures.

##### Proteome-based exposure scores

**Supplementary Table 18.** Held-out predictive accuracy of each proteome-based exposure score, by exposure and category. The metric column states whether accuracy is reported as AUC or $R^2$.

**Supplementary Table 19.** Held-out Cox C-index for each exposure–disease pair under four nested models: covariates alone, covariates plus the score, covariates plus the self-reported exposure, and covariates plus both.

##### Study definitions

**Supplementary Table 20.** Full list of candidate exposomic features, giving the category, UK Biobank field, variable type, baseline missingness, and whether the feature entered the analysis.

**Supplementary Table 21.** Incident-disease outcomes analyzed by mediation, with the UK Biobank first-occurrence ICD-10 field and the maximum number of incident cases.

#### 3 Supplementary Data

**Supplementary Data 1. Variance decomposition.** Per-protein variance-component $R^2$ under every alternative specification, at two levels of granularity. Coarse partition separates covariate, genetic, exposomic and gene-by-environment components with cumulative model totals. Finer partition separates cis from trans genetic contributions and splits the exposomic and gene-by-environment terms by exposure category.

**Supplementary Data 2. Exposure–protein associations.** Exposure–protein association estimates for every model term and protein, repeated under each covariate specification.

**Supplementary Data 3. Gene-by-environment associations.** Polygenic gene-by-environment association estimates, repeated under each covariate specification.

**Supplementary Data 4. Mediation of incident disease.** Exposome and genetic mediation decompositions for each protein and disease, under alternative covariate specifications.

**Supplementary Data 5. Mendelian randomization.** All Mendelian randomization edges with their confidence tiers, covering every exposure, protein and disease direction tested, reported separately for the UK Biobank Olink and deCODE SomaScan instrument panels, together with the full set of exposure–protein–disease triads evaluated.

**Supplementary Data 6. Intervention concordance.** Per-protein effects from the HERITAGE and STEP1/STEP2 trials alongside the corresponding UK Biobank exposure–protein estimates.

**Supplementary Data 7. Proteome-based exposure scores.** Performance of the proteome-based exposure scores (PES) in how well the PES reads its exposure, tracks within-person changes in exposure, and predicts incident disease onset under alternative adjustments.

**Supplementary Data 8. Proteome-based exposure score weights.** Deployable score weights for each exposure with the protein beta coefficients and metadata needed to compute a score in another cohort.
